# Enhancing Scientific Validity by Integrating EHR and Claims Data in the National Clinical Cohort Collaborative

**DOI:** 10.64898/2026.10.12.26365348

**Authors:** Steve Makkar, David Bogumil, Taylor Estepp, Ayya Keshet, Nathan Hotaling, Kristen Hansen, Ken Gersing, Hythem Sidky, Therese N. Tripler, the N3C

## Abstract

**Purpose:** Accurately capturing a patient’s health journey is essential for ensuring the validity of scientific and public health studies. Electronic health records (EHR) and administrative claims are commonly used in healthcare research, albeit separately. Previous research has demonstrated that EHR and claims capture different aspects of patient history and care, potentially introducing bias when used alone. In this study, we leveraged the largest public US EHR resource, the National Clinical Cohort Collaborative (N3C), alongside Centers for Medicare & Medicaid Services (CMS) claims data to compare their information coverage and agreement.

**Methods:** We integrated N3C EHR and CMS data to create one cohesive dataset and quantified the proportion of key variables present in EHR alone, CMS alone, and the overlap present in both data sources. Overlap in information coverage was quantified pre- and post-landmark diagnosis of COVID-19. To evaluate the impact of data coverage differences, we conducted a case study evaluating the effect of COVID-19 vaccination on ischemic stroke using both data sources alone and in combination.

**Results:** CMS data helped identify many individuals that were duplicated among EHR identifiers. Examination of the overlap showed that CMS data more completely captured conditions, while EHR data more completely captured medications after COVID-19 diagnosis. Some variables exhibited marked differences over time after landmark COVID-19 diagnosis. Using target trial emulation and self-controlled case series methods, we found that data capture variations altered effect estimates: EHR data alone yielded lower risk of ischemic stroke after vaccination in the target trial emulation (odds ratio 0.47, 95% CI 0.28-0.79), with a smaller, non-significant reduction in the self-controlled case series (incidence rate ratio 0.81, 95% CI 0.64-1.02), whereas analyses using CMS data alone or both sources combined yielded estimates close to the null in both designs.

**Conclusions:** Results indicated that when using either a national database combining multiple EHR sources, or CMS claims data alone, patients’ medical histories are less completely captured compared to when these data sources are combined. Case study results highlighted the value of combining data sources for not only yielding fuller patient health histories and valid scientific results, but for identifying and explaining biases that may emerge in research studies that rely on a single data source.

## Introduction

Electronic Health Records (EHR) and administrative claims are important data sources used to support population health studies, comparative effectiveness research, and public health initiatives. Each data source on its own holds unique strengths and weaknesses that may strengthen (or hinder) the capacity to make valid conclusions from such studies **(Supplementary Figure 1)**. Prior studies have demonstrated that EHRs and claims data capture different aspects of patients’ healthcare experiences and medical histories(1, 2). When using EHR data sourced from one health system, the researcher will very likely not have access to any part of an individual’s healthcare journey that took place in a different health system, leading to incomplete capture of the patient’s medical information and history(1, 3, 4). Claims data, in contrast, excel at capturing the *continuity of care* patients receive across different institutions over time, and thus may more thoroughly identify the full range of patients’ comorbidities and health conditions(5–8). EHR data are particularly good for capturing medication prescriptions, and particularly inpatient medications, lab measures, and procedures in great detail, while such detail is generally lost in claims data due to bundling practices for billing(1, 9). Consequently, each data source on its own may not capture the complete medical history of a patient, ultimately impacting researchers’ ability to ascertain exposures, identify occurrence of adverse outcomes, perform valid statistical analyses, and infer accurate conclusions.

Although EHRs excel at capturing prescriptions, studies indicate a relatively high rate of *primary nonadherence* (i.e., failure to fill or obtain a newly prescribed medication the first time it is prescribed), indicating EHRs may overestimate actual medication exposure(10). Estimates of primary non-adherence can vary largely, from 5%-13% for newly initiated oral medications(11), 23%-55% for new medications for chronic use in adults(11), 22% of all prescriptions, and 28% of new prescriptions(12). While estimations of primary non-adherence are likely highly dependent on population and medication, there are few ways to validate medication use with EHR data alone. But, when medications are present and paid for in claims data, researchers can be more confident in patients’ adherence to prescriptions(12).

The relative strengths and weaknesses of EHR and claims data, and improvements in coverage regarding patients’ healthcare history from combining both data sources have been examined by previous researchers, but often within limited domains of medical information. Young and colleagues found that the choice of data source presented substantial discrepancies in patient data related to opioid use (1), but did not examine or measure the differences in other medical information or provide guidelines about effective ways to integrate EHR and claims data for the purposes of conducting scientific research. Fischer et al. compared e-prescriptions to filled claims to study primary non-adherence (12), thereby assessing the EHR prescriptions that were not subsequently present in linked claims data. Their analysis, however, was limited to data from one state and did not evaluate the case of prescription fills in claims data that were not present in the linked EHR. Lin et al. assessed EHR continuity and patient data completeness in two separate EHR health system data sets linked through claims data (3, 4), but settled for using the portion of EHR observations with the highest completeness, effectively dropping over half of their observations, and failing to evaluate any potential increases in information coverage by using the linked claims data. Collectively, the major focus of prior studies has been medication overlap, but comparison of conditions, procedures, and outcomes such as mortality, or changes in data overlap over time have not been thoroughly studied.

Each of these previous studies examined relatively limited domains of information overlap between EHR and claims data and its potential effect on scientific research. Our primary goal, therefore, was to characterize the degree of overlap between EHR and claims data across a more comprehensive scope of patient medical information (e.g., vaccines, medications, condition, procedures, and outcomes) in the largest US EHR resource, the National Clinical Cohort Collaborative (N3C), which brings together a wide variety of healthcare systems across the country, alongside available linked Centers for Medicare & Medicaid Services (CMS) claims data. In addition to examining the degree of overlap and discrepancy between each data source, we undertook a process of integrating the EHR and claims data to produce a dataset that combined all medical information across each source and provided a summary of this process and its challenges. Finally, the extent to which overlap (or lack thereof) impacts the results of scientific research studies has also not been thoroughly investigated. We therefore used each data source independently and in combination to undertake a scientific study related to vaccine safety, to better understand how combining EHR and claims data may yield incremental improvements in the consistency and validity of such studies **(Figure 1)**.

**Figure 1:**
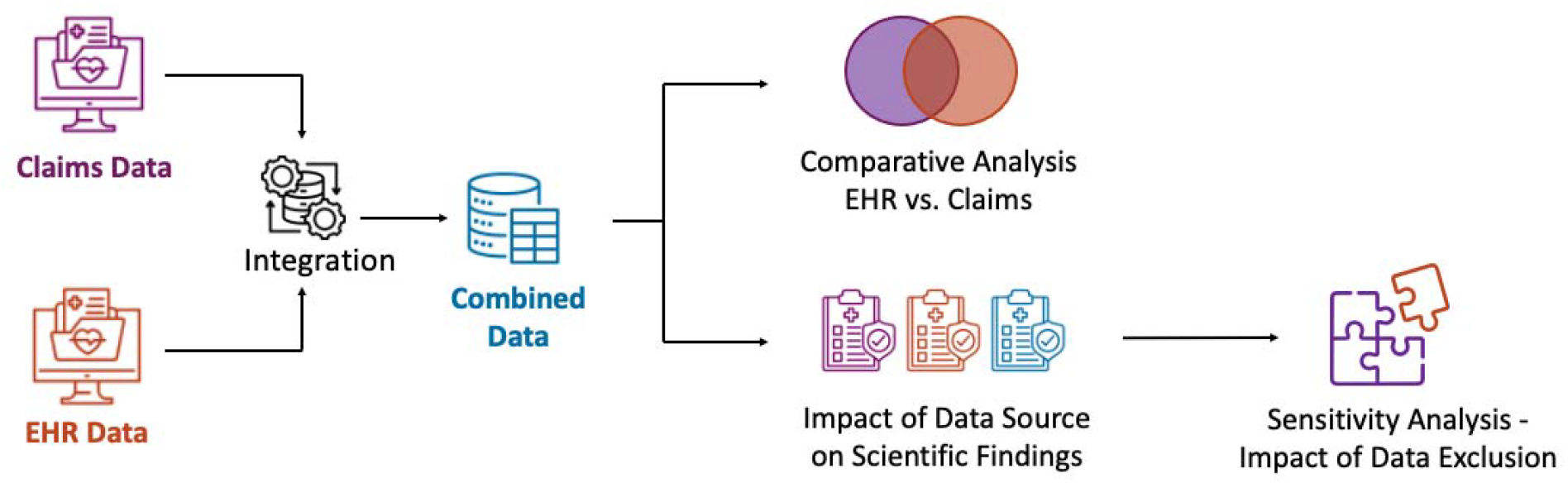
Study design overview.

We utilized two causal inference methods commonly used to examine adverse effects of vaccines: (i) target trial emulation (TTE) cohort study and (ii) self-controlled case series (SCCS)(13, 14). Both methods require having complete data pertaining to vaccinations and outcomes to yield valid estimates of the direction and magnitude of their association. For the SCCS in particular, incomplete data about adverse events, including deaths, is likely to lead to violations of strict statistical assumptions which must be met for the SCCS to yield valid results(15). Cohort studies, in addition, are heavily dependent on having a rich set of pretreatment covariates (e.g., comorbidities or other medications) to adjust for baseline differences between vaccinated and unvaccinated patients which may contribute to confounding(16, 17). If combining EHR and claims leads to more robust and valid scientific research by compensating for the gaps in each respective data source, we would expect to see greater consistency of results across both causal inference methods and with respect to published literature, than analyses using each data source alone.

## Methods

### Data Sources

The study cohort was compiled from N3C (Release Version 197, December, 2025), a centralized real-world repository of de-identified electronic health records supported by the National Institutes of Health (NIH)(18). N3C includes detailed information on clinical encounters for COVID-19 positive patients and matched controls. Records in N3C are aggregated across participating clinical institutions in the US, harmonized using the Observational Medical Outcomes Partnership (OMOP) data model and subjected to quality review and checks (18). All available linked CMS Medicare claims data available in N3C were used. CMS claims covered visits between November, 2009 through December, 2022 for individuals 65 years and older (and those under 65 years with long-term disabilities enrolled in Medicare)(19).

### Data Integration

As a first step, EHR and CMS data were integrated using a linkage key which linked the CMS and EHR person identifiers (IDs). Following linkage, some CMS beneficiaries mapped to multiple N3C person identifiers, likely reflecting care received across multiple participating health systems. These records were consolidated to create a single patient-level record prior to harmonization. Data pertaining to comorbidities (including measurements and observations), exposures (including drugs and procedures), demographics, and COVID-19 related details (including index dates and vaccinations) were harmonized across their multiple N3C records. Most comorbidities were coded as binary indicators, so the maximum value across all records was taken. Patients’ COVID-19 diagnosis date was taken as the earliest date of either an International Classification of Diseases-10-Clinical Modification COVID-19 diagnosis code (U07.1) or a positive SARS-CoV-2 PCR or antigen test across all records. Counts of COVID-19 vaccination doses were summed across all sources, although multiple dose records occurring within a 14-day interval were considered a single dose and merged. After duplicated N3C IDs were consolidated, the same harmonization guidelines were used to combine EHR and CMS data. For demographic information, CMS data was generally given priority, unless missing or otherwise ambiguous. Our strategy for deduplication and harmonization of EHR and CMS data is described in more detail in the **Supplementary Material**.

### Descriptive Analysis of EHR and CMS

Following data integration, overlap between EHR and CMS records was evaluated before and after patients’ COVID-19 diagnosis. Because N3C was established to support COVID-19 research, the COVID-19 diagnosis date was selected as a clinically meaningful landmark for assessing data concordance across sources. Key variables were chosen to span clinically relevant categories of diseases, conditions, procedures, medications, and mortality, and included the exposure (COVID-19 vaccination) and outcome (ischemic stroke) used in the subsequent case study analysis. Overlap, which we quantified as the proportion of patients having a record for a given health variable (e.g., heart disease, ischemic stroke) in both data sources and in each data source alone, was assessed both before and after landmark COVID-19 diagnosis. To better understand in detail the extent of overlap across variables, a more comprehensive set of variables were assessed over varying time periods after COVID-19 diagnosis (starting 30, 90, and 180 days after COVID-19 diagnosis).

### Case Study Analysis of EHR and CMS

To evaluate the extent to which combining EHR and CMS data affected scientific conclusions due to increased coverage of patients’ medical history, the association between COVID-19 vaccination and ischemic stroke was estimated using EHR and CMS data separately, and then in combination. Two causal inference methodologies were implemented to address the research question: a cohort study applying TTE principles, and an SCCS approach.

#### Target Trial Emulation Study Population

We specified a randomized target trial that evaluated whether first exposure to a COVID-19 vaccination increased the risk of first incidence of ischemic stroke in older adults aged 65 and older, between January and December 2021. Patients were eligible for trial entry if they had no prior record of COVID-19 vaccination or ischemic stroke, had complete covariate data for included variables and BMI recorded (EHR and combined, only), did not die within 24 hours of baseline, were in the health system (EHR) or had insurance (CMS) for at least 12 months prior to the start of the trial, no hospitalization or ED visit on baseline or in the 14 days prior, no recorded COVID-19 diagnosis prior to baseline, and were not from a site flagged as having poor data quality (i.e., the site met completeness criteria for mortality/medication/vaccination/follow-up, consistent with N3C data quality assessment procedures; EHR and combined, only) **(Figure 2)**. We applied a sequential trial approach (described below), whereby for each month of 2021 a new ‘trial’ was initiated, leading to a sequence of 12 non-randomized trials. A single patient could contribute to multiple sequential trial emulations provided they remained (i) eligible, (ii) had not experienced ischemic stroke, and (iii) had not yet received a COVID-19 vaccination.

**Figure 2:**
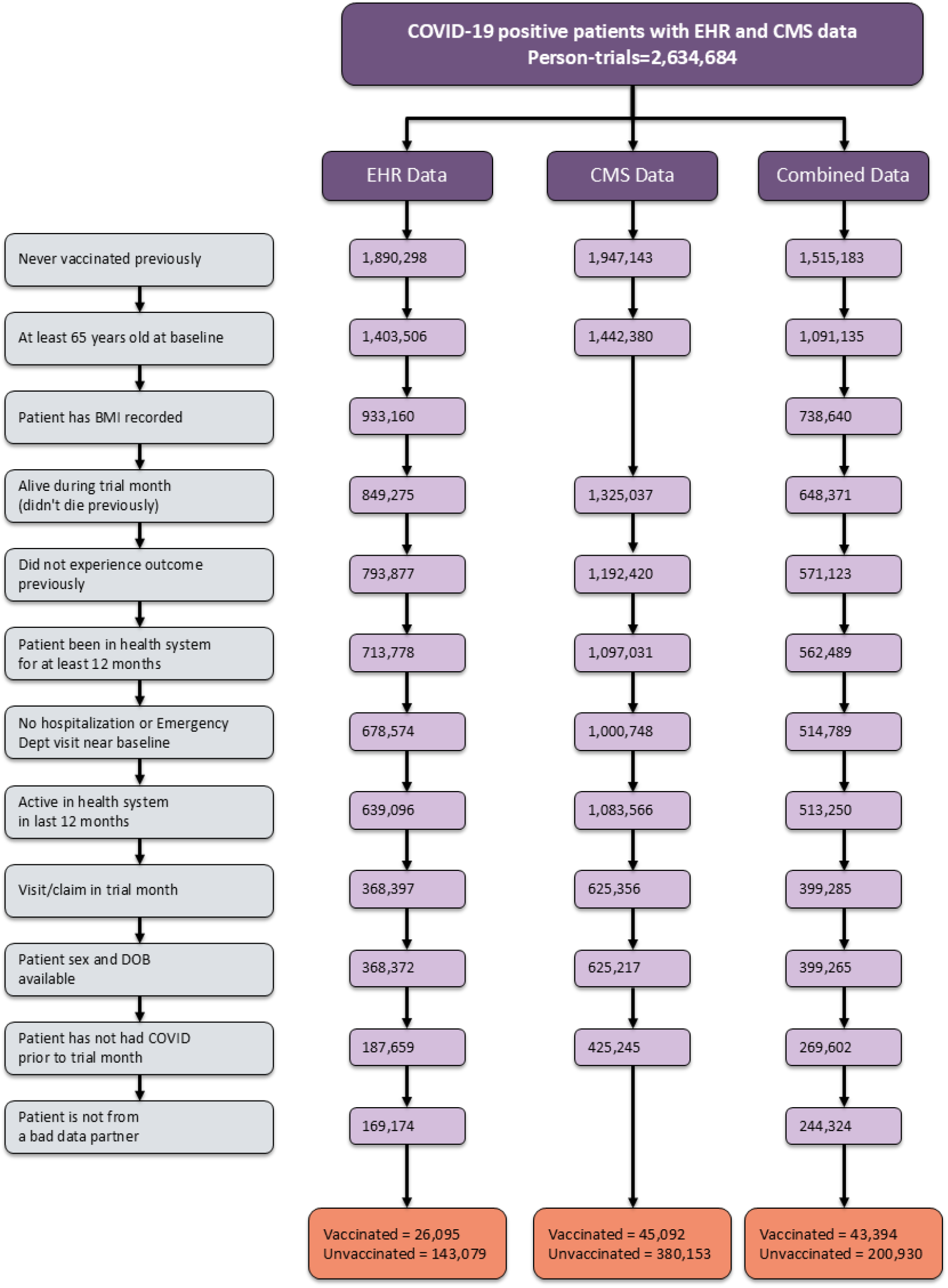
CONSORT diagram showing number of person-trials remaining in each dataset following application of exclusion criteria for the TTE study.

#### Target Trial Emulation Exposure and Outcome

The treatment (exposure) of interest was receipt of a COVID-19 vaccination in the trial month. Eligible patients were classified as either treated, meaning they had a record of COVID-19 vaccination during the trial month, or untreated, meaning there was no record of vaccination during the trial month. Baseline was defined as the vaccination date for treated patients and the first day of the trial month for untreated patients, consistent with the monthly trial emulation framework. The primary outcome was the occurrence of ischemic stroke within 30 days post-baseline.

#### Target Trial Emulation Statistical Analysis

A sequence of monthly target trials was emulated from January through December 2021. In each trial, previously unvaccinated adults aged ≥65 years were classified according to receipt of a first COVID-19 vaccination during that month **(Supplementary Figure 2)**. The causal contrast of interest was the intention-to-treat (ITT) effect (i.e., the comparative effect of being assigned to treatment (vaccinated) versus no treatment (unvaccinated) at baseline, irrespective of whether patients adhered to their assigned treatment strategy post-baseline)(20), measured using the ratio of the hazard of ischemic stroke between treated and untreated patients. Individuals who did not receive the vaccine during the index month could receive vaccination in any subsequent month.

To estimate the ITT effect using the observational data, we calculated inverse probability of treatment weights (IPTW) using the marginal mean weighting through stratification (MMWS) method (16, 21). This involved fitting a propensity score logistic regression model in each trial and predicting the probability of vaccination based on pre-treatment covariates which included prior comorbidities, demographics, health system history, medication exposures, and healthcare institution (available only in the EHR alone and combined analyses). Prior to fitting each propensity model, variable selection was performed in each data source separately to identify covariates that were either (i) related to treatment assignment (i.e., receipt of vaccination) or the (ii) ischemic stroke outcome (16, 22) (described further in the **Supplementary Material**). The final list of covariates used in the propensity model for each source is shown in Supplementary Table 3.

Patients were stratified according to predicted logit propensity scores, and MMWS weights were calculated to balance baseline covariates between treatment groups, approximating the distribution expected under randomization. Balance was attained with 10 strata using the CMS and combined sources, and with 45 strata using the EHR. Prior to estimating weights, patients outside the region of common support (i.e., interval bounded by the larger of the two treatment-group minimum propensity score logits and the smaller of the two treatment-group maximum propensity score logits) were excluded(21). Weights were calculated to estimate the average treatment effect (ATE). Balance on pre-treatment covariates across and within trials was assessed via absolute standardized mean differences. A weighted Cox regression model was fit, pooling across trials, to estimate the hazard ratio (HR) of incident ischemic stroke between the treated and untreated groups. Weighted Cox models were fit using robust sandwich variance estimators to obtain valid confidence intervals and p-values under IPTW and clustering of sequential trials within patients. The analysis was repeated using the data from the EHR and CMS independently and in combination. Analyses were performed using the Survival package in *R*.

#### Target Trial Emulation Sensitivity Analyses

A series of sensitivity analyses were conducted to explore which specific components of either EHR or CMS data, when removed, produced the largest change in the causal estimates obtained when the EHR and CMS data sources were combined. Six sensitivity analyses were conducted systematically, excluding exposure, outcome, or covariate information from either the EHR or CMS component of the integrated dataset **(Figure 3)**. Results for each sensitivity analysis were compared to the results obtained using the full combined dataset, and results from each data source independently (i.e. using CMS or EHR data alone).

**Figure 3:**
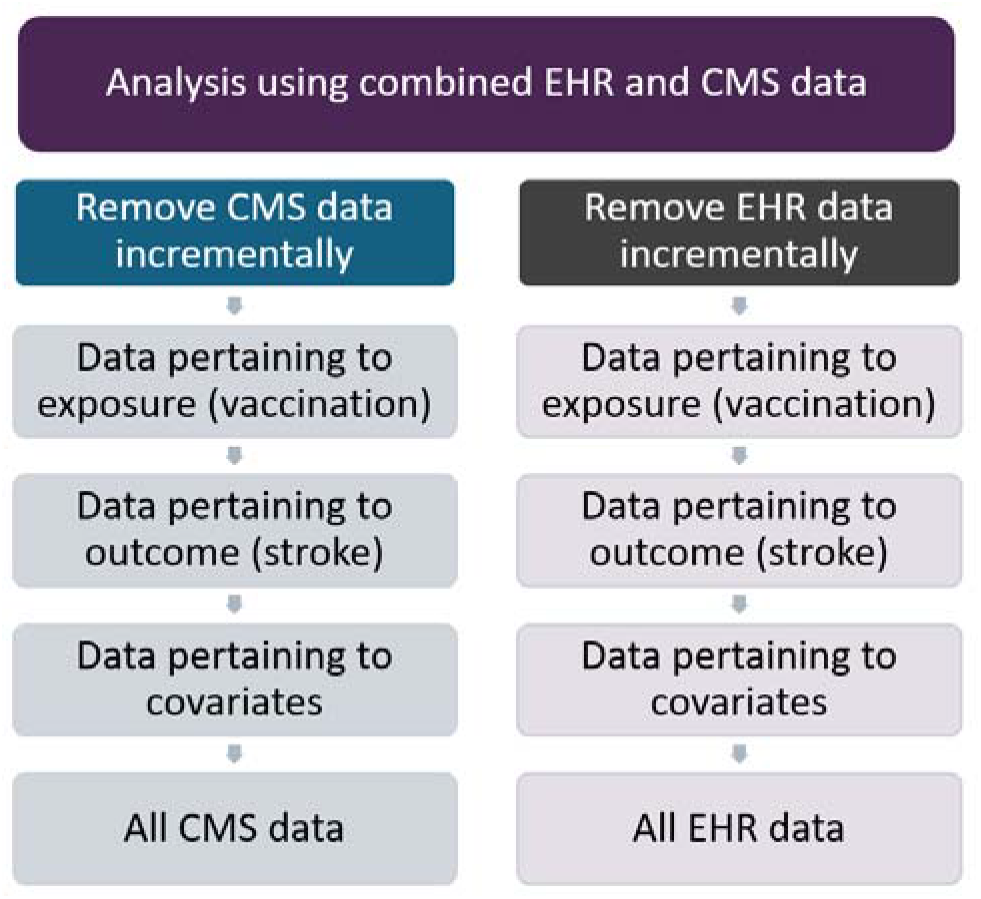
Schematic outlining data omission for sensitivity analysis.

#### Self-Controlled Case Series Study Population

An SCCS analysis was performed during the defined study period of February 2021 to December 2022. Patients were eligible for the SCCS if they: (i) received a COVID-19 vaccination on or after age 65 and during the study period; (ii) had no history of ischemic or hemorrhagic stroke prior to age 65 or study entry; (iii) experienced their first ischemic stroke at age 65 or older during the study period; and (iv) had complete data on date of birth and sex **(Figure 4)**.

**Figure 4:**
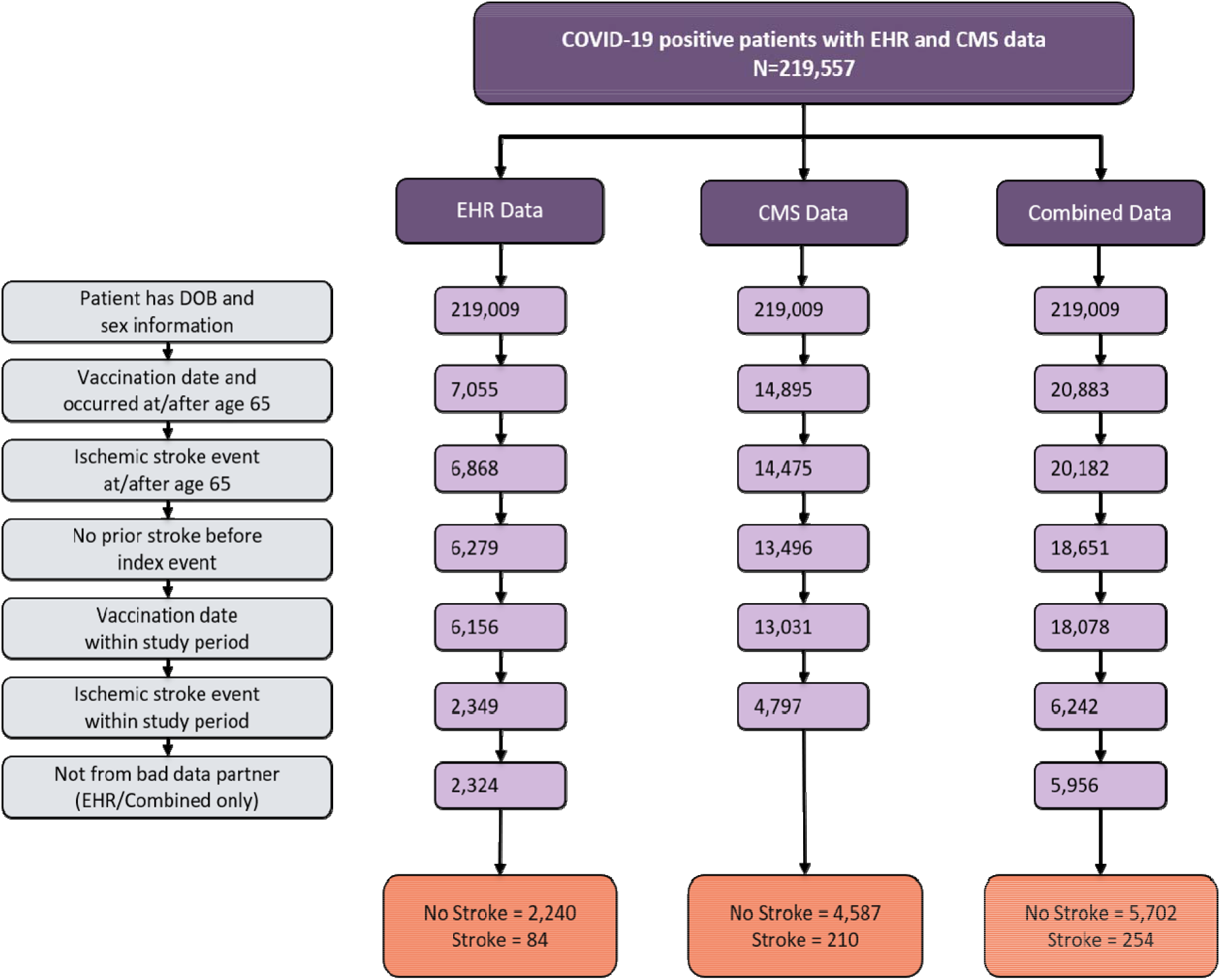
CONSORT diagram showing number of individuals left in each dataset following exclusion criteria for the SCCS study.

#### Self-Controlled Case Series Exposure and Outcome

The primary exposure was patients’ receipt of their first COVID-19 vaccination, and a secondary analysis was done with COVID-19 Diagnosis as the exposure. The outcome was the first incidence of ischemic stroke within 30 days of vaccination. An ischemic stroke codeset was created consisting of numerous ischemic stroke concepts in N3C **(Supplementary Table 1)**. The first healthcare encounter containing one or more ischemic stroke concepts was coded as the incident ischemic stroke event (23).

We defined the observation period for each patient as the latter of February 1, 2021 or when the patient turned 65 years old, and ended on December 21, 2022, the final date available for the CMS data. The observation period was partitioned into three parts: (i) pre-exposure period: 28 days prior to vaccination; (ii) exposure period: day 1 through day 30 following vaccination; and (iii) baseline periods: the remaining time during patients’ observation period, before the pre-exposure period and after the risk period **(Supplementary Figure 3)**. Day 0 was excluded from the risk period since health events occurring on the day of vaccination are vulnerable to reverse causality, and less likely to be credibly due to the vaccination itself.

#### Self-Controlled Case Series Statistical Analysis

The risk of ischemic stroke associated with COVID-19 vaccination was estimated using the standard, exposure-anchored SCCS. The SCCS method compares the incidence of ischemic stroke during risk periods to incidence during baseline periods within the same individual, thereby controlling for fixed patient-level confounders(24). The analysis was entirely conducted on cases who eventually experienced the outcome event (ischemic stroke). Age was included as a time varying covariate binned into 15 quantiles. The relative risk of ischemic stroke in the risk period relative to the baseline period was calculated using conditional Poisson regression and reported as incident rate ratios (IRRs). Analyses were performed using the EHR and CMS data separately and in combination using the SCCS package in *R*.

## Results

### Data Integration

Prior to any descriptive analyses or application of eligibility criteria for the TTE and SCCS, any individual with multiple EHR IDs corresponding to a single CMS ID were deduplicated, and their information was harmonized across EHRs. Records indicated that for one patient, that person’s CMS ID was linked to two EHR IDs which we subsequently deduplicated (see Supplement). Following deduplication, 219,557 unique patients with linked EHR-CMS data were retained.

### Descriptive Analysis of EHR and CMS

We assessed overlap between EHR and CMS data across conditions, medications, procedures, mortality, COVID-19 vaccination, and ischemic stroke before and after COVID-19 diagnosis. Overlap statistics for all variables are shown in **Supplementary Table 2**. Overall, CMS data more completely captured most chronic conditions, both before and after COVID-19 diagnosis. In contrast, medications were more frequently recorded in the EHR data after COVID-19 diagnosis, reflecting greater clinical detail, whereas before diagnosis CMS captured a similar or larger share of patients for several outpatient drug classes (e.g., selective serotonin reuptake inhibitors: CMS 79.9%, EHR 68.4%). After COVID-19 diagnosis, nearly all recorded deaths were captured by CMS (99.4%-99.6% across windows), compared with 64.4%-73.7% by EHR (**Supplementary Figure 4**).

COVID-19 vaccination was captured more completely in CMS than EHR data, with 75.0% and 63.6% of vaccinated individuals identified in the CMS and EHR, respectively, before COVID-19 diagnosis (**Figure 5**). Only 38.6% of vaccinated individuals were captured in both data sources. Ischemic stroke was also more completely captured in CMS than EHR data (90.6% vs. 45.8%), with 36.4% overlap between sources **(Figure 5)**. These findings indicate that reliance on either source alone would result in substantial under-ascertainment of both exposure and outcome.

**Figure 5:**
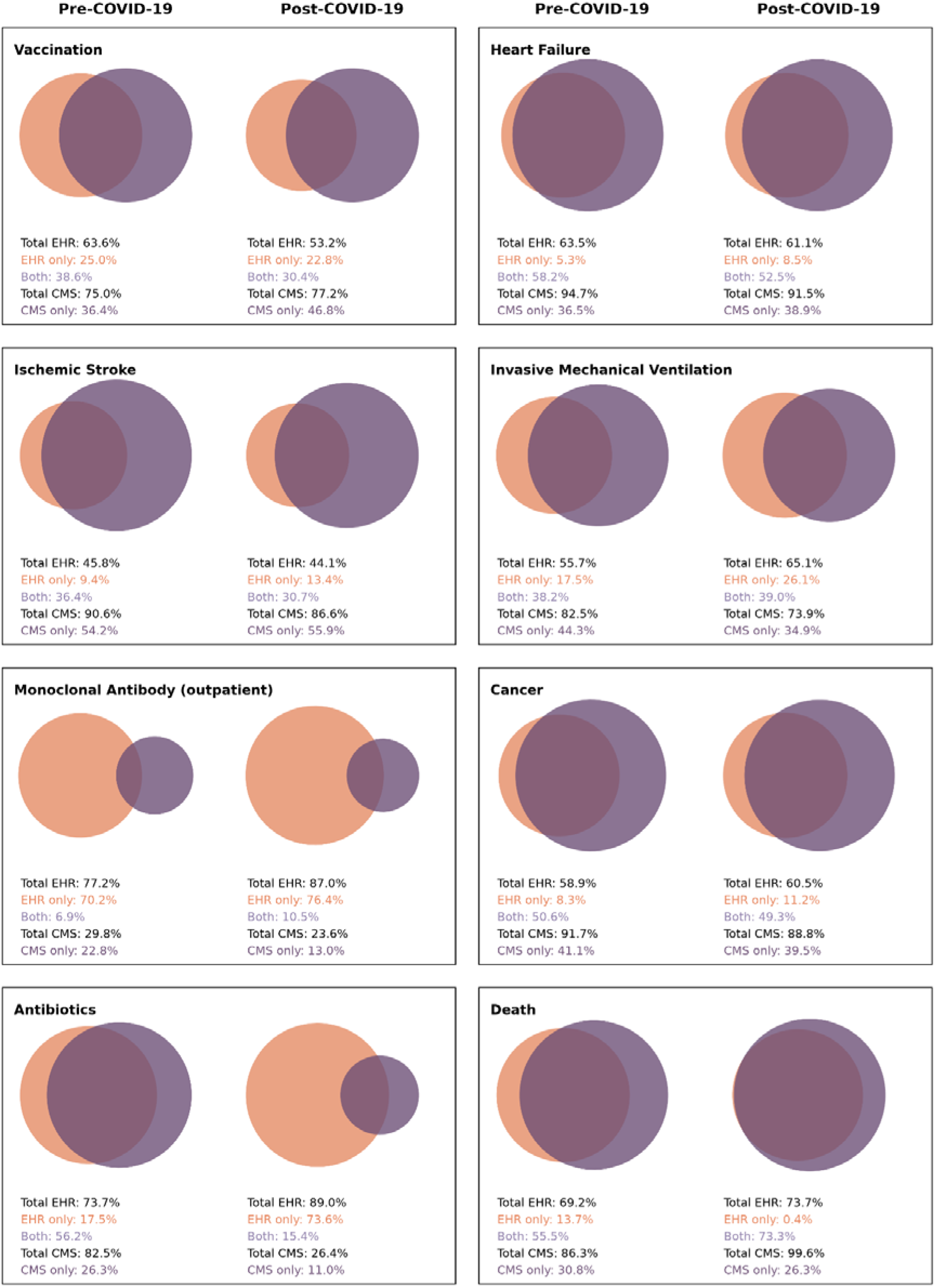
Venn diagrams showing the proportion of patients with data available for key variables pre- and post-COVID-19 diagnosis in the EHR data alone, CMS data alone and the overlap between both sources. *Note.* Monoclonal antibody (outpatient) included the following: tixagevimab/cilgavimab, bebtelovimab, bamlanivimab, etesevimab, sotrovimab, casirivimab, and imdevimab

With respect to comorbidities, prior to COVID-19 diagnosis, CMS data identified a greater proportion of individuals with a history of conditions such as heart failure (94.7%) compared to EHR (63.5%) **(Figure 5)**. For medications pre-COVID-19 diagnosis, EHR captured a considerably higher proportion of patients with outpatient COVID-19-related monoclonal antibody exposure than CMS (77.2% vs. 29.8%), whereas CMS captured a higher proportion of patients with prior antibiotic exposure (82.5% vs. 73.7%). After COVID-19 diagnosis, EHR captured considerably more of both exposures (antibiotics 89.0%, monoclonal antibodies 87.0%), with CMS capturing only 26.4% and 23.6%, respectively. The discrepancies in medication coverage between EHR and CMS highlight the differences in how each data source captures medications: prescriptions for EHR and prescription *fills* for CMS. Procedures, such as invasive ventilation, were better captured in the CMS data both before and immediately after COVID-19 diagnosis **(Figure 5).** EHR captured 55.7% of invasive ventilation cases pre-COVID-19, and this increased to 65.1% post-COVID-19. Comparatively, CMS data captured 82.5% of invasive ventilation cases pre-COVID-19 and 73.9% post-COVID-19.

A more comprehensive set of variables were assessed for overlap at intervals beginning 30, 90, and 180 days after COVID-19 diagnosis. Coverage patterns were broadly similar across the windows beginning at, 30, 90, and 180 days after diagnosis, with the largest shifts occurring between the pre-diagnosis window and the windows after diagnosis **(Supplementary Figure 4; Supplementary Table 2)**. Most conditions were more completely captured by CMS than by EHR data in every window (e.g., hypertension: CMS 96.1% vs. EHR 77.2% before diagnosis and 88.9% vs. 65.4% at 30 days), and overlap between sources declined after diagnosis (hypertension: 73.2% before diagnosis vs. 54.3% at 30 days). Medication exposures showed the most marked change: before diagnosis, CMS captured a similar or larger share of patients for many outpatient drug classes (e.g., antibiotics: CMS 82.5%, EHR 73.7%), whereas after diagnosis EHR captured the large majority (antibiotics immediately after diagnosis: EHR 89.0%, CMS 26.4%), and CMS coverage remained low in the later windows (23.5%-28.0%). Convalescent plasma was the exception, with CMS coverage exceeding EHR coverage before diagnosis and from 30 days onward. Invasive mechanical ventilation was better captured by CMS than by EHR in every window, with the smallest difference immediately after diagnosis (CMS 73.9%, EHR 65.1%). COVID-19 vaccination was captured more completely by CMS in every window (75.0% vs. 63.6% before diagnosis; 75.5%-77.5% vs. 52.9%-53.2% after diagnosis), with overlap of 38.6% before and 28.5%-30.4% after diagnosis. Overlap in mortality increased after COVID-19 diagnosis (55.5% before vs. 73.3% immediately after) and then declined (63.9%-66.5% in the later windows); after diagnosis, CMS captured nearly all recorded deaths (99.4%-99.6%). For nearly all variables, neither source alone captured all patients with a record at any time point, highlighting that a substantial portion of patient information is missed when either data source is used alone.

### Case Study Analysis of EHR and CMS

#### Target Trial Emulation Patient Population

After deduplication and harmonization, eligibility criteria for TTE and SCCS analyses were applied to the linked EHR-CMS dataset. As described, for the TTE, patients could enter up to 12 trials (one for each month) provided they met eligibility criteria for each trial. Each unique combination of patient and trial is herein referred to as a “person-trial”. With 219,557 unique patients available, we had a total of 2,634,684 person-trials before exclusions **(Figure 2).** For the EHR-only analysis, applying the exclusion criteria reduced the cohort to 49,852 individuals resulting in 169,174 person-trials (26,095 vaccinated, 143,079 unvaccinated). For the CMS-only analysis, 88,072 patients remained, yielding 425,245 person-trials (45,092 vaccinated, 380,153 unvaccinated). The combined dataset analysis included 68,480, which led to 244,324 person-trials (43,394 vaccinated, 200,930 unvaccinated). There were fewer patients in the combined versus CMS analysis presumably because the combined analysis applied the BMI and site filters, and because additional data captured by the EHR rendered some CMS patients ineligible.

The characteristics of the eligible cohorts are summarized in **Table 1**. Standardized Mean Differences (SMDs) on all covariates included in the analyses for each data source, before and after weighting, are shown in **Supplementary Table 3**. A higher percentage of vaccinated individuals were found in the EHR data (15.42%, n=26,095 person-trials), compared to CMS data (10.60%, n=45,092 person-trials). The combination of the two data sources resulted in a final sample in which 17.76% (n=43,394) of individuals had received at least one vaccination. Stroke remained uncommon across all data sources, with a crude 30-day cumulative incidence of ≤ 0.64% among unvaccinated and ≤ 0.41% among vaccinated person-trials.

**Table 1:** Characteristics of patients (person-trials) utilized from each data source. Statistics are presented as *n* (%), unless noted by *, which are presented as mean (*SD*). SSRI = Selective Serotonin Reuptake Inhibitor. Percentages are calculated within each vaccination group (i.e., relative to the column N). Prior COVID-19 infection is not shown because person-trials with COVID-19 before the trial month were ineligible.

|  | **EHR**<br>(N=169,174) |  | **CMS**<br>(N=425,245) |  | **Combined**<br>(N=244,324) |  |
| --- | --- | --- | --- | --- | --- | --- |
| **Variable** | **Unvaccinated**<br>**N=143,079** | **Vaccinated**<br>**N=26,095** | **Unvaccinated**<br>**N=380,153** | **Vaccinated**<br>**N=45,092** | **Unvaccinated**<br>**N=200,930** | **Vaccinated**<br>**N=43,394** |
| **Ischemic stroke within 30 days** | 858 (0.60%) | 48 (0.18%) | 1,733 (0.46%) | 183 (0.41%) | 1,283 (0.64%) | 160 (0.37%) |
| **Age\*** | 75.23 (7.32) | 75.27 (7.13) | 75.74 (6.95) | 76.38 (7.22) | 74.87 (7.11) | 75.38 (7.16) |
| **Female** | 82,498 (57.66%) | 14,602 (55.96%) | 211,732 (55.70%) | 25,115 (55.70%) | 114,114 (56.79%) | 24,291 (55.98%) |
| **White, non-Hispanic** | 117,899 (82.40%) | 20,698 (79.32%) | 319,309 (83.99%) | 38,571 (85.54%) | 166,949 (83.09%) | 35,868 (82.66%) |
| **Hispanic or Latino any race** | 3,168 (2.21%) | 846 (3.24%) | 10,247 (2.70%) | 831 (1.84%) | 4,605 (2.29%) | 1,112 (2.56%) |
| **Asian, non-Hispanic** | 1,761 (1.23%) | 723 (2.77%) | 5,913 (1.56%) | 631 (1.40%) | 2,083 (1.04%) | 888 (2.05%) |
| **Black or African American, non-Hispanic** | 16,453 (11.50%) | 2,973 (11.39%) | 31,186 (8.20%) | 3,475 (7.71%) | 21,582 (10.74%) | 4,316 (9.95%) |
| **Antibiotics** | 100,861 (70.49%) | 19,278 (73.88%) | 287,235 (75.56%) | 35,443 (78.60%) | 177,272 (88.23%) | 38,620 (89.00%) |
| **Anticoagulants** | 128,706 (32.59%) | 15,235 (3.86%) | 82,603 (10.50%) | 6,360 (0.81%) | 219,985 (28.61%) | 21,774 (2.83%) |
| **SSRI** | 27,753 (19.40%) | 4,913 (18.83%) | 71,674 (18.85%) | 9,562 (21.21%) | 48,020 (23.90%) | 10,251 (23.62%) |
| **Mental health conditions** | 60,539 (42.31%) | 9,442 (36.18%) | 194,607 (51.19%) | 21,301 (47.24%) | 105,790 (52.65%) | 21,353 (49.21%) |
| **Overweight or obese** | 44,887 (31.37%) | 7,116 (27.27%) | 159,567 (41.97%) | 16,204 (35.94%) | 88,556 (44.07%) | 17,123 (39.46%) |
| **Heart condition** | 65,022 (45.44%) | 10,138 (38.85%) | 206,852 (54.41%) | 22,344 (49.55%) | 110,216 (54.85%) | 21,913 (50.50%) |
| **Kidney disease** | 39,576 (27.66%) | 5,602 (21.47%) | 124,467 (32.74%) | 13,195 (29.26%) | 66,511 (33.10%) | 12,935 (29.81%) |
| **Hypertension** | 109,958 (76.85%) | 18,796 (72.03%) | 324,965 (85.48%) | 36,125 (80.11%) | 171,802 (85.50%) | 35,940 (82.82%) |
| **Cancer** | 38,969 (27.24%) | 6,311 (24.18%) | 156,006 (41.04%) | 16,869 (37.41%) | 76,703 (38.17%) | 16,832 (38.79%) |
| **Diabetes** | 49,703 (34.74%) | 7,960 (30.50%) | 157,688 (41.48%) | 16,397 (36.36%) | 85,032 (42.32%) | 16,624 (38.31%) |

Age was consistent across vaccination status and data source, with a mean of approximately 75 years. The proportion of female patients was similar across vaccination status and data source (55.7%-57.7%). Most patients from each data source were White, followed by Black or African American. Recorded medication use was higher in the combined data than in either source alone. In the EHR data, antibiotic use was 70.49% (n=100,861) and 73.88% (n=19,278) for unvaccinated and vaccinated individuals, respectively. In CMS data, antibiotic use was 75.56% (n=287,235) and 78.60% (n=35,443) for unvaccinated and vaccinated, respectively, and in the combined data it was 88.23% (n=177,272) and 89.00% (n=38,620). Selective Serotonin Reuptake Inhibitor (SSRI) use was similar in EHR and CMS data, with reported use of 19.40% (n=27,753) and 18.83% (n=4,913) for unvaccinated and vaccinated individuals respectively in EHR; and 18.85% (n=71,674) and 21.21% (n=9,562) for unvaccinated and vaccinated individuals respectively in CMS. SSRI use in the combined dataset was 23.90% (n=48,020) and 23.62% (n=10,251) for unvaccinated and vaccinated individuals, respectively.

Comorbidities (e.g., hypertension, mental health condition) were generally more prevalent in CMS than in EHR data among both unvaccinated and vaccinated patients. Among unvaccinated individuals, hypertension was reported at 85.5% (n=324,965) in CMS versus 76.9% (n=109,958) in EHR; diabetes at 41.5% (n=157,688) versus 34.7% (n=49,703), and cancer at 41.0% (n=156,006) versus 27.2% (n=38,969), respectively.

#### Target Trial Emulation Analysis

Using the EHR data alone, the risk of ischemic stroke were 53% lower in vaccinated than unvaccinated patients (HR = 0.47, 95% CI: 0.28-0.79), a statistically significant difference **(Figure 6**, **Table 2)**. Based on the CMS data, there was no statistically significant difference in the risk of ischemic stroke between vaccinated and unvaccinated patients (HR = 1.02, 95% CI: 0.80-1.29). Using the combined data source, there was a non-significant 9% lower risk of stroke in vaccinated versus unvaccinated patients (HR = 0.91, 95% CI: 0.70-1.19).

**Figure 6:**
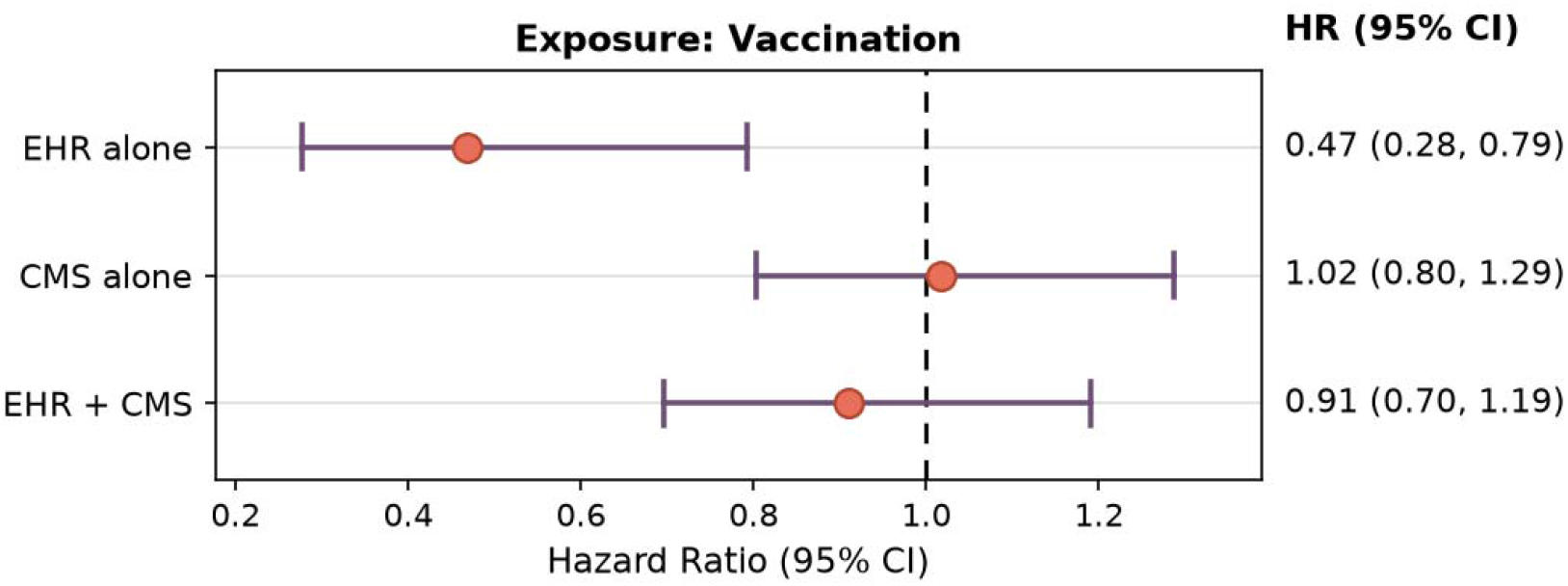
Forest plot of estimated odds ratios with 95% confidence intervals for the association between COVID-19 vaccination and ischemic stroke from the TTE cohort analyses using EHR, CMS, and combined data sources.

**Table 2:**
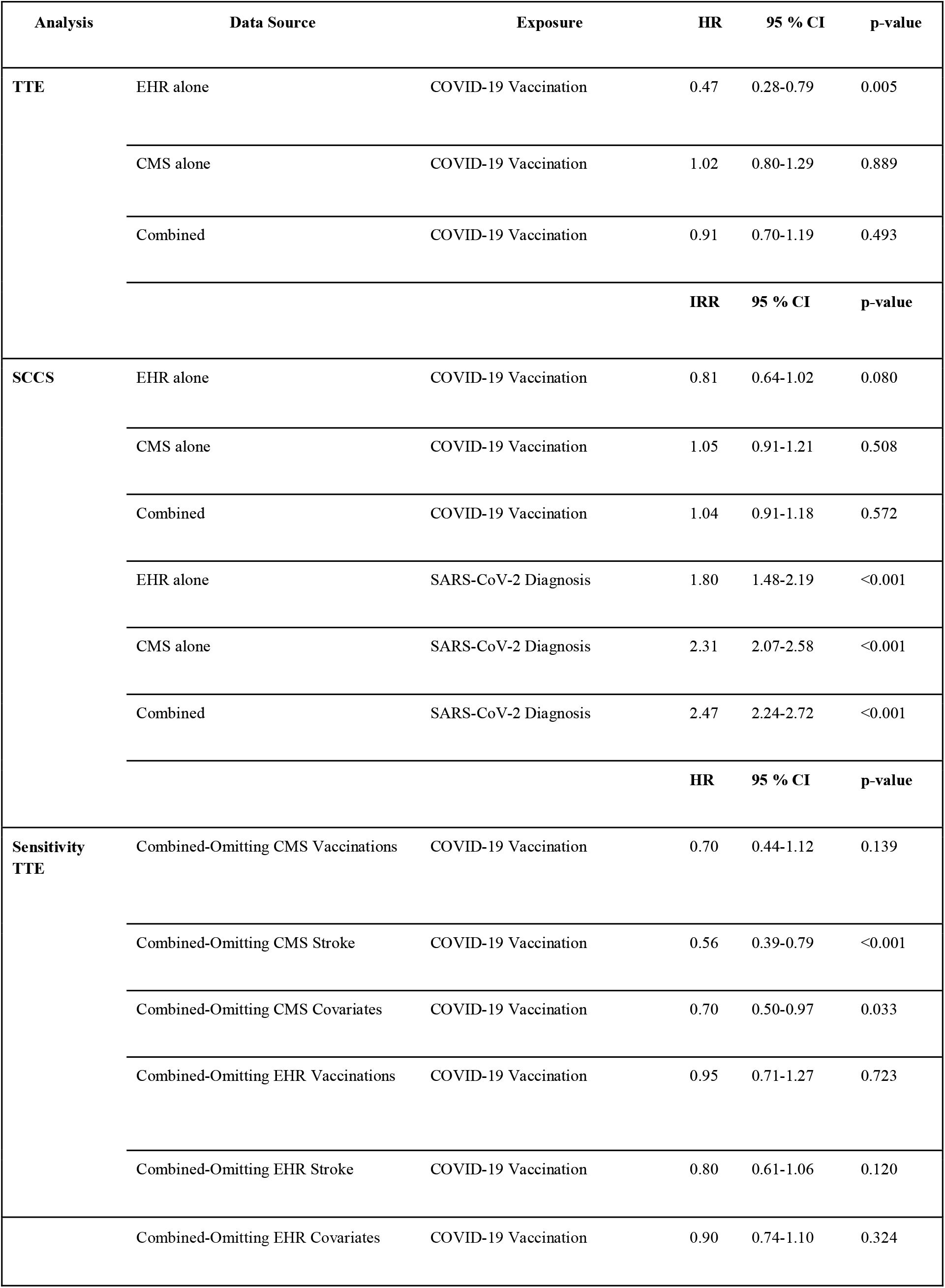
Analysis estimates and confidence intervals. Estimates and confidence intervals of association between vaccination and ischemic stroke for TTE, SCCS, and TTE sensitivity analyses using each data source.

#### Target Trial Emulation Sensitivity Analyses

The sensitivity analysis began with systematically omitting components of the CMS data from the TTE analysis using the full combined data **(Figure 7A)**. When CMS-based vaccination (exposure) was excluded, there was a non-significant 30% decrease in the risk of ischemic stroke in vaccinated patients (HR = 0.70, 95% CI: 0.44-1.12) **(Figure 7B**, **Table 2)**. Removal of CMS-based stroke (outcome) data led to a statistically significant 44% decrease in the risk of ischemic stroke (HR = 0.56, 95% CI: 0.39-0.79). Omitting CMS-based covariate data yielded a statistically significant 30% decrease in the risk of ischemic stroke (HR = 0.70, 95% CI: 0.50-0.97), in contrast to the non-significant result of the main analysis where EHR and CMS data were combined. Lastly, omitting all CMS data (i.e. relying on EHR data alone) resulted in a statistically significant HR of 0.47 (95% CI: 0.28-0.79), as noted previously.

**Figure 7:**
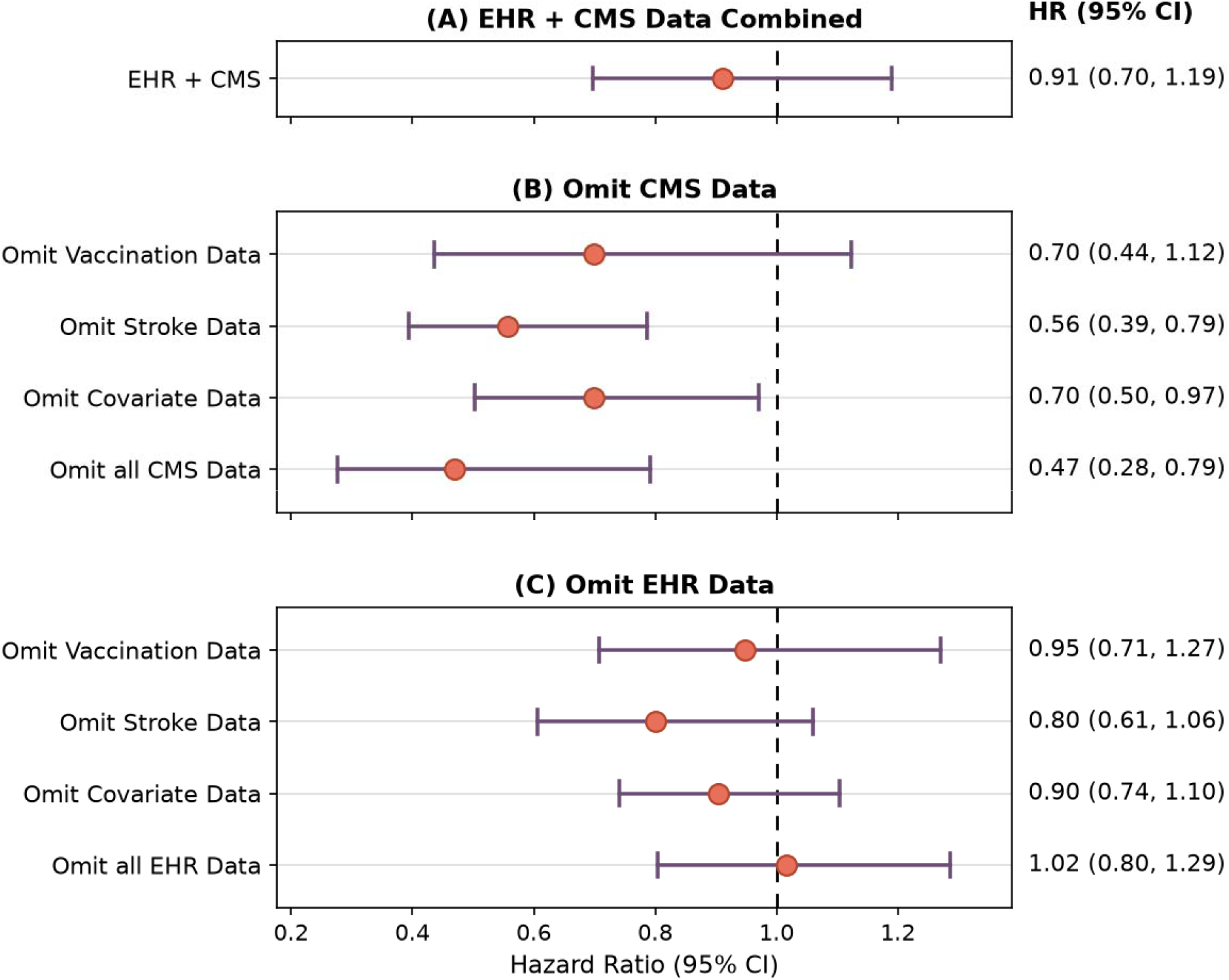
Forest plots depicting the estimated odds ratios and corresponding 95% confidence intervals for the association between vaccination and ischemic stroke across sensitivity analyses. (A) Results from the combined EHR and CMS data. (B) Results from the sensitivity analyses omitting CMS data on vaccination, stroke, or covariate data from the combined dataset. The omission of all CMS data is synonymous with the main EHR-only analysis. (C) Results from the sensitivity analyses of omitting EHR vaccination, stroke, or covariate data from the combined dataset. The omission of all EHR data is synonymous with the main CMS-only analysis.

When EHR-based vaccination (exposure; HR = 0.95, 95% CI: 0.71-1.27), stroke (outcome; HR = 0.80, 95% CI: 0.61-1.06) or covariate data (HR = 0.90, 95% CI: 0.74-1.10) were individually omitted, no significant association between vaccination and ischemic stroke emerged, consistent with the results from the main analysis in which EHR and CMS data were combined **(Figure 7C**, **Table 2)**. Similarly, omitting all EHR data (relying on CMS data alone) resulted in no significant association between vaccination and ischemic stroke (HR = 1.02, 95% CI: 0.80-1.29), as noted previously.

#### Self-Controlled Case Series Patient Population

The same 219,557 patients with linked EHR and CMS data used for the TTE study were used to generate the SCCS study sample. Individuals were required to have received a COVID-19 vaccination and experienced an ischemic stroke on or after the age of 65 during the study period. In the EHR only analysis, application of the exclusion criteria yielded 2,324 eligible patients, of whom 84 experienced an ischemic stroke within 30 days of vaccination and 2,240 did not **(Figure 4).** CMS data contained 4,797 individuals, 210 with stroke within 30 days and 4,587 without. The combined data resulted in 5,956 individuals, 254 with stroke within 30 days and 5,702 without.

Approximately 96% (EHR n=2,240 (96.39%), CMS n=4,587 (95.62%), combined n=5,702 (95.74%)) of individuals in each of the three SCCS cohorts did not experience the outcome of interest (ischemic stroke) within 30 days of COVID-19 vaccination, while about 4% (EHR n=84 (3.61%), CMS n=210 (4.38%), combined n=254 (4.26%)) did **(Table 3)**. Sex distribution was consistent across data sources, with approximately 51%-53% female (EHR n=1,138 (50.80%), CMS n=2,413 (52.61%), combined n=3,003 (52.67%)) among patients without ischemic stroke and 54%-57% female (EHR n=45 (53.57%), CMS n=119 (56.67%), combined n=145 (57.09%)) among patients with ischemic stroke. COVID-19 diagnosis among no-stroke and stroke patients, respectively, in each data source was as follows: 64.33% (n=1,441) and 58.33% (n=49) in EHR data; 80.79% (n=3,706) and 77.62% (n=163) in CMS data, and 81.16% (n=4,628) and 76.77% (n=195) in the combined data. Mortality capture among ischemic stroke non-cases and cases, respectively, in the combined dataset was 20.89% (n=1,191) and 27.95% (n=71), compared to 16.07% (n=360) and a suppressed value (<20 patients) in the EHR data and 19.86% (n=911) and 25.24% (n=53) in the CMS data. Age at baseline averaged approximately 77 years (EHR: 76.82 (7.56), 77.37 (7.34)), (CMS: 77.25 (7.91), 76.54 (8.54)), (combined: 76.98 (7.75), 77.27 (8.16)) for no stroke and stroke, respectively, regardless of data source or outcome status.

**Table 3.** SCCS cohort statistics by data source. Patient characteristics of SCCS cohort. Statistics are presented as n (%) unless noted by *, which are presented as mean (*SD*). Cells with fewer than 20 patients are suppressed (<20) in accordance with N3C minimum cell size requirements.

|  | **EHR**<br>(N=2,324) |  | **CMS**<br>(N=4,797) |  | **Combined**<br>(N=5,956) |  |
| --- | --- | --- | --- | --- | --- | --- |
|  | **No stroke**<br>**within 30 days** | **Stroke within**<br>**30 days** | **No stroke**<br>**within 30 days** | **Stroke within**<br>**30 days** | **No stroke**<br>**within 30 days** | **Stroke within**<br>**30 days** |
| **n** | 2,240 (96.39%) | 84 (3.61%) | 4,587 (95.62%) | 210 (4.38%) | 5,702 (95.74%) | 254 (4.26%) |
| **Female** | 1,138 (50.80%) | 45 (53.57%) | 2,413 (52.61%) | 119 (56.67%) | 3,003 (52.67%) | 145 (57.09%) |
| **COVID-19 Diagnosis** | 1,441 (64.33%) | 49 (58.33%) | 3,706 (80.79%) | 163 (77.62%) | 4,628 (81.16%) | 195 (76.77%) |
| **Mortality** | 360 (16.07%) | <20 | 911 (19.86%) | 53 (25.24%) | 1,191 (20.89%) | 71 (27.95%) |
| **Total Visit Prior to**<br>**Baseline\*** | 63.64 (77.31) | 53.57 (54.85) | 217.60 (186.37) | 210.43 (170.53) | 225.85 (186.35) | 223.79 (165.29) |
| **Age at Baseline\*** | 76.82 (7.56) | 77.37 (7.34) | 77.25 (7.91) | 76.54 (8.54) | 76.98 (7.75) | 77.27 (8.16) |

#### Self-Controlled Case Series Analysis

For the SCCS, no statistically significant association between vaccination and ischemic stroke was observed in any data source. The point estimate using EHR data alone was below the null (IRR = 0.81, 95% CI: 0.64-1.02) **(Figure 8A**, **Table 2)**, whereas the CMS data alone and the combined data yielded estimates close to the null (CMS alone: IRR = 1.05, 95% CI: 0.91-1.21; Combined data: IRR = 1.04, 95% CI: 0.91-1.18).

**Figure 8:**
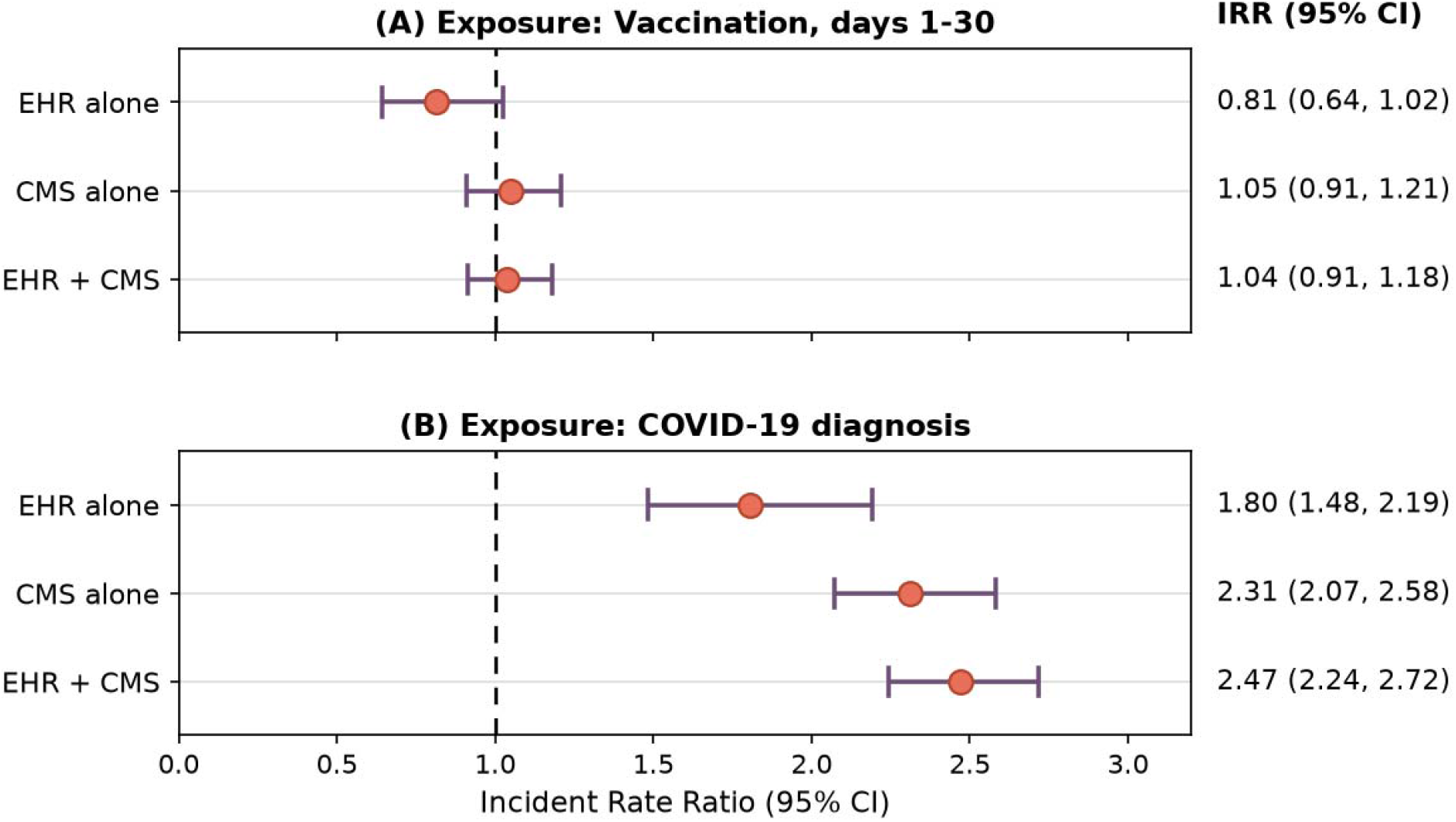
Forest plots of the estimated incidence rate ratios with 95% confidence intervals from the SCCS analyses using EHR, CMS, and combined data sources. Two sets of analyses were done: (A) Association between COVID-19 vaccination and ischemic stroke, and (B) Association between COVID-19 diagnosis and ischemic stroke.

When examining COVID-19 diagnosis in relation to ischemic stroke, irrespective of data source, results demonstrated a statistically significant, approximately 1.8- to 2.5-fold increased incidence of ischemic stroke **(Figure 8B**, **Table 2)**. The IRR was 1.80 (95% CI: 1.48-2.19) for EHR data alone, 2.31 (95% CI: 2.07-2.58) for CMS data alone, and 2.47 (95% CI: 2.24-2.72) using the combined dataset.

## Discussion

This study demonstrates the critical role of integrating EHR and CMS data and provides compelling evidence that multi-modal data-integration advances methodological rigor and clinical insights. We have shown how the combination of both data sources, compared to each data source alone, provides a more complete, accurate, and reliable representation of a patient’s medical history across multiple healthcare institutions. Obtaining a comprehensive picture of patient health journeys contributes to increased validity of causal inferences and reduces the risk of bias when conducting real-world studies of public health importance, particularly those relying on retrospective causal inference frameworks.

### Overlap and Complementarity of EHR and CMS

Once integrated, notable differences were observed between CMS and EHR data, the extent to which data was captured between the two sources. We examined overlap in data domains relevant to exposure, outcome, and covariates (i.e. diseases/conditions, medications, and mortality). With the understanding that visits to medical institutions may be influenced by a major medical event like COVID-19 diagnosis and hospitalization (25), we assessed these key variables pre- and post-diagnosis.

The comparison of diseases and conditions captured across EHR and CMS data over time has been understudied. Previous research has focused mainly on cross-sectional examination of medications or other limited domains of medical information overlap between EHRs and claims (1, 3, 4, 12). We found that diagnoses and conditions, such as heart failure, ischemic stroke, and cancers showed considerable overlap between EHR and CMS data sources but were more completely captured by CMS both before and after COVID-19 diagnosis, and data source overlap slightly declines from before to after COVID-19 diagnosis. Failing to include claims data could lead to significant amounts of misclassification of individuals in relation to diseases and conditions (whether as covariates or outcomes), possibly jeopardizing scientific results.

The coverage and overlap of data contained in EHR and claims data has not been thoroughly assessed for medical events that happen exclusively during a healthcare facility visit. Such events include invasive mechanical ventilation (IMV), transplant, extracorporeal membrane oxygenation (ECMO), and monoclonal antibodies, which are or the most part administered in a clinical setting. Monoclonal antibodies were captured better by EHR data before and after COVID-19 diagnosis. Interestingly, invasive mechanical ventilation was captured better by CMS data both before and immediately after diagnosis, whereas EHR better captured transplant across all periods overall, and better captured ECMO post-COVID-19 diagnosis (**Supplementary Table 2**). Prior research suggested that EHR data may show more coverage of in-facility care variables than CMS due to bundling practices often used in claims for billing purposes, which often leads to details of specific inpatient procedures and medications being lost or under-represented in claims data (1, 26). Our results are generally in line with this perspective but nonetheless show that there is variability with respect to specific inpatient procedures (e.g., IMV, ECMO). More research into data source coverage and overlap for in-facility procedures and medications may help in discovering clearer trends. Interestingly however, antibiotics, which are not exclusively administered inpatient, showed a similar pattern in coverage as COVID-19-related monoclonal antibody medications after diagnosis (but not before, when CMS captured more antibiotic exposures), albeit with a higher degree of overlap both before (56.2% vs. 6.9%) and after (15.4% vs. 10.5%) COVID-19 diagnosis.

Medication information poses a particular challenge because EHR and claims data differ in how they capture medication exposure and in the reliability of what each record implies. EHR medication records generally represent prescriptions rather than fills, so researchers using EHR data alone often cannot determine whether a prescription was filled or whether the medication was actually taken; this limitation is reflected in primary non-adherence, which has been reported to range from 5% to 55% (11, 12) depending on medication type. EHR data may also miss medications obtained outside the contributing healthcare system. In contrast, CMS data capture prescription *fills*, providing a better, though still approximate, indicator of medication adherence (1); however, CMS data do not capture medications paid for out of pocket, by other insurance, or through free services. Consistent with these differences, our findings indicated that medication data were more comprehensively captured in the EHR after landmark diagnosis, whereas before diagnosis CMS coverage of several outpatient drug classes was similar to or higher than EHR coverage and then decreased dramatically after COVID-19 diagnosis. The overlap between EHR and CMS records can increase confidence in patients’ actual medication exposure, and CMS-exclusive fills can add more reliable information pertaining to medication exposure, even if limited in volume. Ultimately, neither data source can prove that a patient actually took a medication or establish the treatment intent, especially when medications may be used off-label (27, 28), implying that even when EHR and claims data are combined, they don’t provide a complete representations of a patient’s drug exposure history.

These medication-specific limitations foreshadow a broader challenge: linking claims and EHR data can provide a fuller—but still incomplete—view of patients’ healthcare history and utilization. EHRs may capture provider-generated clinical information not tied to billing, while CMS claims can capture care outside a patient’s primary health system. However, both sources remain rooted in healthcare encounters and therefore offer only limited snapshots of the health experiences that led people to seek care. Even combined, they may miss daily functioning, broader well-being, and factors shaping health outside clinical settings. Furthermore, the primary (and *non-*research) purpose of each data source places limits on the utility of each source in fully and accurately capturing patients’ healthcare histories, particularly for claims. Namely, the way in which diagnosis and procedure codes are recorded are directly tied to a healthcare institution’s coding practices for reimbursement and billing. Some evidence indicates institutions may adopt unreliable coding practices (29). Even more alarming, coding may be inflated or coded more aggressively to enhance reimbursement via diagnosis-upcoding or risk score inflation (30, 31). Both aspects compromise the validity of the information captured in the claims data. EHR data can help fill gaps or verify claims-based codes through provider-created information such as problem lists, but the combined data still do not fully capture patients’ health history or functioning because they were created for operational purposes rather than comprehensive documentation of patients’ lives, health, and functioning.

### Implications for Causal Inference

Vaccine safety, particularly in relation to adverse effects of COVID-19 vaccination, has been investigated using routinely collected data by numerous researchers(14, 32–34). Importantly, studies that have relied on EHR or claims data alone have yielded conflicting findings as to whether or not COVID-19 increases the risk of stroke(2, 35). Such contrasting results could be due to differential capture of patient medical data needed to adequately address the research question between EHR and claims. Studies indicate that vaccine records are better captured by EHRs (36, 37), although in the current study, CMS captured a larger share of recorded COVID-19 vaccinations than EHR (75.0% vs. 63.6% before diagnosis; **Figure 5**). Patient outcomes (such as ischemic stroke) and comorbidities are generally often better captured by claims(1), which is supported by the current findings.

We compared EHR data, CMS data, and their combination across two causal inference study designs: a TTE cohort study and SCCS, to assess similarities and differences in observed associations between COVID-19 vaccination and ischemic stroke. Using either data source individually may introduce biases due to misclassification or missing data, potentially contributing to inconsistent results across different causal inference methods. Indeed, the analyses that used EHR data alone yielded results that differed in statistical significance across the TTE and SCCS and departed from the combined data estimates, whereas analyses using CMS data alone yielded estimates close to the null in both designs.

For example, EHR data produced a statistically significant negative association using TTE (HR=0.47; 95% CI 0.28 to 0.79) and a smaller, non-significant negative association using SCCS (IRR=0.81; 95% CI 0.64 to 1.02). In contrast, results from the analysis of CMS data showed no association between vaccination and stroke in either design (TTE HR=1.02; 95% CI 0.80 to 1.29; SCCS IRR=1.05; 95% CI 0.91 to 1.21). The EHR analyses may have incorrectly included patients who had a prior stroke or who had some other serious illness, which was not fully captured in the EHRs (i.e., ineligible patients). This may have produced a *healthy vaccinee effect* where patients who had recently suffered a stroke or serious illness delayed being vaccinated to allow recovery, which would cause a rapid decline in severe health events closer to vaccination day (38–40). Essentially, this would mean that the pool of patients who did get vaccinated were selectively healthier than average, leading to the observed negative association between vaccination and stroke in the EHR analysis. The CMS (and by extension, the combined) analysis may have been better able to adjust for this effect via improved capture of (i) stroke, (ii) covariate, and/or (iii) vaccination data, which is supported by our descriptive analysis (**Figure 5, Supplementary Figure 4**). Indeed, the sensitivity analyses revealed that omitting CMS-related outcome (i.e. stroke) and covariate data from the combined analyses shifted the combined result towards the significant, negative association observed in the EHR-only result. This would imply that integrating CMS data ensured more critical health information about patients was captured, allowing for improved adjustment of patient characteristics that give rise to the healthy vaccinee bias, and yielding a non-significant vaccination-stroke association consistent with prior research(32, 41).

The significant negative association between vaccination and stroke did not emerge in the SCCS for the EHR (or other data sources) most likely for two reasons. First, patients acted as their own control, thereby eliminating the need for a comprehensive set of time-invariant covariates to adjust for selection bias. Hence, the increased capture of health conditions attributable to CMS data would not have contributed any meaningful reduction in bias when using EHR alone. Second, the SCCS analysis explicitly included a pre-exposure risk period which is used to adjust for the rapid decline of adverse events close to the vaccination that may contribute to such bias (also known as the *event-dependent exposure* bias (42)). Consequently, we reported a non-significant association between vaccination and stroke which was consistent across data sources and prior research (17, 35, 43, 44).

Our findings imply that using EHR data alone may leave gaps in exposure, outcome, covariate, and eligibility information, introducing selection, healthy vaccinee, and misclassification biases that threaten the validity of causal inferences. However, given that the CMS data alone yielded consistent results across causal inference methods, comparable to the combined data, one may ask whether integrating CMS with EHR data offers any benefit. We believe it does. First, consistency of results should not be mistaken for adequacy of data: only through linkage could we show that the null association persisted despite richer patient characterization, and that the divergent EHR-only estimate likely reflected source-specific bias. Second, a substantial share of comorbidity and medication records were found only in the EHR data (**Figure 5**), which also uniquely provided clinical measurements (e.g., body mass index) and site of care; the latter was markedly imbalanced by vaccination status (**Supplementary Table 3**) and may capture differences in healthcare access and care-seeking behavior that the CMS-only analyses could not address (45, 46). Third, ischemic stroke almost invariably generates a claim; outcomes requiring laboratory, disease severity, or lifestyle data are likely to benefit more from linkage. Finally, EHR data typically become available sooner than claims, enabling more timely safety surveillance (47, 48). Thus, the value of linkage may lie less in altering effect estimates than in strengthening the credibility and rigor of real-world evidence.

### Limitations

Our study has several limitations. First, the CMS data we used were limited to Medicare beneficiaries, restricting our analysis to patients aged 65 and older. As a result, the added value of combining EHR and CMS may not generalize to patients across all age groups or all insurance types. Additionally, CMS data were only available for a subset of older adults in N3C, limiting our conclusions to patients with linked CMS data. However, the improvements in data quality and validity observed here could extend to broader populations if additional CMS sources were included, such as Medicaid or private insurance claims. Second, while we observed that using each data source alone produced inconsistent results, we did not formally quantify the extent to which these limitations in each source contributed to this inconsistency. Our sensitivity analysis provided insight into data source contributions, but no corrections for multiple comparisons were applied, which may limit the strength of our results. Third, vaccinations given in settings without claims or EHR documentation (i.e. mass vaccination clinics) may have led to exposure misclassification and biased effect estimates. Fourth, in the TTE and SCCS analyses, the effect measures were the HR and IRR, respectively. Although both are measurements of the vaccination-stroke association, they are not identical quantities. The HR quantifies the relative risk of ischemic stroke between the treated versus untreated cohorts, whereas the IRR quantifies the relative incidence of ischemic stroke between risk and baseline periods. Thus, these causal inference approaches are not directly comparable, but are nonetheless complementary, and arguably should yield consistent results. Finally, the benefit of linking EHR and CMS data may vary depending on the specific research question and provide more value in scenarios where more complete data capture pertaining to exposures and outcomes is required.

### Conclusions

This study demonstrates that relying on a single data source can result in incomplete capture or misclassification of patients’ exposures, outcomes, and covariates. In our case study of COVID-19 vaccination and ischemic stroke, analyses using EHR data alone departed from the combined analysis, likely reflecting incomplete capture of strokes and comorbidities occurring outside participating health systems, whereas analyses using CMS data alone yielded estimates consistent with the combined data and with prior research. Nevertheless, integrating EHR and CMS data provided benefits that neither source offered alone: it enabled more accurate identification of eligible patients, captured exposures and outcomes occurring across care settings, contributed clinical information available only in the EHR (e.g., body mass index, site of care) for confounding adjustment, and, critically, allowed cross-source comparisons that helped identify likely bias in the EHR-only estimate and confirmed the robustness of the null finding. The added value of linkage is likely to be greater still for outcomes and confounders that are poorly captured in claims. Ultimately, leveraging both clinical and claims data is essential for producing more complete, credible, and actionable scientific evidence to guide public health and clinical decision-making.

## Data Availability

The datasets generated and/or analyzed during the current study are available in the N3C Data Enclave at http://unite.nih.gov. The N3C enclave is available for public use. To access data used within this manuscript, institutions must have a signed Data Use Agreement executed with the U.S. National Center for Advancing Translational Sciences (NCATS) and their investigators must complete mandatory training and must submit a Data Use Request (DUR) to N3C. To request N3C data access, researchers must follow instructions at https://covid.cd2h.org/onboarding. Code is available to those with valid login credentials for the N3C Data Enclave, which can be accessed at: https://www.palantir.com/platforms/foundry/. It was written for use in the enclave on the Palantir Foundry platform, where the analysis can be reproduced by researchers. It can be exported for review upon request.

## Declarations

National COVID Cohort Collaborative attribution.

The analyses described in this publication were conducted with data or tools accessed through the NCATS N3C Data Enclave https://covid.cd2h.org and N3C Attribution & Publication Policy v1.2-2020-08-25b supported by NCATS U24 TR002306, and Axle Informatics Subcontract: NCATS-P00438-B. This research was possible because of the patients whose information is included within the data and the organizations (https://ncats.nih.gov/n3c/resources/data-contribution/data-transfer-agreement-signatories) and scientists who have contributed to the on-going development of this community resource (13). Enclave data is protected, and can be accessed for COVID-related research by those with an approved protocol and data use request from an institutional review board. Data access is governed under the authority of the National Institutes of Health; more information on accessing the data can be found at https://covid.cd2h.org/for-researchers. See Haendel et. al. (18) for details on how data is ingested, managed, and protected within the N3C Data Enclave.

### Ethics approval and consent to participate

This study was performed in line with the principles of the Declaration of Helsinki. Use of N3C data for this study does not involve human subjects (45 CFR 46.102) as determined by the NIH Office of IRB Operations. Thus, this study did not include factors necessitating patient consent.

The N3C data transfer to the National Centre for Advancing Translational Sciences (NCATS) is performed under a Johns Hopkins University Reliance Protocol #IRB00249128 or individual site agreements with the National Institutes of Health (NIH). The N3C Data Enclave is managed under the authority of the NIH; information can be found at https://ncats.nih.goc/n3c/resources. The content is solely the responsibility of the authors and does not necessarily represent the official views of the National Institutes of Health or the N3C program.

### Consent for publication

Not applicable.

Availability of data and materials

### Competing Interests

The authors declare that they have no competing interests.

### Funding

This research was supported in part by the Extramural Research Program of the National Center for Advancing Translational Sciences, National Institutes of Health. The content of this publication does not necessarily reflect the views or policies of the Department of Health and Human Services, nor does mention of trade names, commercial products, or organizations imply endorsement by the U.S. Government.

### Authors’ contributions

Authorship was determined using ICMJE recommendations.

⍰ Conception: SM, HS, KG
⍰ Design: SM, TT, HS
⍰ Analysis: SM, DB, TE
⍰ Acquisition of data: NH, KH, KG
⍰ Manuscript drafting: SM, TT, DB, TE, AK
⍰ Critical revision of manuscript: SM, DB, TT, KH, NH, AK, HS, KG

## Acknowledgements

We would like to thank Michele Jonsson Funk for valuable discussions and feedback.

## We gratefully acknowledge the following core contributors to N3C

Adam B. Wilcox, Adam M. Lee, Alexis Graves, Alfred (Jerrod) Anzalone, Amin Manna, Amit Saha, Amy Olex, Andrea Zhou, Andrew E. Williams, Andrew Southerland, Andrew T. Girvin, Anita Walden, Anjali A. Sharathkumar, Benjamin Amor, Benjamin Bates, Brian Hendricks, Brijesh Patel, Caleb Alexander, Carolyn Bramante, Cavin Ward-Caviness, Charisse Madlock-Brown, Christine Suver, Christopher Chute, Christopher Dillon, Chunlei Wu, Clare Schmitt, Cliff Takemoto, Dan Housman, Davera Gabriel, David A. Eichmann, Diego Mazzotti, Don Brown, Eilis Boudreau, Elaine Hill, Elizabeth Zampino, Emily Carlson Marti, Emily R. Pfaff, Evan French, Farrukh M Koraishy, Federico Mariona, Fred Prior, George Sokos, Greg Martin, Harold Lehmann, Heidi Spratt, Hemalkumar Mehta, Hongfang Liu, Hythem Sidky, J.W. Awori Hayanga, Jami Pincavitch, Jaylyn Clark, Jeremy Richard Harper, Jessica Islam, Jin Ge, Joel Gagnier, Joel H. Saltz, Joel Saltz, Johanna Loomba, John Buse, Jomol Mathew, Joni L. Rutter, Julie A. McMurry, Justin Guinney, Justin Starren, Karen Crowley, Katie Rebecca Bradwell, Kellie M. Walters, Ken Wilkins, Kenneth R. Gersing, Kenrick Dwain Cato, Kimberly Murray, Kristin Kostka, Lavance Northington, Lee Allan Pyles, Leonie Misquitta, Lesley Cottrell, Lili Portilla, Mariam Deacy, Mark M. Bissell, Marshall Clark, Mary Emmett, Mary Morrison Saltz, Matvey B. Palchuk, Melissa A. Haendel, Meredith Adams, Meredith Temple-O’Connor, Michael G. Kurilla, Michele Morris, Nabeel Qureshi, Nasia Safdar, Nicole Garbarini, Noha Sharafeldin, Ofer Sadan, Patricia A. Francis, Penny Wung Burgoon, Peter Robinson, Philip R.O. Payne, Rafael Fuentes, Randeep Jawa, Rebecca Erwin-Cohen, Rena Patel, Richard A. Moffitt, Richard L. Zhu, Rishi Kamaleswaran, Robert Hurley, Robert T. Miller, Saiju Pyarajan, Sam G. Michael, Samuel Bozzette, Sandeep Mallipattu, Satyanarayana Vedula, Scott Chapman, Shawn T. O’Neil, Soko Setoguchi, Stephanie S. Hong, Steve Johnson, Tellen D. Bennett, Tiffany Callahan, Umit Topaloglu, Usman Sheikh, Valery Gordon, Vignesh Subbian, Warren A. Kibbe, Wenndy Hernandez, Will Beasley, Will Cooper, William Hillegass, Xiaohan Tanner Zhang. Details of contributions available at covid.cd2h.org/core-contributors

## Data Partners with Released Data

The following institutions whose data is released or pending:

Available: Advocate Health Care Network — UL1TR002389: The Institute for Translational Medicine (ITM) • Boston University Medical Campus — UL1TR001430: Boston University Clinical and Translational Science Institute • Brown University — U54GM115677: Advance Clinical Translational Research (Advance-CTR) • Carilion Clinic — UL1TR003015: iTHRIV Integrated Translational health Research Institute of Virginia • Charleston Area Medical Center — U54GM104942: West Virginia Clinical and Translational Science Institute (WVCTSI) • Children’s Hospital Colorado — UL1TR002535: Colorado Clinical and Translational Sciences Institute • Columbia University Irving Medical Center — UL1TR001873: Irving Institute for Clinical and Translational Research • Duke University — UL1TR002553: Duke Clinical and Translational Science Institute • George Washington Children’s Research Institute — UL1TR001876: Clinical and Translational Science Institute at Children’s National (CTSA-CN) • George Washington University — UL1TR001876: Clinical and Translational Science Institute at Children’s National (CTSA-CN) • Indiana University School of Medicine — UL1TR002529: Indiana Clinical and Translational Science Institute • Johns Hopkins University — UL1TR003098: Johns Hopkins Institute for Clinical and Translational Research • Loyola Medicine — Loyola University Medical Center • Loyola University Medical Center — UL1TR002389: The Institute for Translational Medicine (ITM) • Maine Medical Center — U54GM115516: Northern New England Clinical & Translational Research (NNE-CTR) Network • Massachusetts General Brigham — UL1TR002541: Harvard Catalyst • Mayo Clinic Rochester — UL1TR002377: Mayo Clinic Center for Clinical and Translational Science (CCaTS) • Medical University of South Carolina — UL1TR001450: South Carolina Clinical & Translational Research Institute (SCTR) • Montefiore Medical Center — UL1TR002556: Institute for Clinical and Translational Research at Einstein and Montefiore • Nemours — U54GM104941: Delaware CTR ACCEL Program • NorthShore University HealthSystem — UL1TR002389: The Institute for Translational Medicine (ITM) • Northwestern University at Chicago — UL1TR001422: Northwestern University Clinical and Translational Science Institute (NUCATS) • OCHIN — INV-018455: Bill and Melinda Gates Foundation grant to Sage Bionetworks • Oregon Health & Science University — UL1TR002369: Oregon Clinical and Translational Research Institute • Penn State Health Milton S. Hershey Medical Center — UL1TR002014: Penn State Clinical and Translational Science Institute • Rush University Medical Center — UL1TR002389: The Institute for Translational Medicine (ITM) • Rutgers, The State University of New Jersey — UL1TR003017: New Jersey Alliance for Clinical and Translational Science • Stony Brook University — U24TR002306 • The Ohio State University — UL1TR002733: Center for Clinical and Translational Science • The State University of New York at Buffalo — UL1TR001412: Clinical and Translational Science Institute • The University of Chicago — UL1TR002389: The Institute for Translational Medicine (ITM) • The University of Iowa — UL1TR002537: Institute for Clinical and Translational Science • The University of Miami Leonard M. Miller School of Medicine — UL1TR002736: University of Miami Clinical and Translational Science Institute • The University of Michigan at Ann Arbor — UL1TR002240: Michigan Institute for Clinical and Health Research • The University of Texas Health Science Center at Houston — UL1TR003167: Center for Clinical and Translational Sciences (CCTS) • The University of Texas Medical Branch at Galveston — UL1TR001439: The Institute for Translational Sciences • The University of Utah — UL1TR002538: Uhealth Center for Clinical and Translational Science • Tufts Medical Center — UL1TR002544: Tufts Clinical and Translational Science Institute • Tulane University — UL1TR003096: Center for Clinical and Translational Science • University Medical Center New Orleans — U54GM104940: Louisiana Clinical and Translational Science (LA CaTS) Center • University of Alabama at Birmingham — UL1TR003096: Center for Clinical and Translational Science • University of Arkansas for Medical Sciences — UL1TR003107: UAMS Translational Research Institute • University of Cincinnati — UL1TR001425: Center for Clinical and Translational Science and Training • University of Colorado Denver, Anschutz Medical Campus — UL1TR002535: Colorado Clinical and Translational Sciences Institute • University of Illinois at Chicago — UL1TR002003: UIC Center for Clinical and Translational Science • University of Kansas Medical Center — UL1TR002366: Frontiers: University of Kansas Clinical and Translational Science Institute • University of Kentucky — UL1TR001998: UK Center for Clinical and Translational Science • University of Massachusetts Medical School Worcester — UL1TR001453: The UMass Center for Clinical and Translational Science (UMCCTS) • University of Minnesota — UL1TR002494: Clinical and Translational Science Institute • University of Mississippi Medical Center — U54GM115428: Mississippi Center for Clinical and Translational Research (CCTR) • University of Nebraska Medical Center — U54GM115458: Great Plains IDeA-Clinical & Translational Research • University of North Carolina at Chapel Hill — UL1TR002489: North Carolina Translational and Clinical Science Institute • University of Oklahoma Health Sciences Center — U54GM104938: Oklahoma Clinical and Translational Science Institute (OCTSI) • University of Rochester — UL1TR002001: UR Clinical & Translational Science Institute • University of Southern California — UL1TR001855: The Southern California Clinical and Translational Science Institute (SC CTSI) • University of Vermont — U54GM115516: Northern New England Clinical & Translational Research (NNE-CTR) Network • University of Virginia — UL1TR003015: iTHRIV Integrated Translational health Research Institute of Virginia • University of Washington — UL1TR002319: Institute of Translational Health Sciences • University of Wisconsin-Madison — UL1TR002373: UW Institute for Clinical and Translational Research • Vanderbilt University Medical Center — UL1TR002243: Vanderbilt Institute for Clinical and Translational Research • Virginia Commonwealth University — UL1TR002649: C. Kenneth and Dianne Wright Center for Clinical and Translational Research • Wake Forest University Health Sciences — UL1TR001420: Wake Forest Clinical and Translational Science Institute • Washington University in St. Louis — UL1TR002345: Institute of Clinical and Translational Sciences • Weill Medical College of Cornell University — UL1TR002384: Weill Cornell Medicine Clinical and Translational Science Center • West Virginia University — U54GM104942: West Virginia Clinical and Translational Science Institute (WVCTSI)

Submitted: Icahn School of Medicine at Mount Sinai — UL1TR001433: ConduITS Institute for Translational Sciences • The University of Texas Health Science Center at Tyler — UL1TR003167: Center for Clinical and Translational Sciences (CCTS) • University of California, Davis — UL1TR001860: UCDavis Health Clinical and Translational Science Center • University of California, Irvine — UL1TR001414: The UC Irvine Institute for Clinical and Translational Science (ICTS) • University of California, Los Angeles — UL1TR001881: UCLA Clinical Translational Science Institute • University of California, San Diego — UL1TR001442: Altman Clinical and Translational Research Institute • University of California, San Francisco — UL1TR001872: UCSF Clinical and Translational Science Institute

Pending: Arkansas Children’s Hospital — UL1TR003107: UAMS Translational Research Institute • Baylor College of Medicine — None (Voluntary) • Children’s Hospital of Philadelphia — UL1TR001878: Institute for Translational Medicine and Therapeutics • Cincinnati Children’s Hospital Medical Center — UL1TR001425: Center for Clinical and Translational Science and Training • Emory University — UL1TR002378: Georgia Clinical and Translational Science Alliance • HonorHealth — None (Voluntary) • Loyola University Chicago — UL1TR002389: The Institute for Translational Medicine (ITM) • Medical College of Wisconsin — UL1TR001436: Clinical and Translational Science Institute of Southeast Wisconsin • MedStar Health Research Institute — UL1TR001409: The Georgetown-Howard Universities Center for Clinical and Translational Science (GHUCCTS) • MetroHealth — None (Voluntary) • Montana State University — U54GM115371: American Indian/Alaska Native CTR • NYU Langone Medical Center — UL1TR001445: Langone Health’s Clinical and Translational Science Institute • Ochsner Medical Center — U54GM104940: Louisiana Clinical and Translational Science (LA CaTS) Center • Regenstrief Institute — UL1TR002529: Indiana Clinical and Translational Science Institute • Sanford Research — None (Voluntary) • Stanford University — UL1TR003142: Spectrum: The Stanford Center for Clinical and Translational Research and Education • The Rockefeller University — UL1TR001866: Center for Clinical and Translational Science • The Scripps Research Institute — UL1TR002550: Scripps Research Translational Institute • University of Florida — UL1TR001427: UF Clinical and Translational Science Institute • University of New Mexico Health Sciences Center — UL1TR001449: University of New Mexico Clinical and Translational Science Center • University of Texas Health Science Center at San Antonio — UL1TR002645: Institute for Integration of Medicine and Science • Yale New Haven Hospital — UL1TR001863: Yale Center for Clinical Investigation.

## Supplementary Material

### Data Harmonization and Deduplication Strategy

Three main categories of data were harmonized across CMS and multiple EHRs:

1. Conditions, Comorbidities, Measurements, Observations, Lab Values, Drug Exposures
2. COVID-19-Related details (index dates and vaccinations)
3. Demographics and Mortality

#### 1. Conditions, Comorbidities, Measurements, Observations, Lab Values, Drug Exposures

To deduplicate multiple N3C IDs pertaining to the same individual into a single N3C record, this first category of data was consolidated as follows:

- Most comorbidities were treated as binary indicators. For each patient, the maximum value of the condition was used across multiple EHR records (grouped by the CMS ID) For example, if a patient had a record of a diagnosis of cancer in one EHR source but not another, the patient was assumed to have a history of cancer.
- For laboratory values, the maximum value was used only if all records fall within the measurable or reasonable range of the laboratory measure. Specifically:

◦ The acceptable range for each lab value was first identified based on clinical plausibility.
◦ Extreme or impossible laboratory values were handled by nullifying or truncating them up to a maximum value of +/-4 standard deviations.
◦ The maximum valid value across patients’ EHR records was used for each patient.

After deduplication of individuals with multiple N3C IDs, the remaining patient data was consolidated along with the harmonization of the CMS data. The following sections outline the process of harmonization for single or multiple N3C EHR records with the relevant CMS record per patient for the second two categories of data (COVID-19-related details and demographics and mortality).

#### 2. COVID-19-Related Details

##### COVID-19 Index Date

- To focus on patients’ first instance of a positive COVID-19 indicator, the record with the earliest COVID-19 index date (across the CMS and duplicate EHR records of the patient) was used.
- Subsequent index dates that appeared on patients’ other records were treated as potential reinfections if they occured more than 30 days after the previous index date.

##### Vaccination

The CMS data may provide additional dates regarding vaccination. Vaccination status was matched according to the selected COVID-19 index date. Aggregating the existing vaccination columns in the patient summary-level table (e.g., **number_of_COVID_vaccine_doses_before_or_day_of_covid**) across all the patients’ EHR and CMS records was not appropriate if the COVID-19 index date differs between multiple EHR records.

For example: The value of the **number_of_COVID_vaccine_doses_before_or_day_of_covid** column might be two for a COVID-19-index day of February 1, 2023 for one of the EHR records. If a duplicate EHR record for this patient revealed an earlier COVID-index day of April 1, 2022, the corresponding value of this column would be 0. It would be incorrect to take the maximum value of **number_of_COVID_vaccine_doses_before_or_day_of_covid** (which would be two) across all the EHR records because the patient was in fact NOT vaccinated before their real and earlier COVID-19 index date of April 1, 2022 (the vaccination likely occurred between these two dates), so we need to apply another strategy.

The best strategy to ensure the appropriate number of vaccines was matched in relation to the patients’ correct (i.e., earliest) COVID-19-index day, was to perform the aggregation of multiple EHR records on the visit level table (**cohort_all_facts_table**):

- Specifically, the column **had_vaccine_administered** was aggregated by taking the maximum value of this column across all the patients’ EHR records (grouped by their single CMS ID)
- The aggregation steps were followed to create the patient level summary table, or was added as additional code in that node.
- Next, ensure the patients’ final COVID-19 index date was used (see above) and that it populated the column **COVID_first_poslab_or_diagnosis_date** before running the code for the **COVID_Patient_Summary_Table** node.
- This ensured that the right number of vaccinations was coded in relation to the patients’ final and correct COVID-19 index date

#### 3. Demographics and Mortality

##### Date of Birth

- If there was conflict among EHRs, the CMS date of birth was taken.
- If the CMS date of birth is missing, the EHR date of birth that occurs first chronologically was taken.
- If one record indicates a current age younger than 65, whereas the other indicates a current age ≥ 65, the latter EHR record’s date of birth was taken.

##### Race/Ethnicity

- The CMS race/ethnicity category was used if it was missing for all EHR records.
- When there is conflict in the race/ethnicity between the CMS and EHR:

◦ The EHR race/ethnicity was used if these match because the EHR race/ethnicity has more categories and thus provides richer information
◦ If the race/ethnicity records conflict among the EHR records:

▪ The more informative race/ethnicity category was taken - for example if one record is “Asian”, and the other is unknown or Other - then “Asian” was used.
▪ If the two records had conflicting race/ethnicity categories (e.g., “African” and “Asian” then the CMS race/ethnicity was used to break the tie.
▪ If the CMS race/ethnicity is missing and cannot break the tie, then the race/ethnicity from the record with the longest healthcare history (observation history) was chosen.
▪ If the length of the observation periods match in the EHR records, the race/ethnicity from the record with most patient information available (across demographics and comorbidities) was kept.
▪ If the records have equivalent information, the race/ethnicity from the record having the earliest chronological age was defined.
▪ If the ages match in the EHR records, the race/ethnicity from the record with the earliest COVID-19 index date was used.
▪ If the COVID-19 index dates match, then the races were combined into a supercategory (e.g., African and Asian)

##### Sex

- The CMS record was given priority for defining patients’ sex
- If no CMS sex data was present, and sex information was conflicting between multiple EHRs

◦ The sex associated with the EHR record with the older chronological age was taken.
◦ If the ages match in the EHR records, then the sex from the record with the longest healthcare history (observation history) was defined.
◦ If the length of the observation periods match in the EHR records, the sex from the record with most patient information available (across demographics and comorbidities) was kept.
◦ If the records have equivalent information, the sex from the record having the earliest chronological age was defined.
◦ If the ages match between the records, sex category from the record with the earliest COVID-19 index date was defined.
◦ If the COVID-19 index dates match, then sex was converted to Missing.

##### Zip Codes

- The CMS record was given priority for defining patients’ ZIP
- If ZIP codes were missing, the record (EHR or CMS) which has a ZIP code available was used (i.e. this was when all but one record had missing ZIP code information).
- When ZIP codes among EHR records were conflicted (and were missing from the CMS) the ZIP code from the EHR record was taken using the following prioritization:

◦ The EHR record having the most recent visit was taken.
◦ If the most recent visits are within 1 month of each other, the EHR record with the longest observation period was used.
◦ If these match, the EHR record with the most patient information available across demographics and comorbidities was used.
◦ If these match, the EHR record with the earliest chronological age was used.
◦ If these match, the EHR record that has the LATEST COVID-19 index date was used.

##### Mortality

- Conflicts in data were resolved by assigning a hierarchy to data sources as follows: EHR>SSA>claims>private media>private obituary data>obituary.com.
- If there are multiple death dates within a single source the earliest date was used.

### Variable selection strategy for propensity model variables

Prior to fitting the propensity model, we undertook variable selection to reduce the number of variables in the model by first implementing lasso logistic regression. The aim was to include only those covariates that were predictive of treatment receipt. Lasso is a type of regularized regression (i.e., L1 regression) that decreases model complexity by minimizing the sum of the absolute value of the coefficients. The consequence is that the coefficients for some predictors are shrunk towards zero. We thus retained only those covariates with non-zero coefficients. We then fit univariate logistic regression between each covariate and the primary outcome of ischemic stroke, and retained covariates with a coefficient absolute value greater than the natural log of 1.1 to identify *potential* confounders related to our primary outcome of interest. Inclusion of covariates related to the outcome can increase the precision of the treatment effect estimate(16). The list of variables selected for each data source is shown in Supplementary Table 3.

**Supplementary Figure 1:**
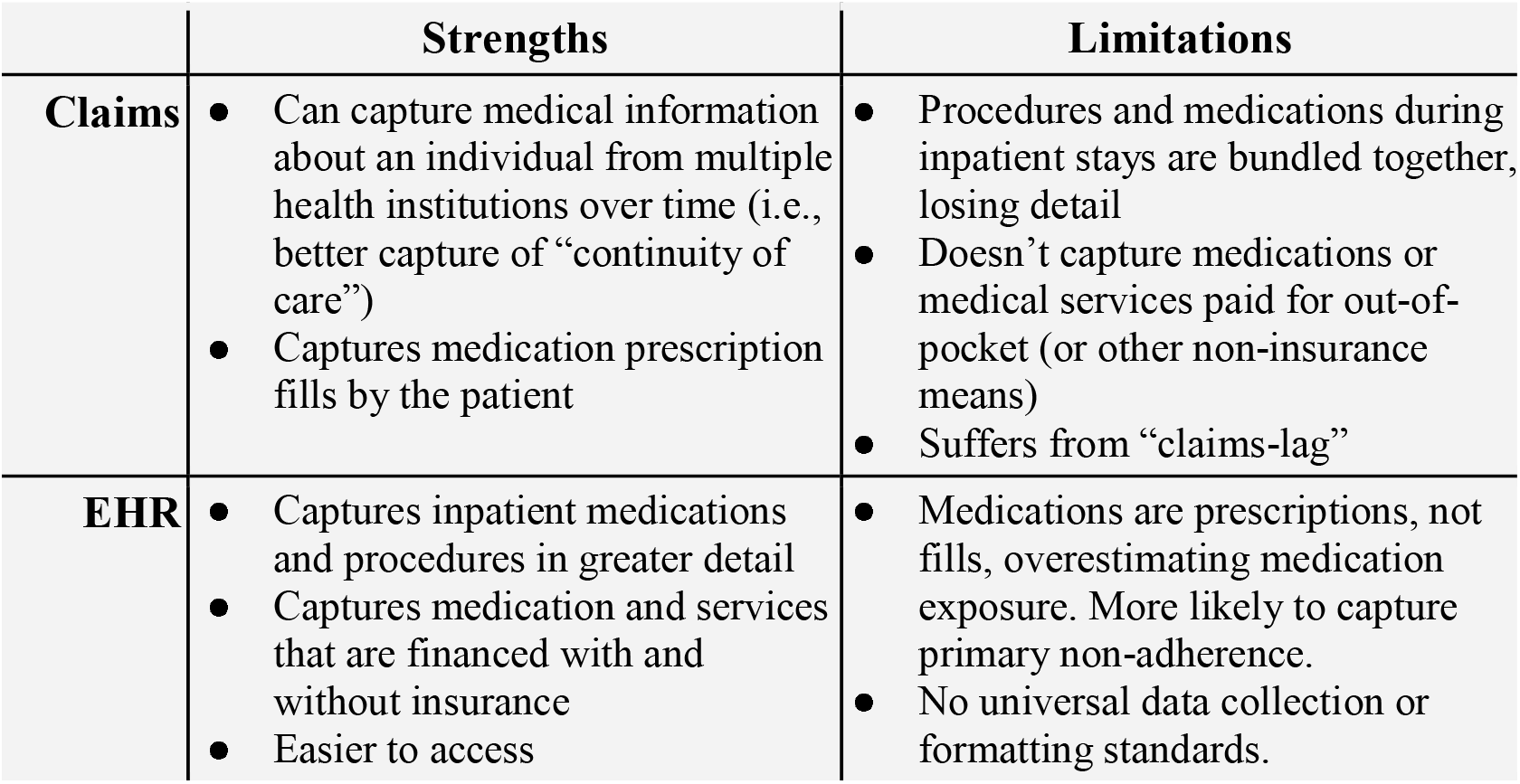
Comparison of strengths and limitations of EHR and claims data.

**Supplementary Figure 2:**
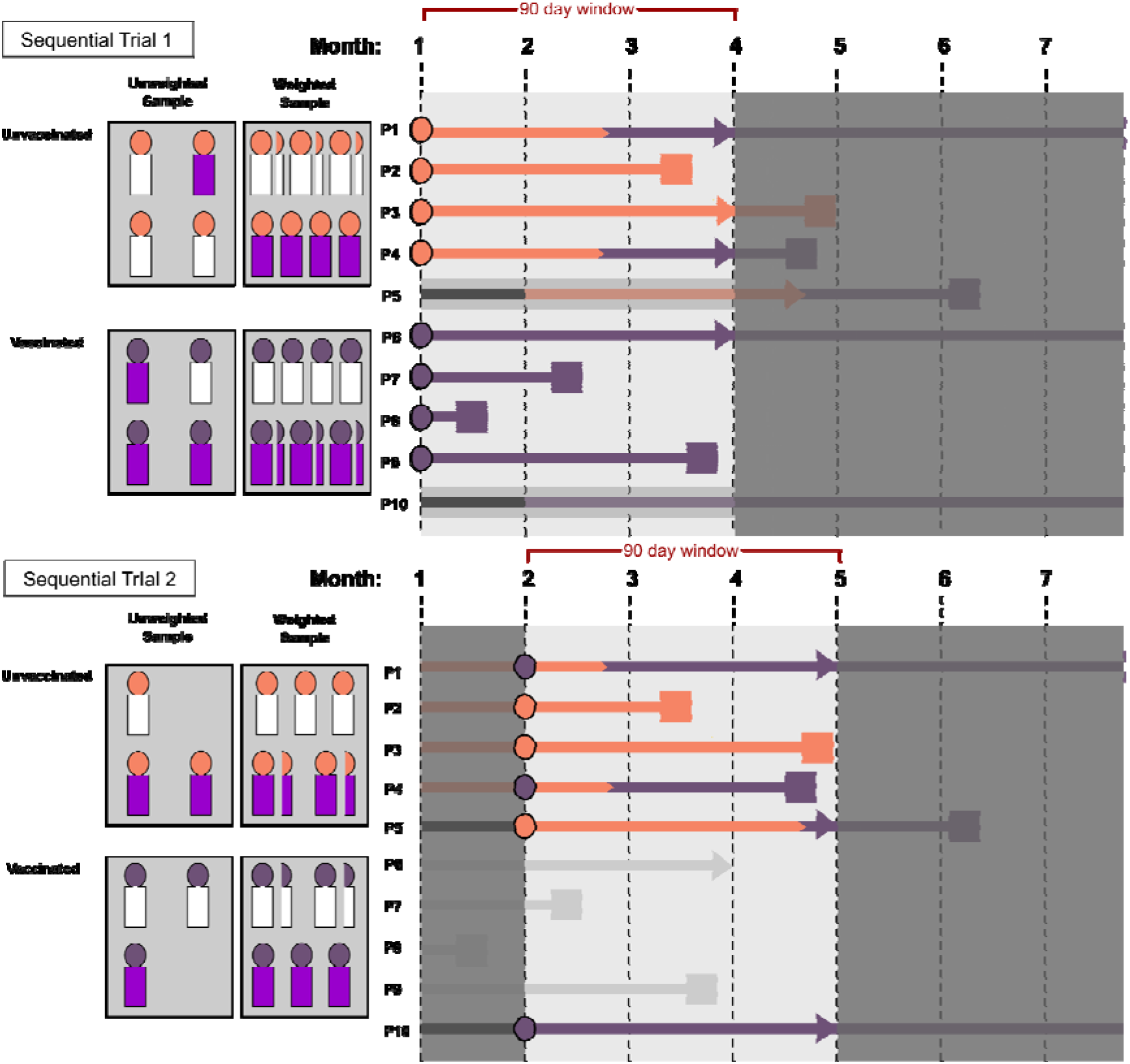
The TTE design for the sequential trial metho d, with two sequential trials displayed. Sequential Trial 1 begins on Month 1 where eligible, unvaccinate d (orange ovals) and vaccinated (purple ovals) individuals are identified. All individuals except 5 and 10 are eligible for Trial 1. Prior to weighting, the distribution of measured covariates such as diabetes (purple shading) differs between treatment groups as shown in the Unweighted Sample. After weighting, however, the distribution of covariates is balanced between the treatment groups as shown in the Weighted Sample. Each patient is followed for 90 days (arrow) or until the occurrence of stroke (square). The treatment effect is estimated as the difference in the risk of ischemic stroke between treatment groups in the weighted sample. The second sequential trial begins on Month 2. Patients 1-4 from Trial 1 are eligible for entry as they meet eligibility criteria, are still at risk of ischemic stroke, and were not vaccinated prior. Patients 5 and 10 now meet eligibility criteria and enter Trial 2. Again, unvaccinated and vaccinated patients are identified and assigned to the treated and untreated groups, respectively. Weighting is used again to ensure the distribution of measured covariates is balanced. The treatment effect is estimated similarly for each sequential trial.

**Supplementary Figure 3:**
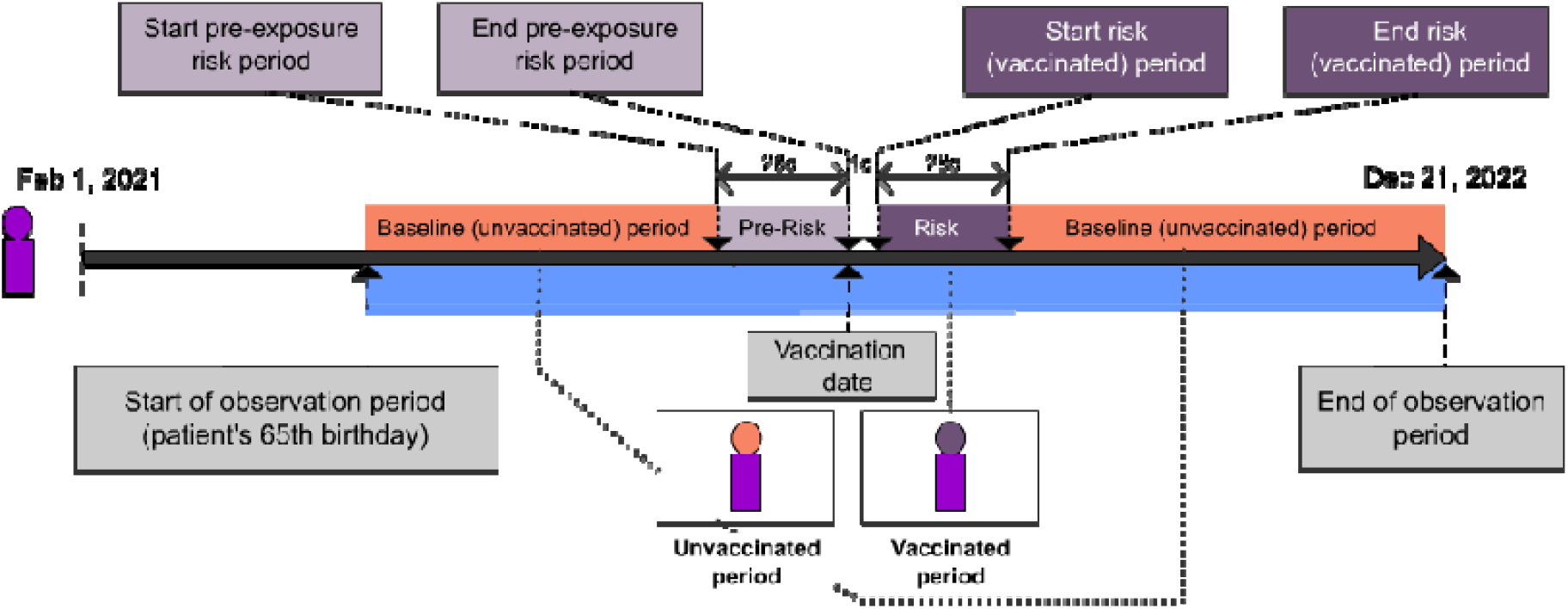
SCCS analysis design overview. Timeline presents the partition of the patient’s observation period to: (i) pre-exposure risk period - the 28 days prior to vaccination, (ii) risk period - the 28-day period following vaccination, and (iii) baseline (non-risk) periods - the remainder of the observation period, before the pre-exposure period and after the risk period. The treatment effect is estimated as the relative risk of ischemic stroke between the (vaccination) risk period and the (unvaccinated) baseline period. Since the comparison is made within patients, fixed covariates, either measured (e.g., diabetes, in purple) or unmeasured are identical between vaccinated and unvaccinated periods meaning there is no need to balance their distribution as in the TTE.

**Supplementary Figure 4:**
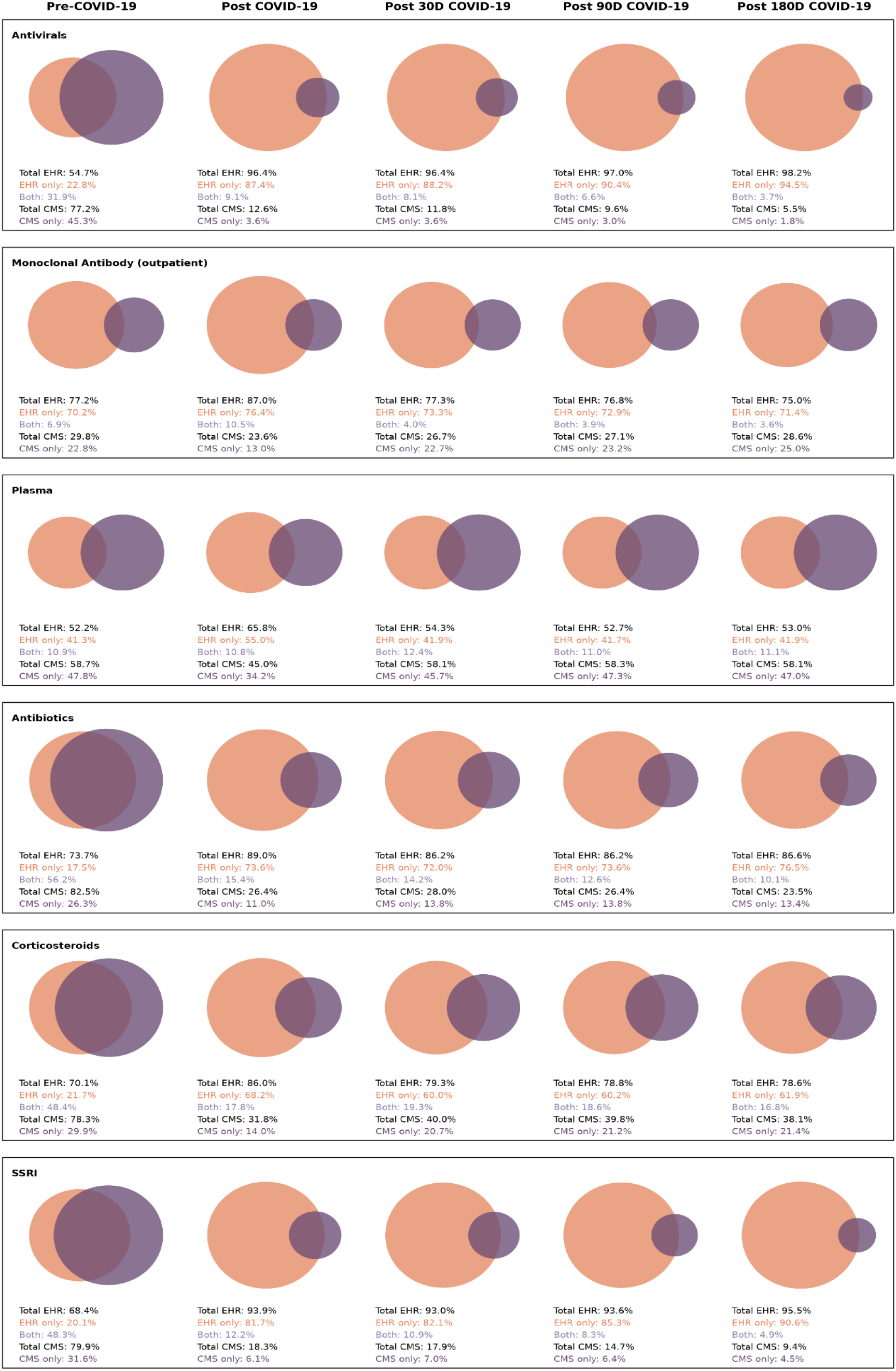
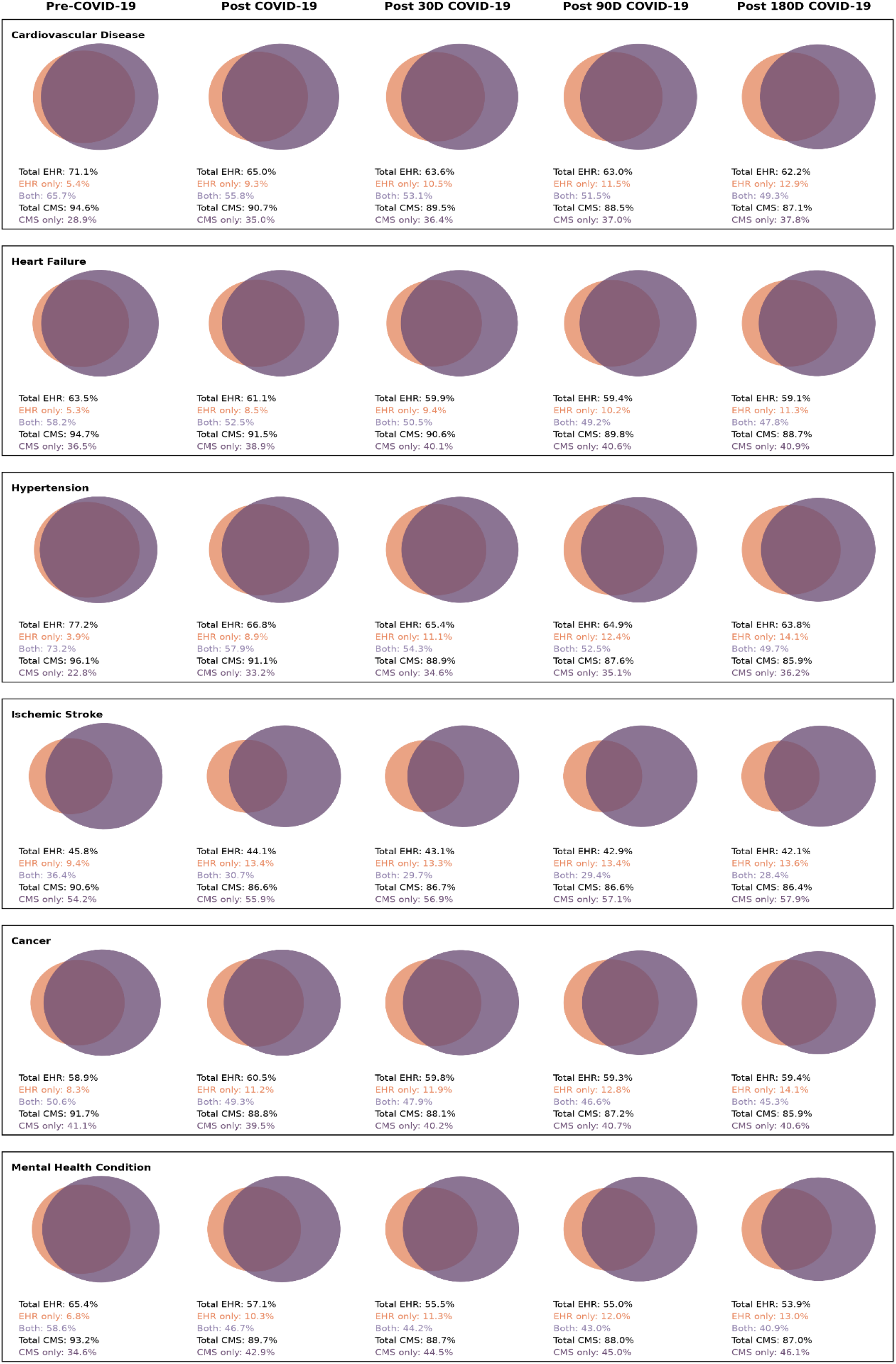
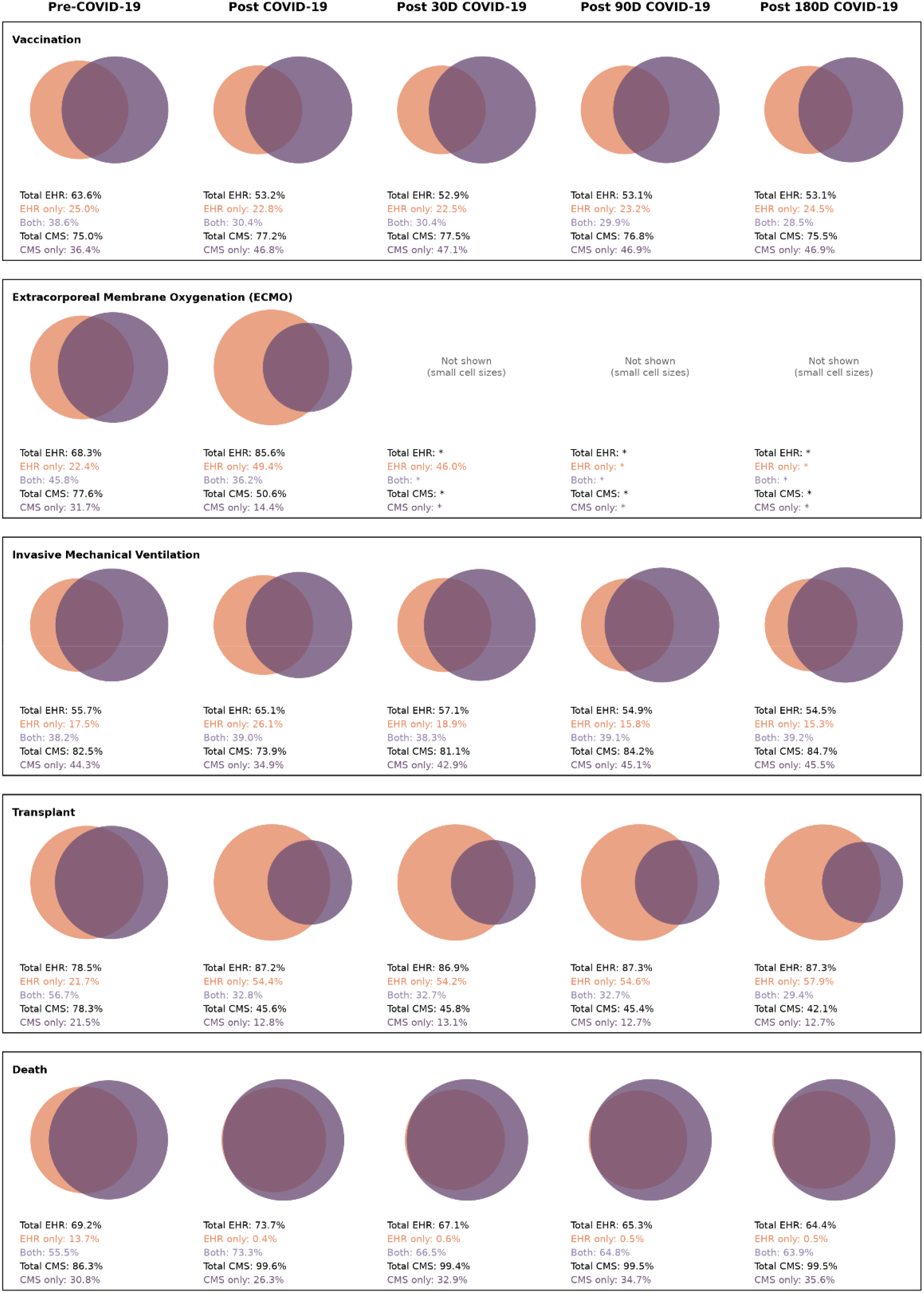
Venn Diagrams for extended variables showing percentages of total data (per variable) that are present in EHR (orange), CMS (dark purple), or both (light purple) data sources. From left to right, Venn diagrams show data pre-COVID-19 diagnosis, immediately post-COVID-19 diagnosis through the end of follow-up, 30 days post-COVID-19 diagnosis through the end of follow-up, 90 days post-COVID-19 diagnosis through the end of follow-up, and 180 days post-COVID-19 diagnosis through the end of follow-up. Monoclonal antibodies include only COVID-19-related treatments. Plasma is convalescent plasma medication.

**Supplementary Table 1:** Concepts included in the ischemic stroke codeset.

| concept_id | concept_name |
| --- | --- |
| 761792 | Cerebrovascular accident due to thrombus of right posterior cerebral artery |
| 762345 | Left vertebral artery thrombosis with stroke |
| 37200499 | Cerebral infarction due to embolism of bilateral carotid arteries |
| 42535112 | Cerebrovascular accident due to occlusion of left middle cerebral artery |
| 3188623 | CVA with associated bilateral paralysis |
| 762344 | Cerebrovascular accident due to thrombus of right vertebral artery |
| 761798 | Basilar artery embolism with stroke |
| 45548029 | Cerebral infarction due to embolism of right posterior cerebral artery |
| 43530669 | Ischemic stroke with coma |
| 45572097 | Cerebral infarction due to embolism of left carotid artery |
| 3176312 | Left middle cerebral artery stroke |
| 37200498 | Cerebral infarction due to embolism of bilateral vertebral arteries |
| 43531605 | Occlusion of cerebral artery with stroke |
| 762340 | Left anterior cerebral artery embolism with stroke |
| 3184799 | CNS infarction due to occlusion of vertebral artery |
| 761836 | Cerebrovascular accident due to occlusion of right posterior communicating artery |
| 42535461 | Cerebrovascular accident due to occlusion of left vertebral artery |
| 42539262 | Cerebrovascular accident due to occlusion of left carotid artery |
| 37110679 | Cerebral ischemic stroke due to stenosis of extracranial large artery |
| 761785 | Cerebrovascular accident due to occlusion of left anterior cerebral artery |
| 761791 | Left cerebellar artery thrombosis with stroke |
| 765281 | Cerebrovascular accident due to occlusion of right anterior cerebral artery |
| 45538395 | Cerebral infarction due to embolism of right vertebral artery |
| 45557551 | Cerebral infarction due to embolism of left vertebral artery |
| 45596212 | Cerebral infarction due to embolism of right carotid artery |
| 762339 | Right anterior cerebral artery embolism with stroke |
| 4108356 | Cerebral infarction due to embolism of cerebral arteries |
| 42535503 | Cerebrovascular accident due to occlusion of right middle cerebral artery by embolus |
| 3188677 | Cerebral ischemia complicating stroke |
| 42539195 | Cerebrovascular accident due to occlusion of left posterior cerebral artery |
| 3184775 | Cerebral infarct complicating carotid stenosis |
| 37312016 | Cerebral ischemic stroke due to aortic arch embolism |
| 45543190 | Cerebral infarction due to embolism of right anterior cerebral artery |
| 42535110 | Cerebrovascular accident due to occlusion of right posterior cerebral artery |
| 45552803 | Cerebral infarction due to embolism of unspecified cerebral artery |
| 381316 | Cerebrovascular accident |
| 45581783 | Cerebral infarction due to embolism of unspecified posterior cerebral artery |
| 45596214 | Cerebral infarction due to embolism of unspecified anterior cerebral artery |
| 3187113 | CVA with associated paralysis of dominant side |
| 42535504 | Cerebrovascular accident due to occlusion of left middle cerebral artery by embolus |
| 4110192 | Cerebral infarction due to thrombosis of cerebral arteries |
| 45552805 | Cerebral infarction due to embolism of left cerebellar artery |
| 4138327 | Acute lacunar infarction |
| 761796 | Right carotid artery embolism with stroke |
| 37395576 | Cerebrovascular accident due to left carotid artery stenosis |
| 42535146 | Cerebrovascular accident due to occlusion of left pontine artery |
| 42535113 | Cerebrovascular accident due to thrombus of right middle cerebral artery |
| 45767658 | Cerebral infarction due to thrombosis of middle cerebral artery |
| 42535149 | Cerebrovascular accident due to occlusion of right cerebellar artery |
| 4006295 | Nonparalytic stroke |
| 4045735 | Anterior cerebral circulation infarction |
| 4110196 | Right sided cerebral hemisphere cerebrovascular accident |
| 45562367 | Cerebral infarction due to embolism of right middle cerebral artery |
| 761835 | Cerebrovascular accident due to occlusion of left posterior communicating artery |
| 4211509 | Cardioembolic stroke |
| 42535458 | Cerebrovascular accident due to stenosis of right vertebral artery |
| 37395574 | Cerebrovascular accident due to right carotid artery occlusion |
| 761795 | Left vertebral artery embolism with stroke |
| 45576889 | Cerebral infarction due to embolism of unspecified middle cerebral artery |
| 3179950 | Right subthalamic lacunar stroke |
| 42535511 | Cerebrovascular accident due to thrombus of basilar artery |
| 37200507 | Cerebral infarction due to embolism of bilateral anterior cerebral arteries |
| 42535111 | Cerebrovascular accident due to occlusion of right middle cerebral artery |
| 45601045 | Cerebral infarction due to embolism of other cerebral artery |
| 4110190 | Cerebral infarction due to embolism of precerebral arteries |
| 37200506 | Cerebral infarction due to embolism of bilateral middle cerebral arteries |
| 37110678 | Cerebral ischemic stroke due to occlusion of extracranial large artery |
| 42535507 | Cerebrovascular accident due to occlusion of right cerebellar artery by embolus |
| 37209562 | Cerebrovascular accident due to stenosis of bilateral carotid arteries |
| 45586594 | Cerebral infarction due to embolism of left middle cerebral artery |
| 42535460 | Cerebrovascular accident due to right vertebral artery occlusion |
| 45576884 | Cerebral infarction due to embolism of unspecified carotid artery |
| 4159140 | Thrombotic stroke |
| 42535147 | Cerebrovascular accident due to occlusion of right pontine artery |
| 37312015 | Cerebral ischemic stroke due to global hypoperfusion with watershed infarct |
| 4310996 | Ischemic stroke |
| 42535506 | Cerebrovascular accident due to occlusion of left posterior cerebral artery by embolus |
| 37110239 | Cerebral ischemic stroke due to extracranial large artery atherosclerosis |
| 37309665 | Cerebrovascular accident due to occlusion of bilateral pontine arteries |
| 3174271 | Right middle cerebral artery stroke |
| 4045734 | CVA - cerebrovascular accident due to cerebral artery occlusion |
| 37312014 | Cerebral ischemic stroke due to hypercoagulable state |
| 4046360 | Lacunar infarction |
| 42535148 | Cerebrovascular accident due to occlusion of left cerebellar artery |
| 37200509 | Cerebral infarction due to embolism of bilateral cerebellar arteries |
| 42535505 | Cerebrovascular accident due to occlusion of right posterior cerebral artery by embolus |
| 4046359 | Partial anterior cerebral circulation infarction |
| 37309657 | Cerebrovascular accident due to stenosis of bilateral vertebral arteries |
| 45548030 | Cerebral infarction due to embolism of left posterior cerebral artery |
| 42535459 | Cerebrovascular accident due to stenosis of left vertebral artery |
| 45605806 | Cerebral infarction due to embolism of other precerebral artery |
| 45552804 | Cerebral infarction due to embolism of left anterior cerebral artery |
| 37312017 | Cerebral ischemic stroke due to dissection of artery |
| 4046358 | Total anterior cerebral circulation infarction |
| 761793 | Left posterior cerebral artery thrombosis with stroke |
| 4023571 | Paralytic stroke |
| 3185422 | Paralysis of nondominant side complicating stroke |
| 4145897 | Multiple lacunar infarcts |
| 45562364 | Cerebral infarction due to embolism of unspecified vertebral artery |
| 761794 | Right vertebral artery embolism with stroke |
| 761797 | Left carotid artery embolism with stroke |
| 4045736 | Posterior cerebral circulation infarction |
| 45581782 | Cerebral infarction due to embolism of basilar artery |
| 4045737 | Pure motor lacunar infarction |
| 42535508 | Cerebrovascular accident due to occlusion of left cerebellar artery by embolus |
| 761789 | Cerebrovascular accident due to thrombus of right cerebellar artery |
| 3188894 | Precerebral artery occlusion w/infarct |
| 43530670 | Ischemic stroke without coma |
| 45572099 | Cerebral infarction due to embolism of unspecified cerebellar artery |
| 443454 | Cerebral infarction |
| 37110237 | Cerebral ischemic stroke due to small artery occlusion |
| 4045738 | Pure sensory lacunar infarction |
| 4153352 | Embolic stroke |
| 37200508 | Cerebral infarction due to embolism of bilateral posterior cerebral arteries |
| 4112022 | Left sided cerebral hemisphere cerebrovascular accident |
| 42535512 | Cerebrovascular accident due to thrombus of right carotid artery |
| 37395575 | Cerebrovascular accident due to right carotid artery stenosis |
| 4110189 | Cerebral infarct due to thrombosis of precerebral arteries |
| 37110238 | Cerebral ischemic stroke due to intracranial large artery atherosclerosis |
| 42535114 | Cerebrovascular accident due to thrombus of left middle cerebral artery |
| 42539166 | Cerebrovascular accident due to thrombus of left carotid artery |
| 45601044 | Cerebral infarction due to embolism of right cerebellar artery |
| 4043731 | Infarction - precerebral |
| 37110765 | Ischemic stroke without residual deficits |
| 43531607 | Cerebral infarction due to stenosis of carotid artery |

**Supplementary Table 2:**
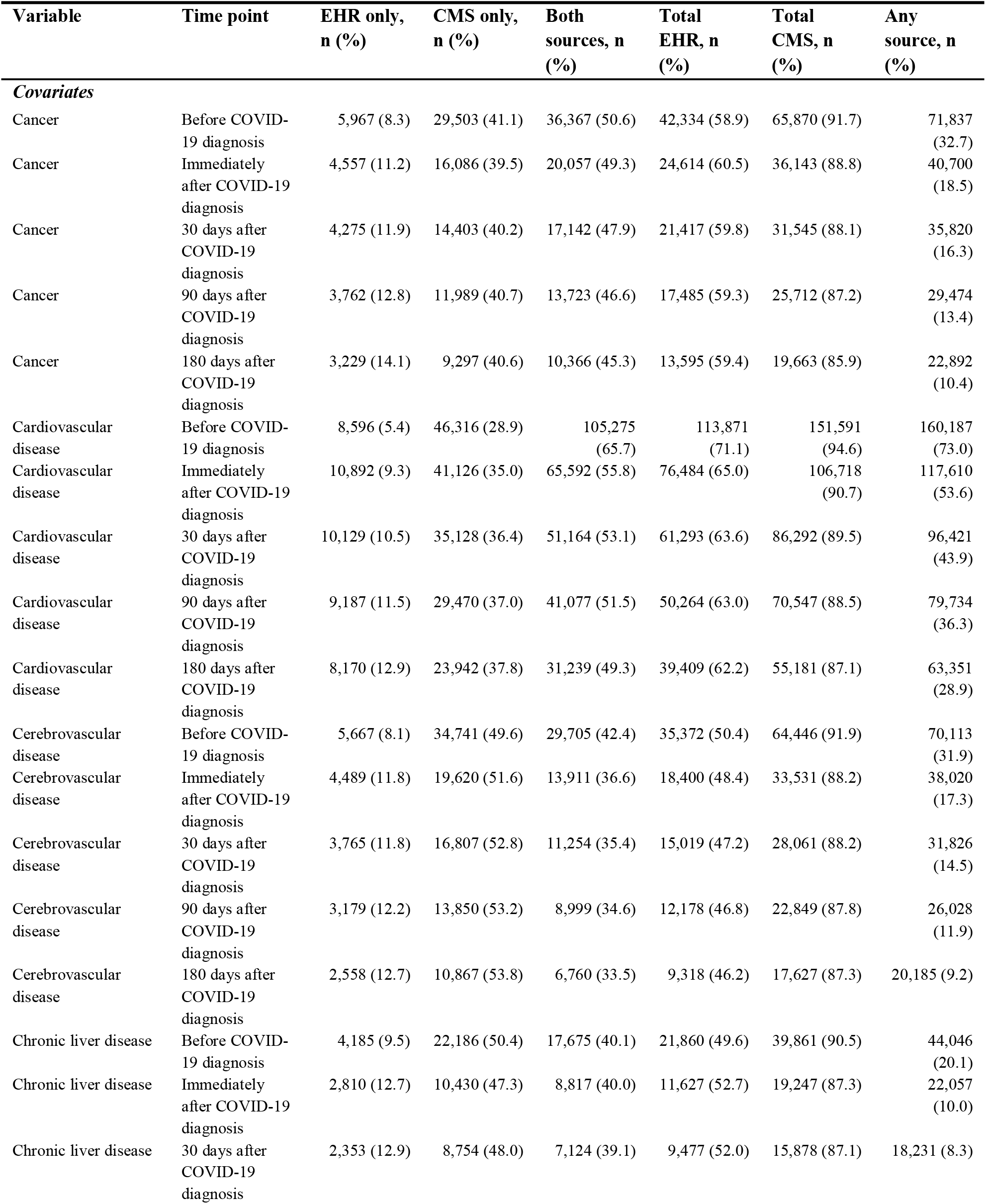
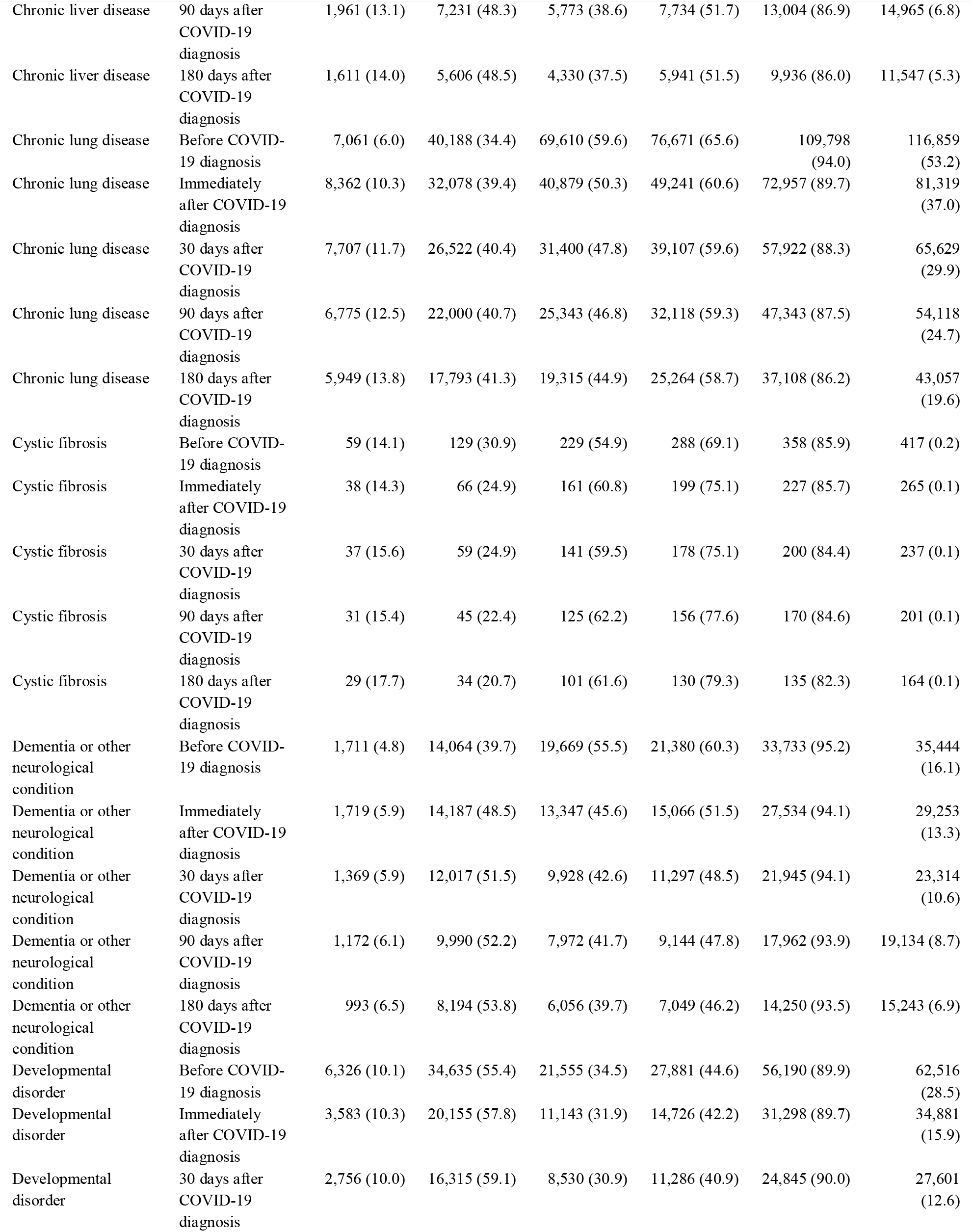
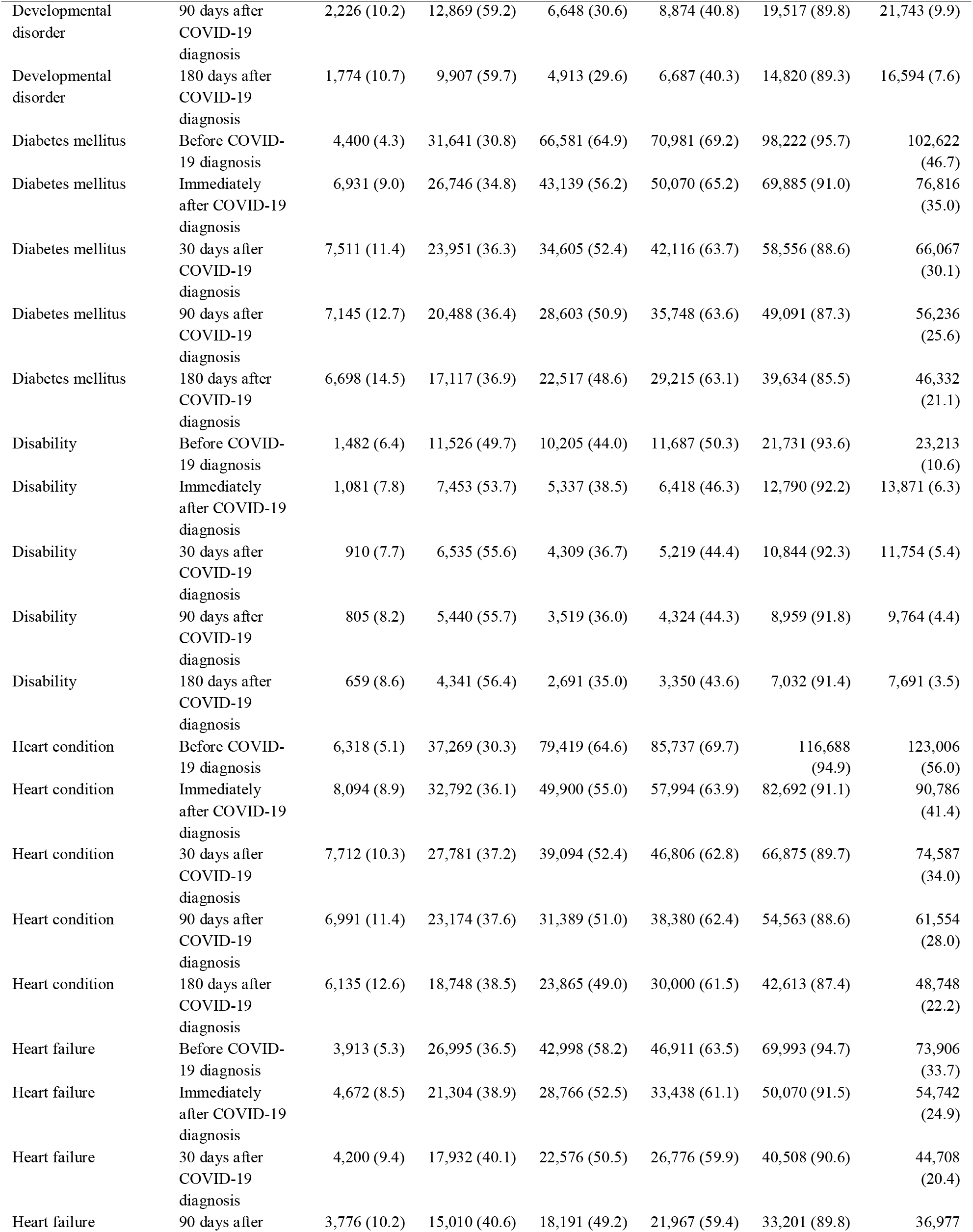
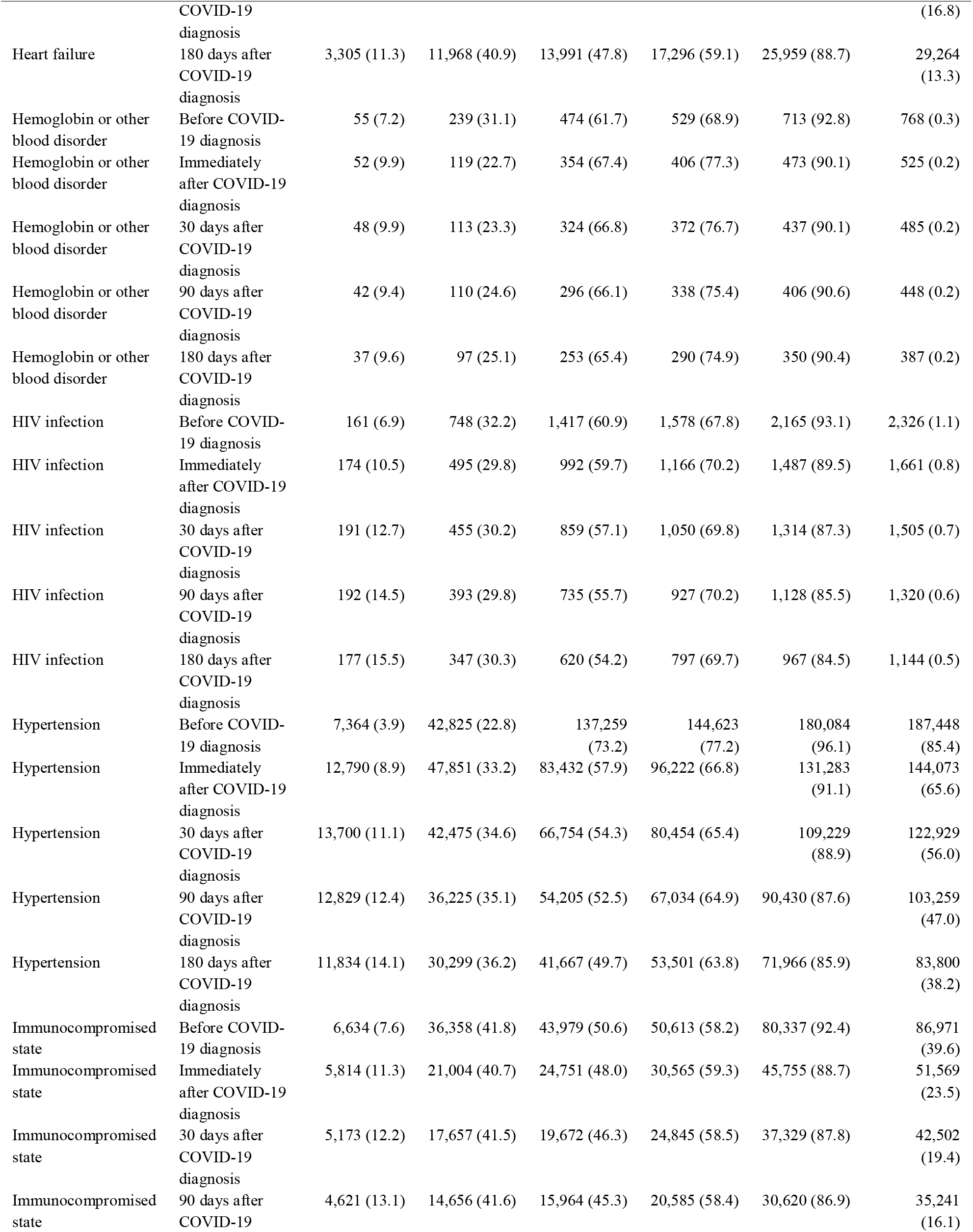
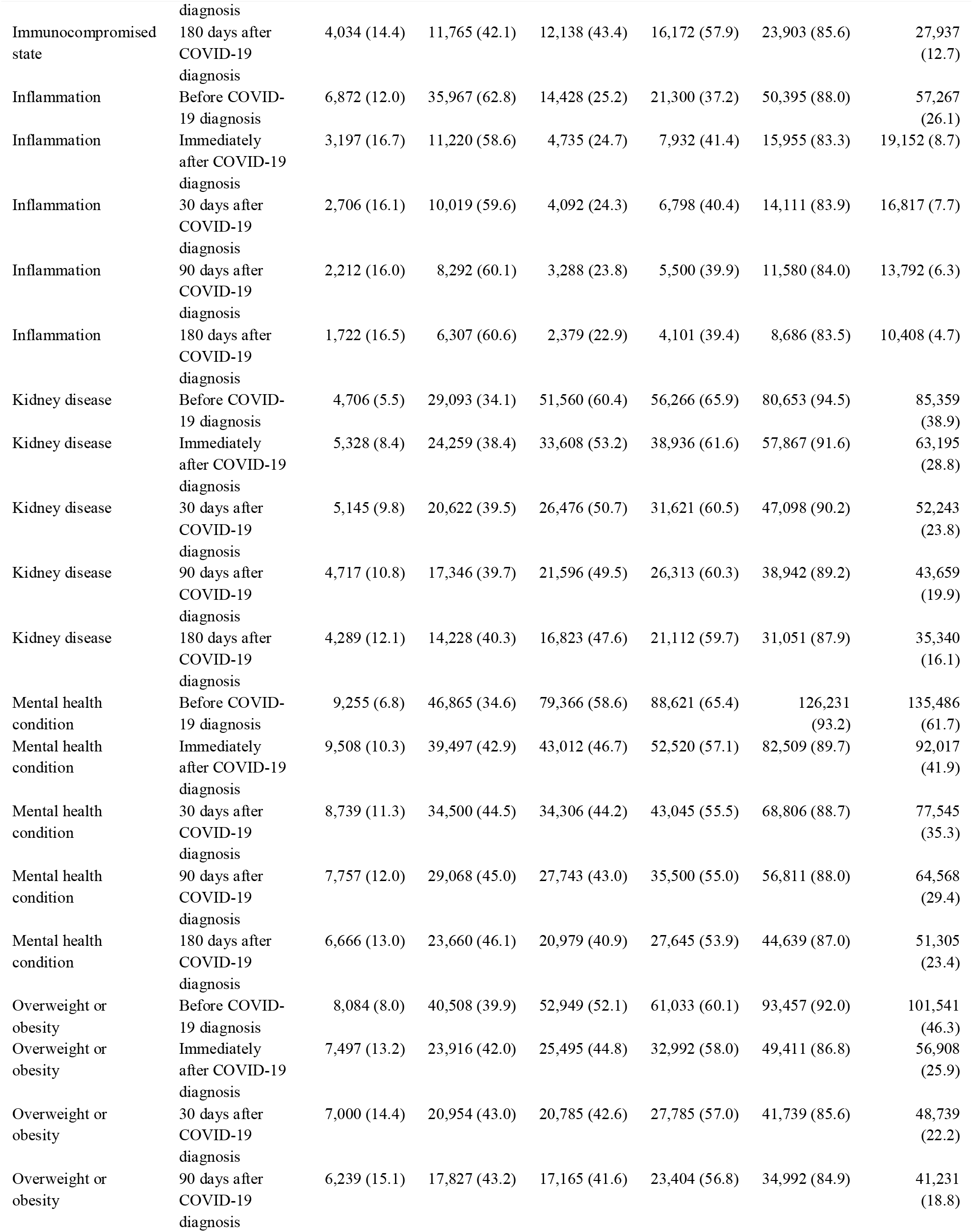
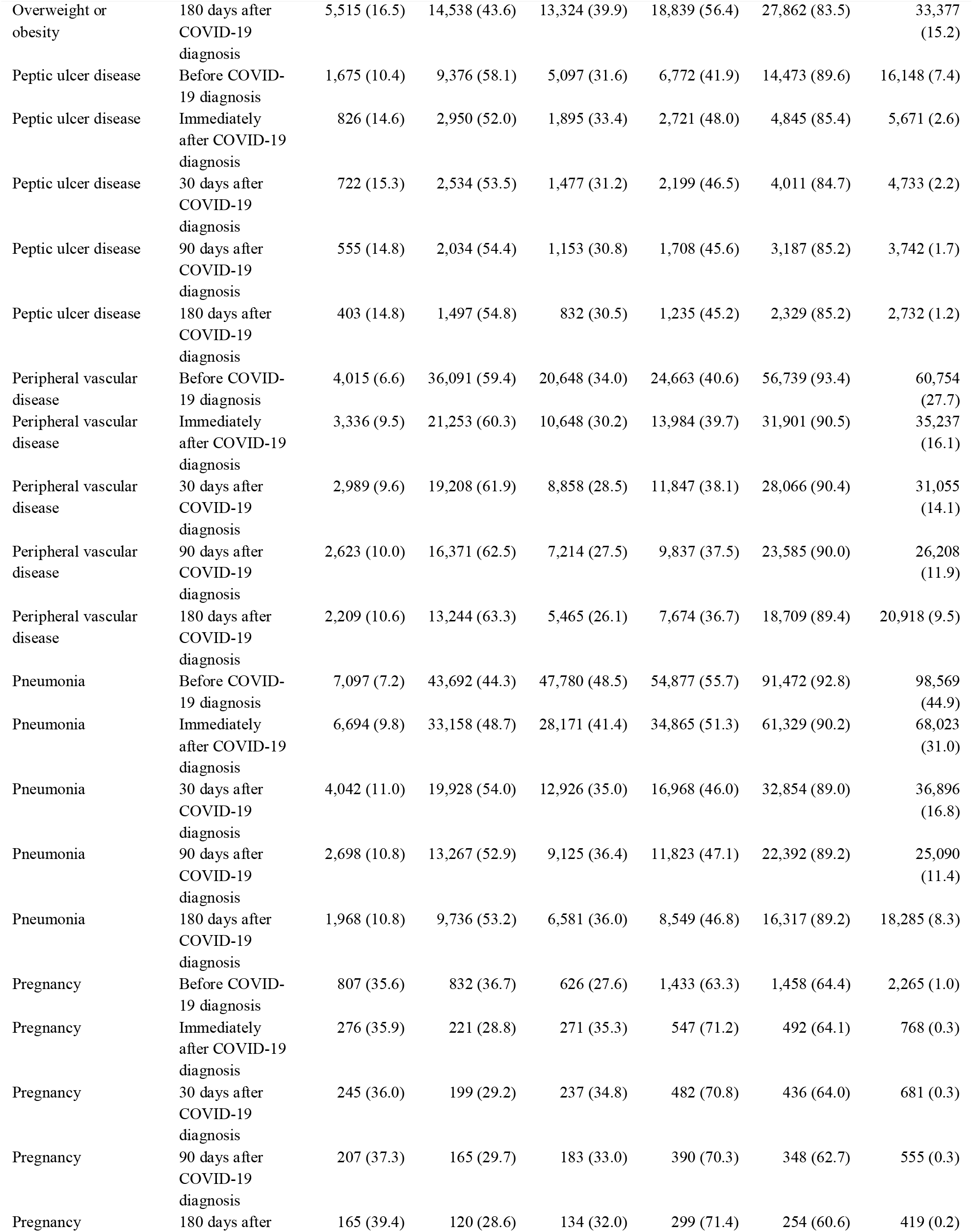
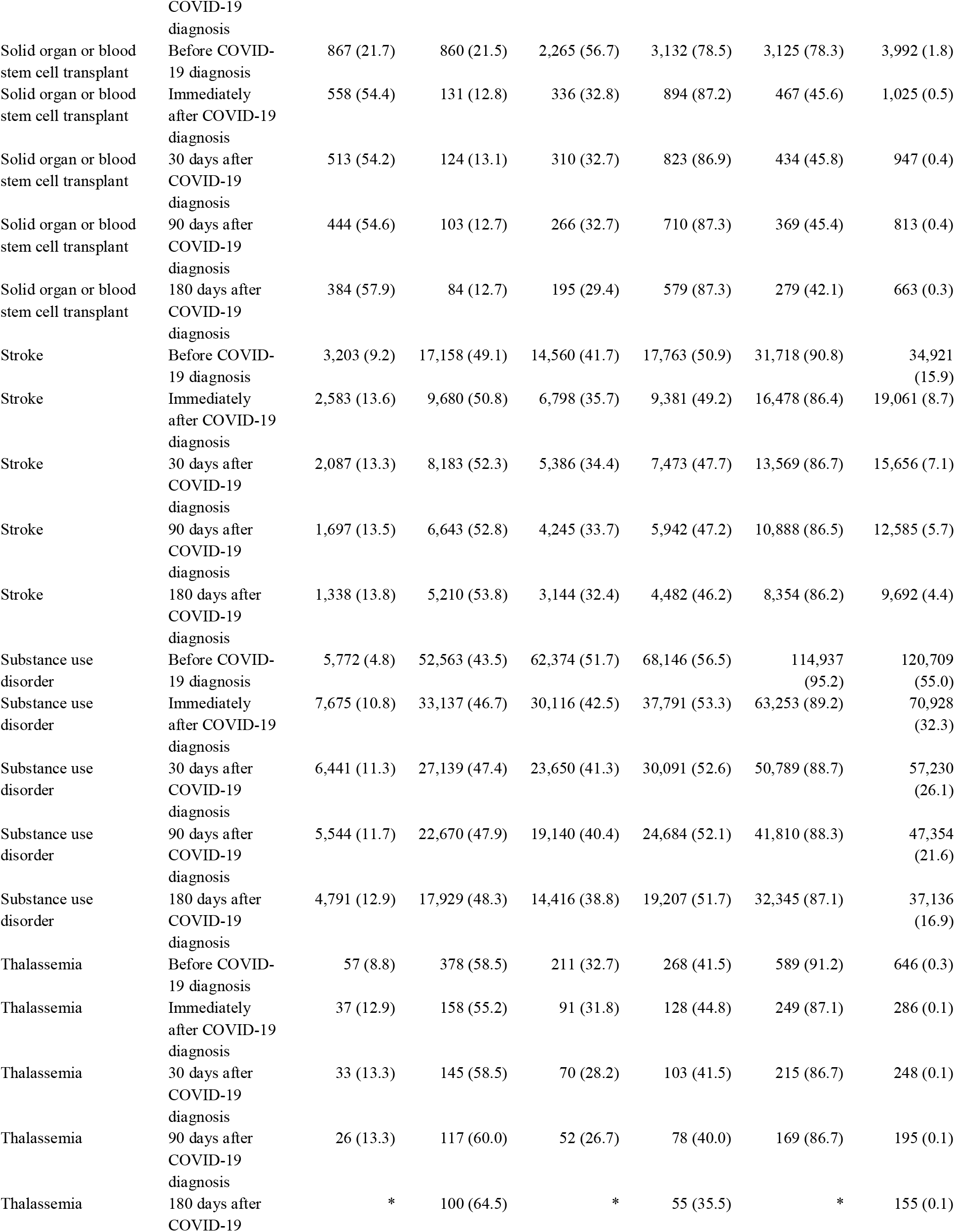
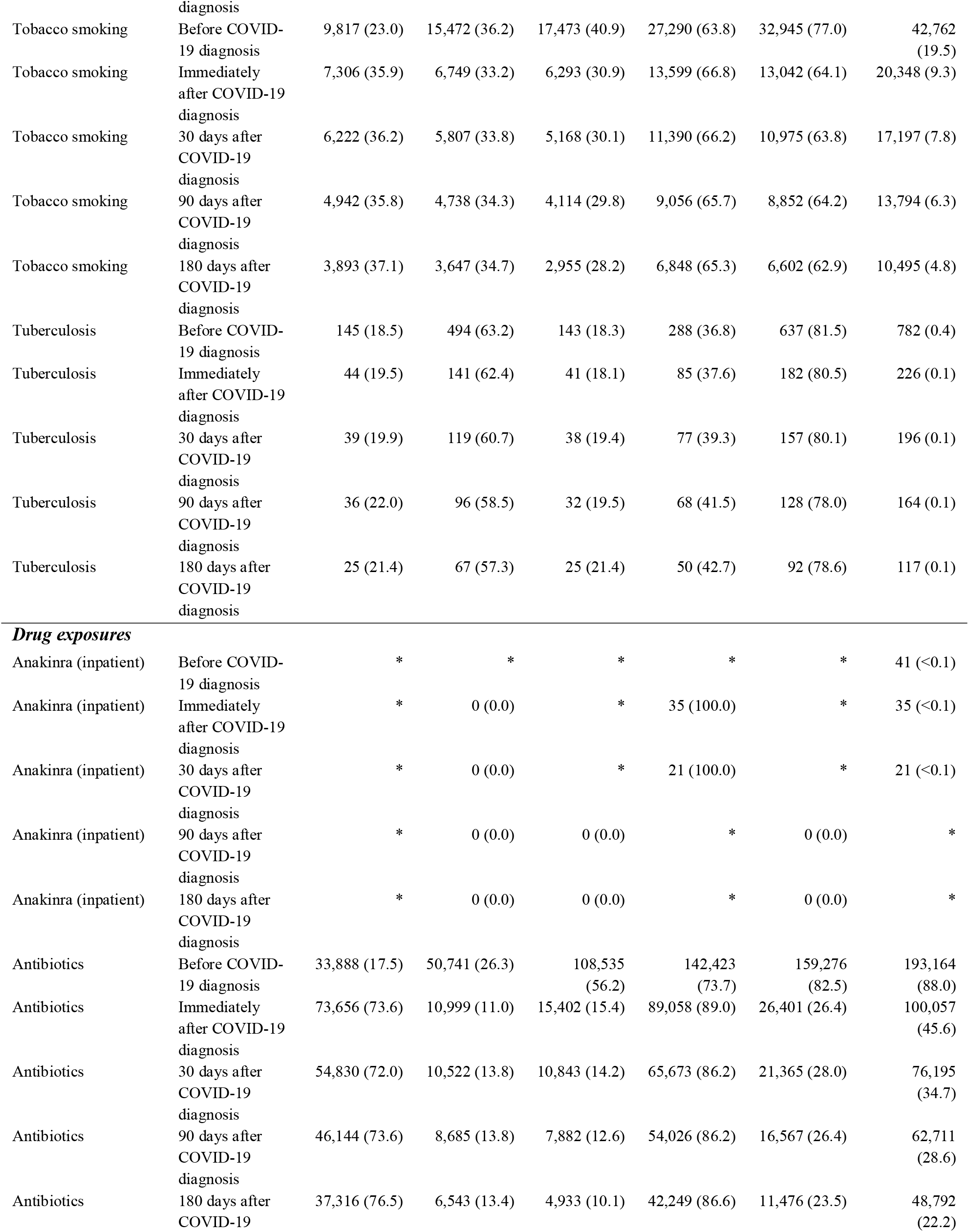
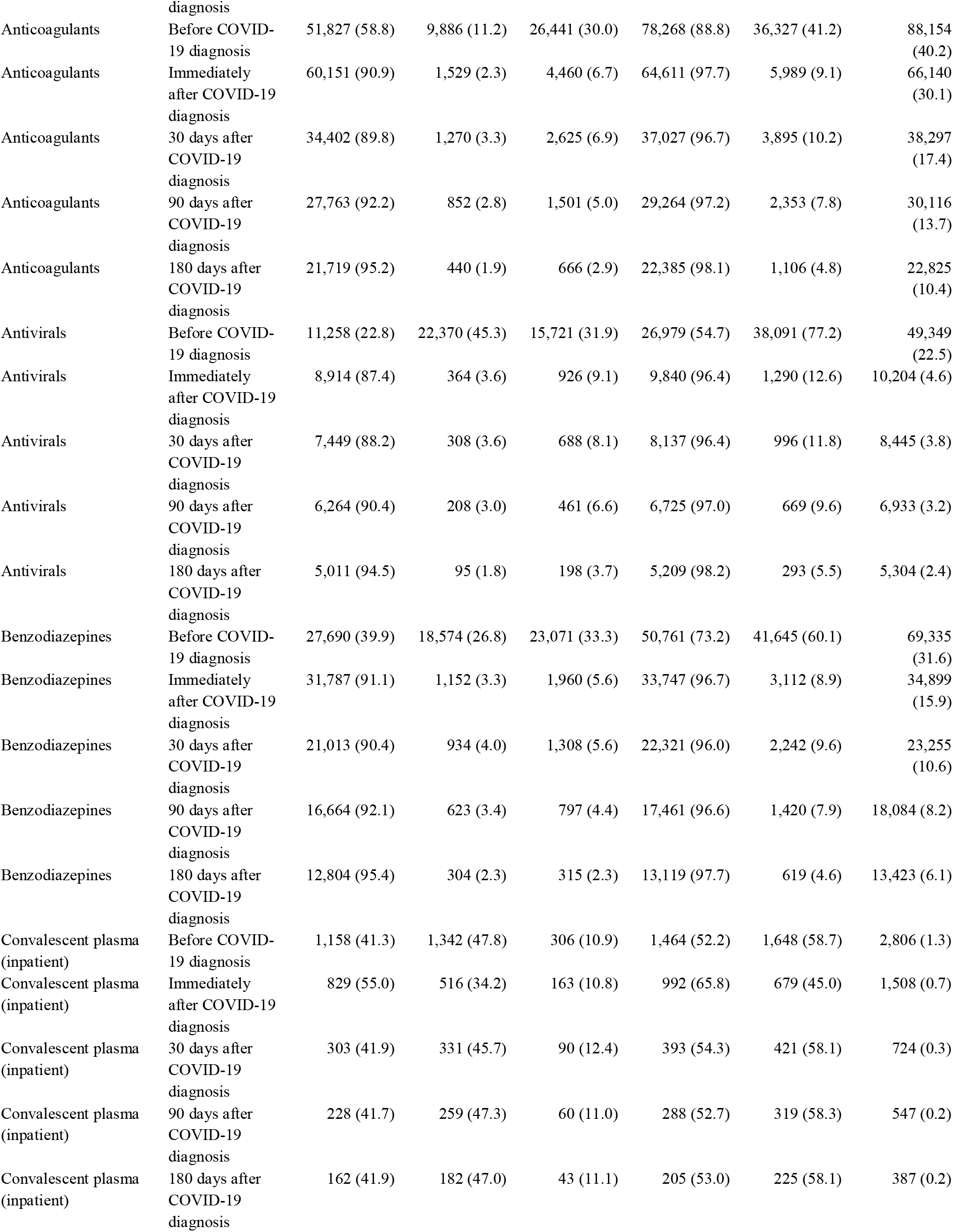
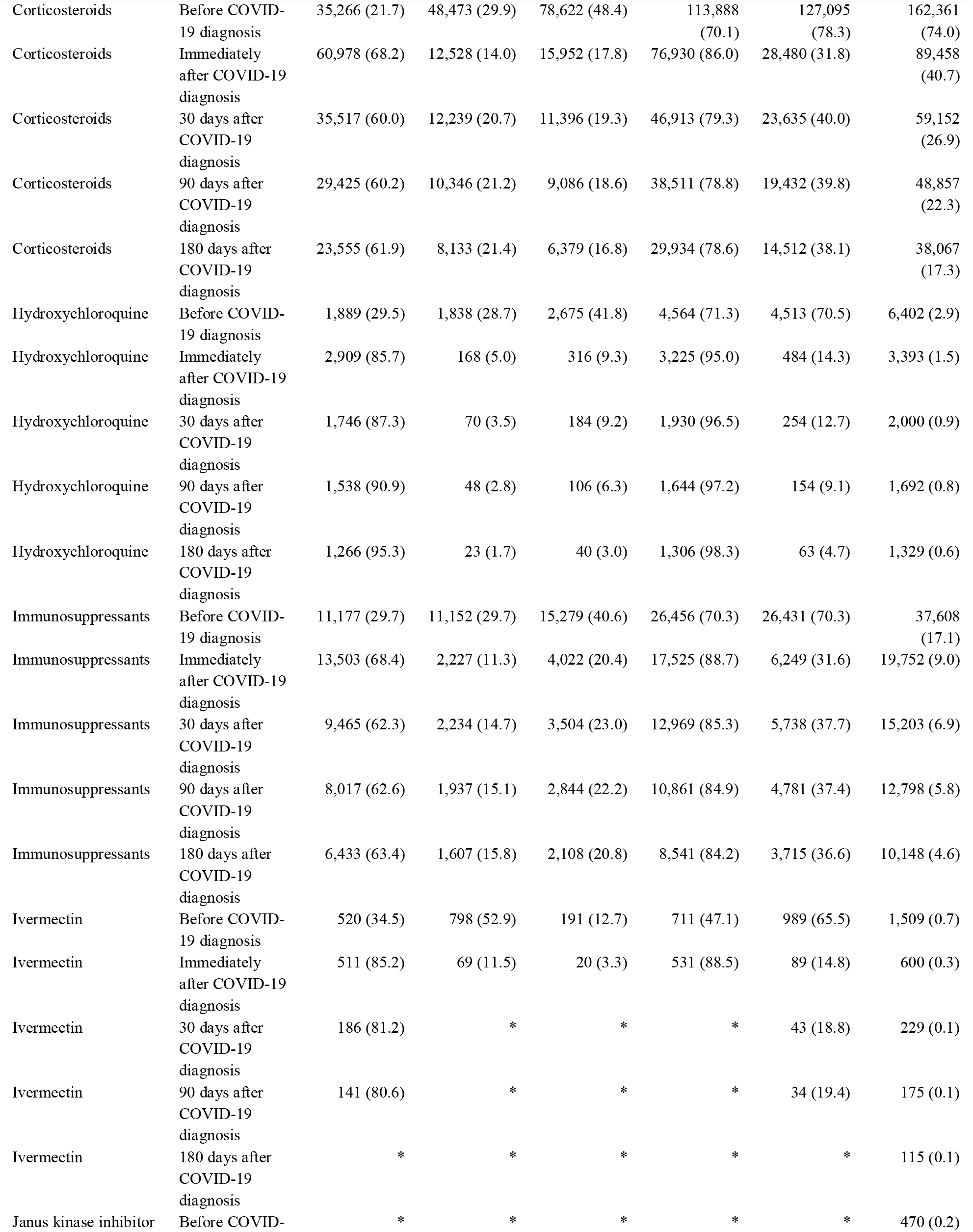
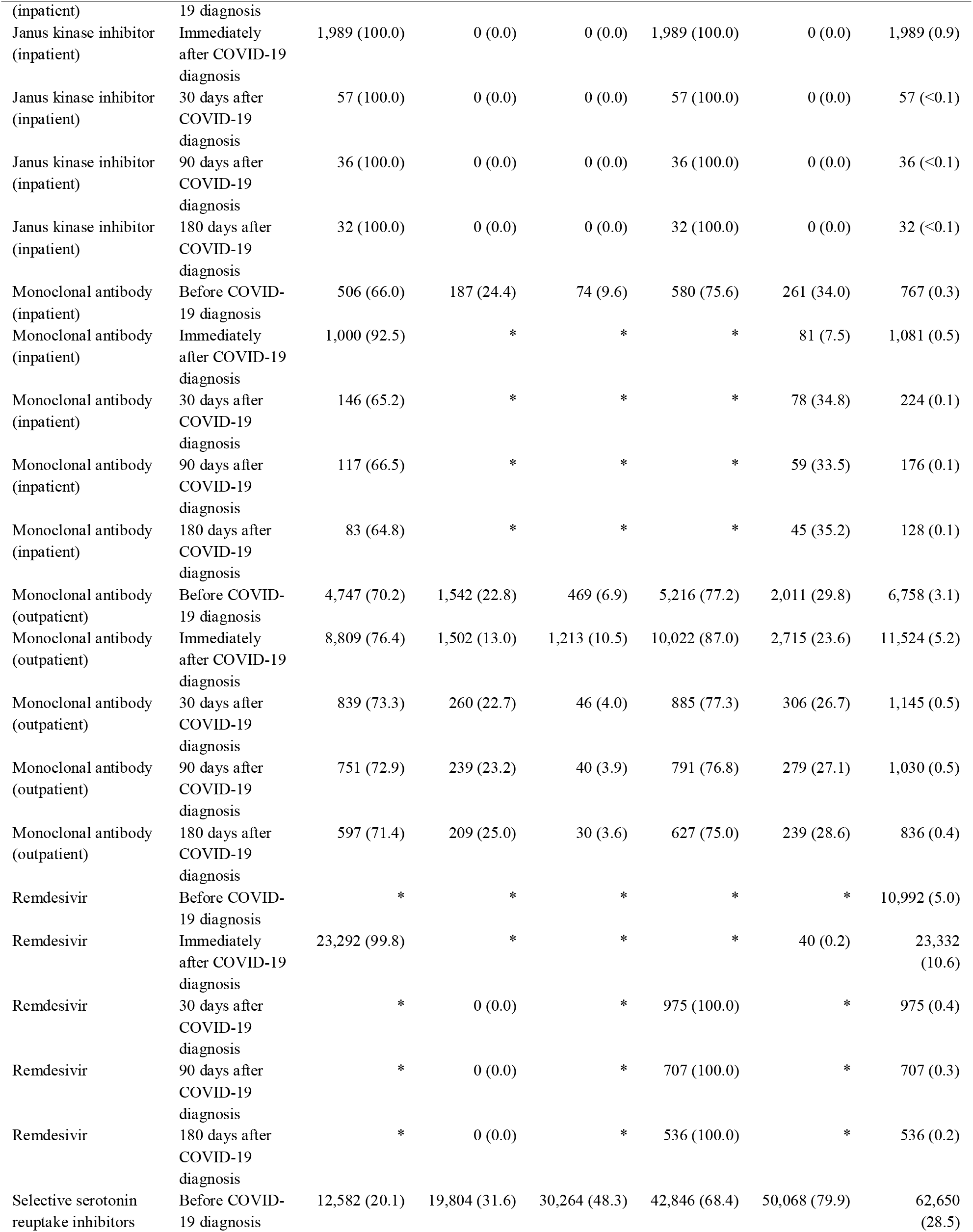
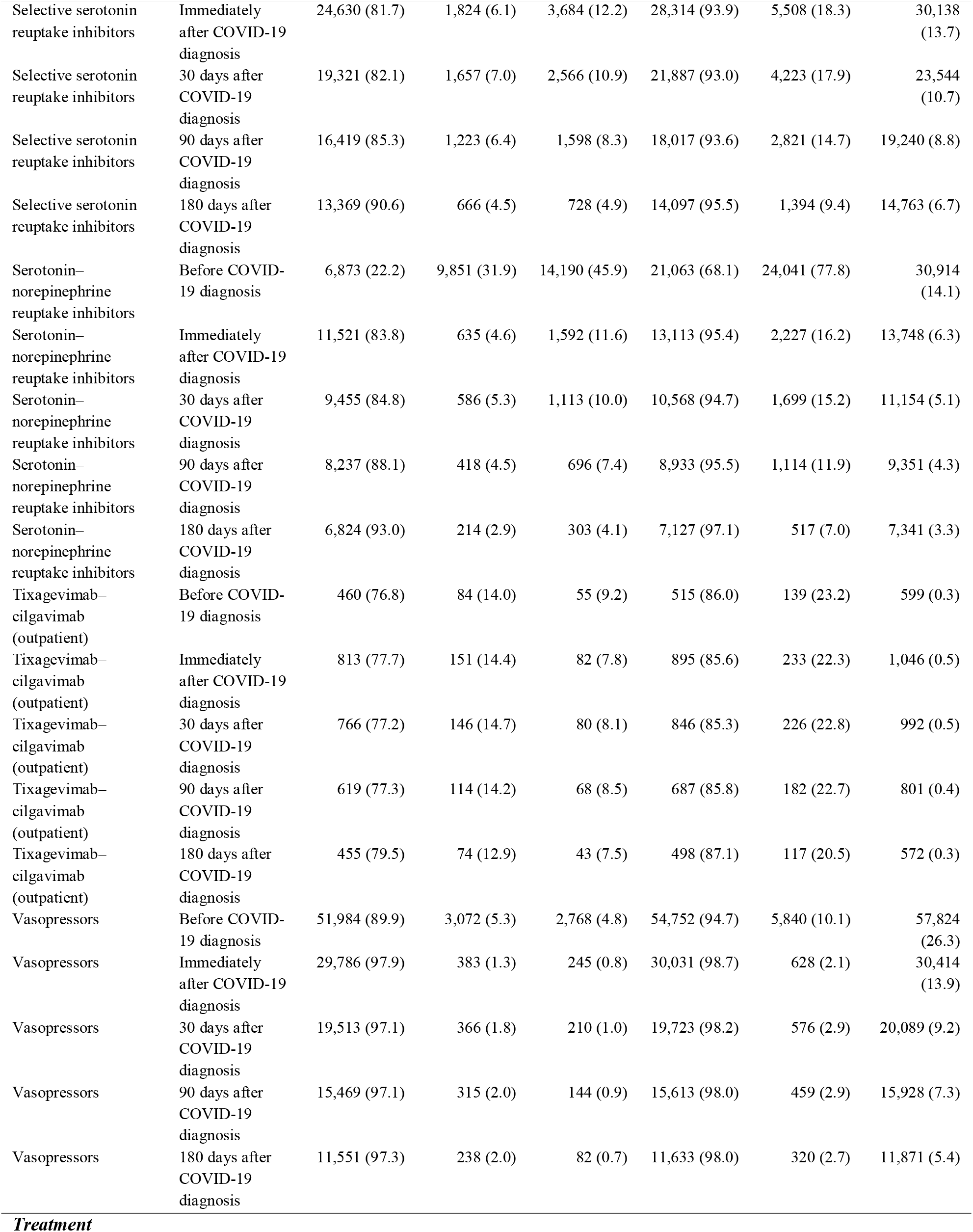
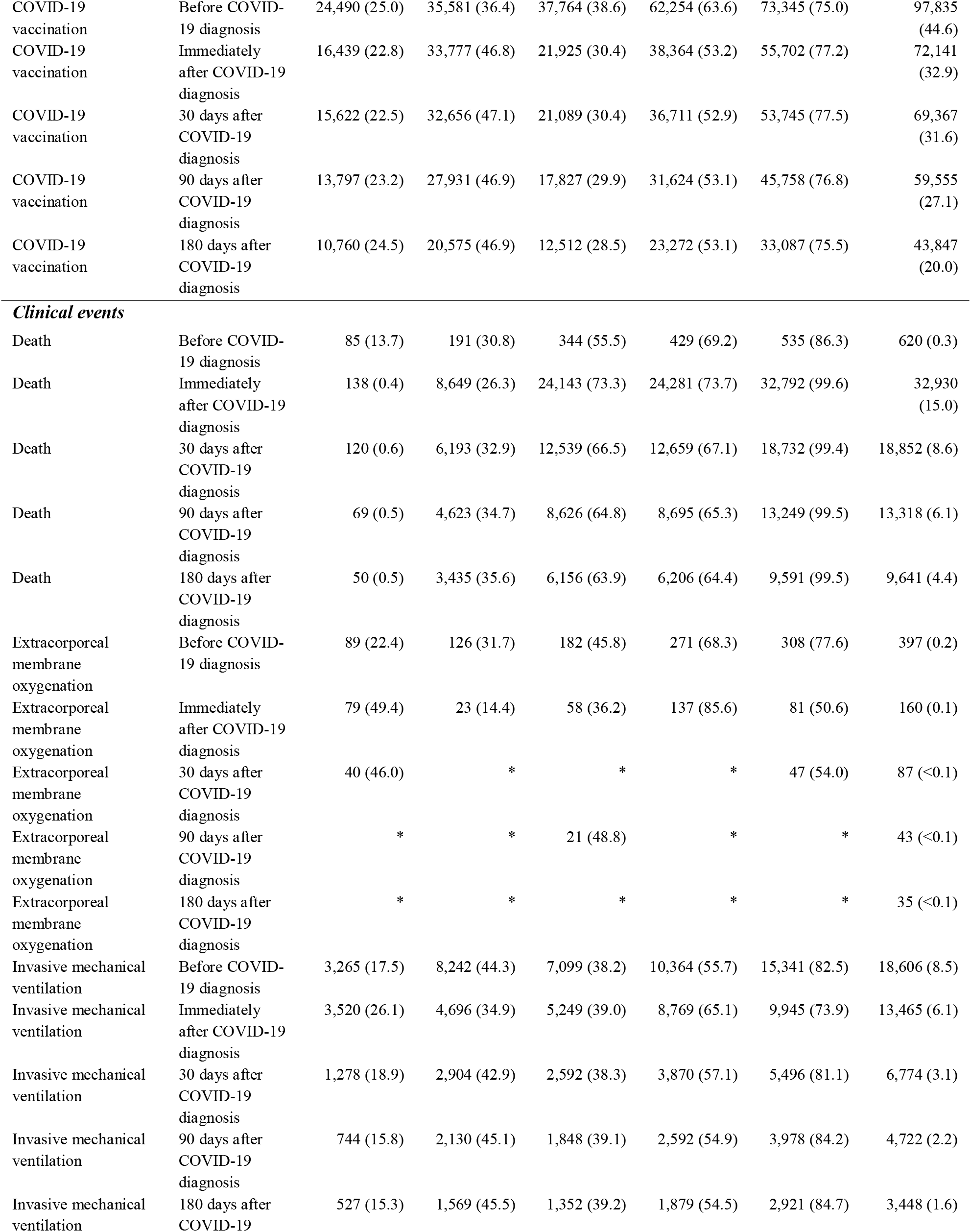
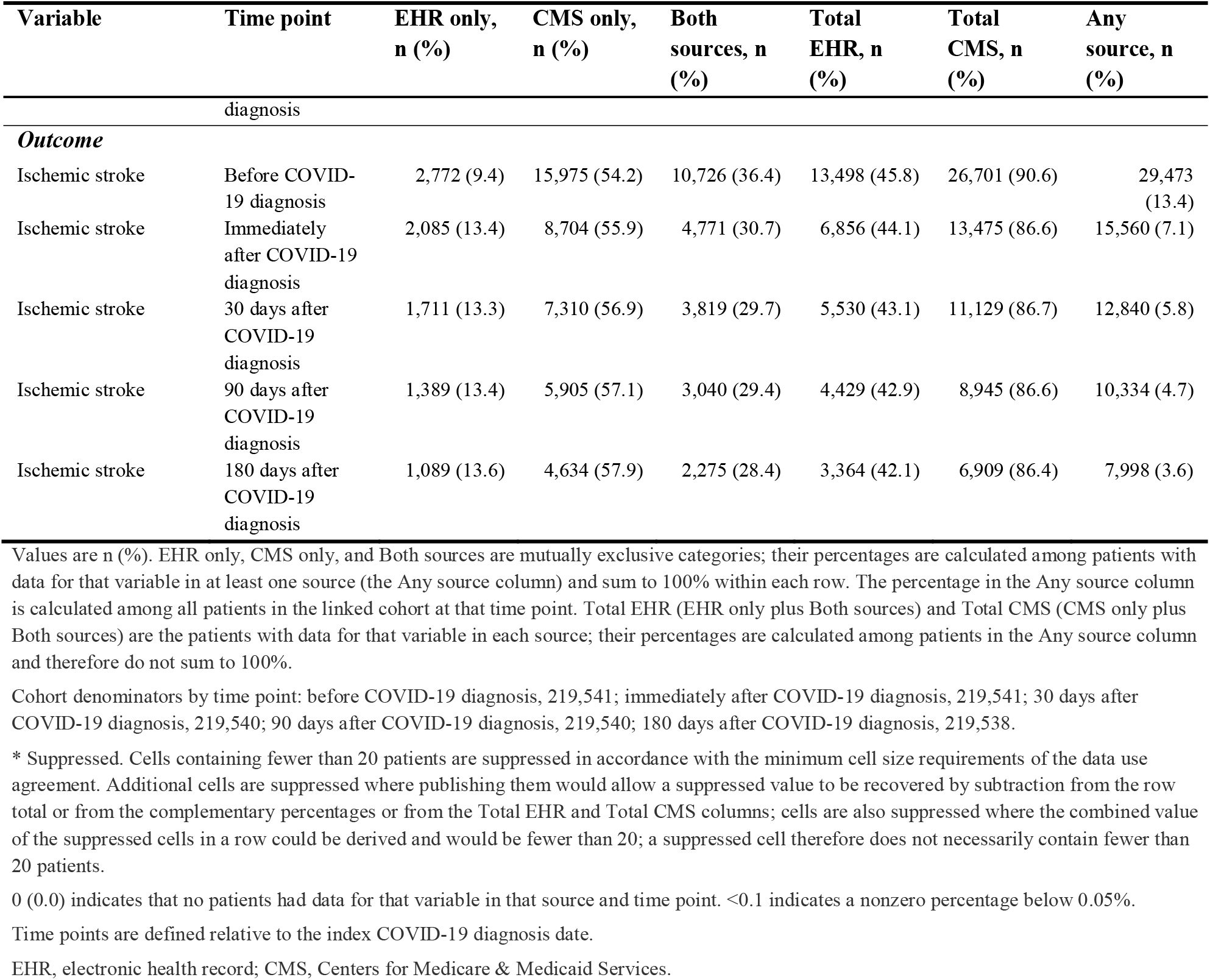
Availability of data for each study variable in electronic health record (EHR) data alone, Centers for Medicare & Medicaid Services (CMS) claims data alone, and in both sources, at five time points relative to COVID-19 diagnosis.

**Supplementary Table 3.**
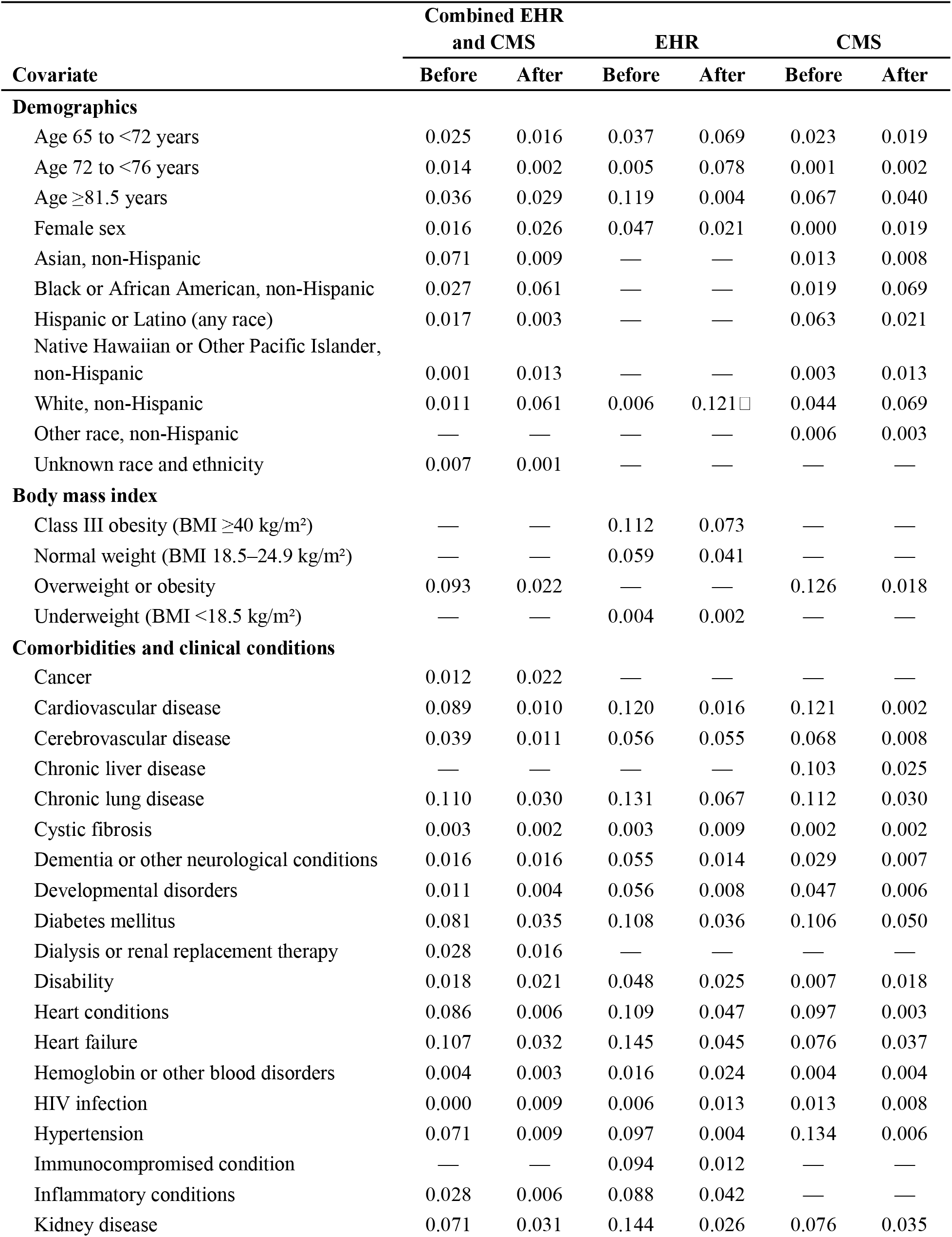
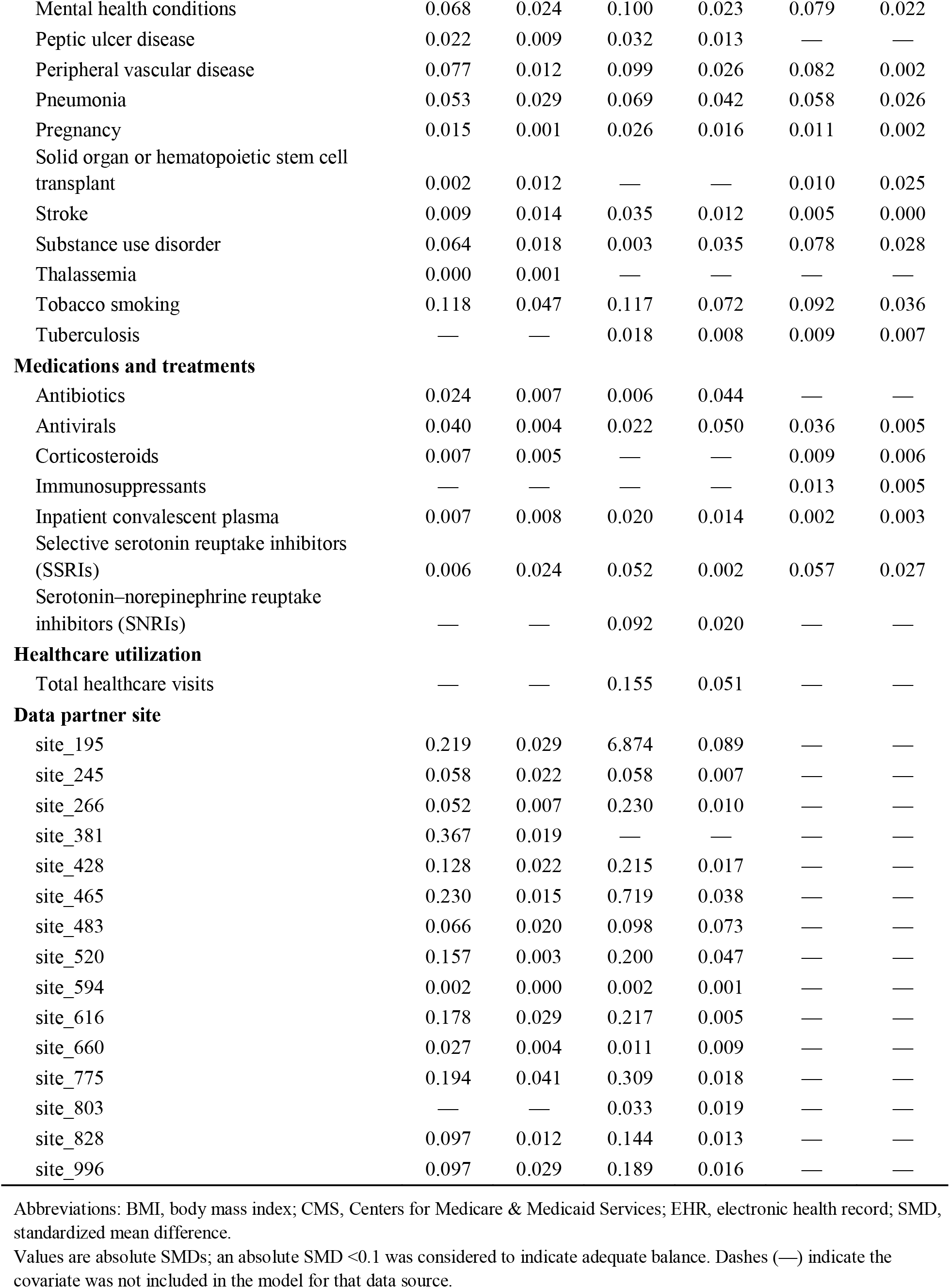
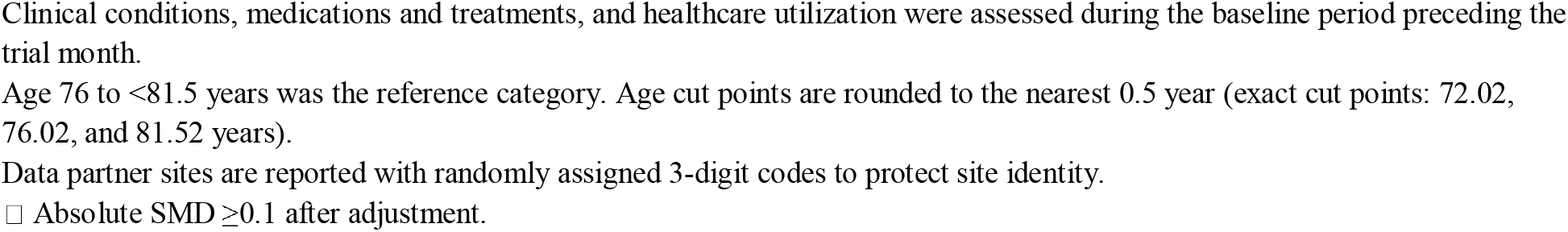
Absolute standardized mean differences for baseline covariates before and after adjustment, by data source.

| Covariate | Combined EHR<br>and CMS |  | EHR |  | CMS |  |
| --- | --- | --- | --- | --- | --- | --- |
|  | Before | After | Before | After | Before | After |
| **Demographics** |  |  |  |  |  |  |
| Age 65 to <72 years | 0.025 | 0.016 | 0.037 | 0.069 | 0.023 | 0.019 |
| Age 72 to <76 years | 0.014 | 0.002 | 0.005 | 0.078 | 0.001 | 0.002 |
| Age ≥81.5 years | 0.036 | 0.029 | 0.119 | 0.004 | 0.067 | 0.040 |
| Female sex | 0.016 | 0.026 | 0.047 | 0.021 | 0.000 | 0.019 |
| Asian, non-Hispanic | 0.071 | 0.009 | — | — | 0.013 | 0.008 |
| Black or African American, non-Hispanic | 0.027 | 0.061 | — | — | 0.019 | 0.069 |
| Hispanic or Latino (any race) | 0.017 | 0.003 | — | — | 0.063 | 0.021 |
| Native Hawaiian or Other Pacific Islander, non-Hispanic | 0.001 | 0.013 | — | — | 0.003 | 0.013 |
| White, non-Hispanic | 0.011 | 0.061 | 0.006 | 0.121□ | 0.044 | 0.069 |
| Other race, non-Hispanic | — | — | — | — | 0.006 | 0.003 |
| Unknown race and ethnicity | 0.007 | 0.001 | — | — | — | — |
| **Body mass index** |  |  |  |  |  |  |
| Class III obesity (BMI ≥40 kg/m<sup>2</sup>) | — | — | 0.112 | 0.073 | — | — |
| Normal weight (BMI 18.5–24.9 kg/m<sup>2</sup>) | — | — | 0.059 | 0.041 | — | — |
| Overweight or obesity | 0.093 | 0.022 | — | — | 0.126 | 0.018 |
| Underweight (BMI <18.5 kg/m<sup>2</sup>) | — | — | 0.004 | 0.002 | — | — |
| **Comorbidities and clinical conditions** |  |  |  |  |  |  |
| Cancer | 0.012 | 0.022 | — | — | — | — |
| Cardiovascular disease | 0.089 | 0.010 | 0.120 | 0.016 | 0.121 | 0.002 |
| Cerebrovascular disease | 0.039 | 0.011 | 0.056 | 0.055 | 0.068 | 0.008 |
| Chronic liver disease | — | — | — | — | 0.103 | 0.025 |
| Chronic lung disease | 0.110 | 0.030 | 0.131 | 0.067 | 0.112 | 0.030 |
| Cystic fibrosis | 0.003 | 0.002 | 0.003 | 0.009 | 0.002 | 0.002 |
| Dementia or other neurological conditions | 0.016 | 0.016 | 0.055 | 0.014 | 0.029 | 0.007 |
| Developmental disorders | 0.011 | 0.004 | 0.056 | 0.008 | 0.047 | 0.006 |
| Diabetes mellitus | 0.081 | 0.035 | 0.108 | 0.036 | 0.106 | 0.050 |
| Dialysis or renal replacement therapy | 0.028 | 0.016 | — | — | — | — |
| Disability | 0.018 | 0.021 | 0.048 | 0.025 | 0.007 | 0.018 |
| Heart conditions | 0.086 | 0.006 | 0.109 | 0.047 | 0.097 | 0.003 |
| Heart failure | 0.107 | 0.032 | 0.145 | 0.045 | 0.076 | 0.037 |
| Hemoglobin or other blood disorders | 0.004 | 0.003 | 0.016 | 0.024 | 0.004 | 0.004 |
| HIV infection | 0.000 | 0.009 | 0.006 | 0.013 | 0.013 | 0.008 |
| Hypertension | 0.071 | 0.009 | 0.097 | 0.004 | 0.134 | 0.006 |
| Immunocompromised condition | — | — | 0.094 | 0.012 | — | — |
| Inflammatory conditions | 0.028 | 0.006 | 0.088 | 0.042 | — | — |
| Kidney disease | 0.071 | 0.031 | 0.144 | 0.026 | 0.076 | 0.035 |
| Mental health conditions | 0.068 | 0.024 | 0.100 | 0.023 | 0.079 | 0.022 |
| Peptic ulcer disease | 0.022 | 0.009 | 0.032 | 0.013 | — | — |
| Peripheral vascular disease | 0.077 | 0.012 | 0.099 | 0.026 | 0.082 | 0.002 |
| Pneumonia | 0.053 | 0.029 | 0.069 | 0.042 | 0.058 | 0.026 |
| Pregnancy | 0.015 | 0.001 | 0.026 | 0.016 | 0.011 | 0.002 |
| Solid organ or hematopoietic stem cell transplant | 0.002 | 0.012 | — | — | 0.010 | 0.025 |
| Stroke | 0.009 | 0.014 | 0.035 | 0.012 | 0.005 | 0.000 |
| Substance use disorder | 0.064 | 0.018 | 0.003 | 0.035 | 0.078 | 0.028 |
| Thalassemia | 0.000 | 0.001 | — | — | — | — |
| Tobacco smoking | 0.118 | 0.047 | 0.117 | 0.072 | 0.092 | 0.036 |
| Tuberculosis | — | — | 0.018 | 0.008 | 0.009 | 0.007 |
| **Medications and treatments** |  |  |  |  |  |  |
| Antibiotics | 0.024 | 0.007 | 0.006 | 0.044 | — | — |
| Antivirals | 0.040 | 0.004 | 0.022 | 0.050 | 0.036 | 0.005 |
| Corticosteroids | 0.007 | 0.005 | — | — | 0.009 | 0.006 |
| Immunosuppressants | — | — | — | — | 0.013 | 0.005 |
| Inpatient convalescent plasma | 0.007 | 0.008 | 0.020 | 0.014 | 0.002 | 0.003 |
| Selective serotonin reuptake inhibitors (SSRIs) | 0.006 | 0.024 | 0.052 | 0.002 | 0.057 | 0.027 |
| Serotonin–norepinephrine reuptake inhibitors (SNRIs) | — | — | 0.092 | 0.020 | — | — |
| **Healthcare utilization** |  |  |  |  |  |  |
| Total healthcare visits | — | — | 0.155 | 0.051 | — | — |
| **Data partner site** |  |  |  |  |  |  |
| site_195 | 0.219 | 0.029 | 6.874 | 0.089 | — | — |
| site_245 | 0.058 | 0.022 | 0.058 | 0.007 | — | — |
| site_266 | 0.052 | 0.007 | 0.230 | 0.010 | — | — |
| site_381 | 0.367 | 0.019 | — | — | — | — |
| site_428 | 0.128 | 0.022 | 0.215 | 0.017 | — | — |
| site_465 | 0.230 | 0.015 | 0.719 | 0.038 | — | — |
| site_483 | 0.066 | 0.020 | 0.098 | 0.073 | — | — |
| site_520 | 0.157 | 0.003 | 0.200 | 0.047 | — | — |
| site_594 | 0.002 | 0.000 | 0.002 | 0.001 | — | — |
| site_616 | 0.178 | 0.029 | 0.217 | 0.005 | — | — |
| site_660 | 0.027 | 0.004 | 0.011 | 0.009 | — | — |
| site_775 | 0.194 | 0.041 | 0.309 | 0.018 | — | — |
| site_803 | — | — | 0.033 | 0.019 | — | — |
| site_828 | 0.097 | 0.012 | 0.144 | 0.013 | — | — |
| site_996 | 0.097 | 0.029 | 0.189 | 0.016 | — | — |

Abbreviations: BMI, body mass index; CMS, Centers for Medicare & Medicaid Services; EHR, electronic health record; SMD, standardized mean difference.
Values are absolute SMDs; an absolute SMD <0.1 was considered to indicate adequate balance. Dashes (—) indicate the covariate was not included in the model for that data source.
Clinical conditions, medications and treatments, and healthcare utilization were assessed during the baseline period preceding the trial month.
Age 76 to <81.5 years was the reference category. Age cut points are rounded to the nearest 0.5 year (exact cut points: 72.02, 76.02, and 81.52 years).
Data partner sites are reported with randomly assigned 3-digit codes to protect site identity.
□ Absolute SMD $\geq 0.1$ after adjustment.

## Notes

### Competing Interest Statement

The authors have declared no competing interest.

### Author Declarations

Use of N3C data for this study does not involve human subjects (45 CFR 46.102) as determined by the NIH Office of IRB Operations. Thus, this study did not include factors necessitating patient consent. The N3C data transfer to the National Centre for Advancing Translational Sciences (NCATS) is performed under a Johns Hopkins University Reliance Protocol #IRB00249128 or individual site agreements with the National Institutes of Health (NIH).

## References

1. Young JC, Dasgupta N, Stürmer T, Pate V, Jonsson Funk M. Considerations for observational study design: Comparing the evidence of opioid use between electronic health records and insurance claims. Pharmacoepidemiology and drug safety. 2022;31(8):913–20.

2. Hippisley-Cox J, Patone M, Mei XW, Saatci D, Dixon S, Khunti K, et al. Risk of thrombocytopenia and thromboembolism after covid-19 vaccination and SARS-CoV-2 positive testing: self-controlled case series study. BMJ (Clinical research ed). 2021;374:n1931.

3. Lin KJ, Singer DE, Glynn RJ, Murphy SN, Lii J, Schneeweiss S. Identifying Patients With High Data Completeness to Improve Validity of Comparative Effectiveness Research in Electronic Health Records Data. Clin Pharmacol Ther. 2018;103(5):899–905.

4. Lin K, Glynn R, Singer D, Murphy S, Lii J, Schneeweiss S. Out-of-system Care and Recording of Patient Characteristics Critical for Comparative Effectiveness Research. Epidemiology (Cambridge, Mass). 2018;29(3):356–63.

5. Datavant. What is Claims Data and Its Advantages and Disadvantages? 2023 [Available from: https://www.datavant.com/real-world-data-rwd/claims-data.

6. Kelley AS, Hanson LC, Ast K, Ciemins EL, Dunning SC, Meskow C, et al. The Serious Illness Population: Ascertainment via Electronic Health Record or Claims Data. J Pain Symptom Manage. 2021;62(3):e148–e55.

7. Schneeweiss S, Desai RJ, Ball R. A future of data-rich pharmacoepidemiology studies: transitioning to large-scale linked electronic health record + claims data. American journal of epidemiology. 2025;194(2):315–21.

8. Mahmoudi E, Kotsis SV, Chung KC. A Review of the Use of Medicare Claims Data in Plastic Surgery Outcomes Research. Plast Reconstr Surg Glob Open. 2015;3(10):e530.

9. Konrad R, Zhang W, Bjarndóttir M, Proaño R. Key considerations when using health insurance claims data in advanced data analyses: an experience report. Health Syst (Basingstoke). 2019;9(4):317–25.

10. Franklin JM, Gopalakrishnan C, Krumme AA, Singh K, Rogers JR, Kimura J, et al. The relative benefits of claims and electronic health record data for predicting medication adherence trajectory. American Heart Journal. 2018;197:153–62.

11. Li X, Cole SR, Westreich D, Brookhart MA. Primary non-adherence and the new-user design. Pharmacoepidemiology and drug safety. 2018;27(4):361–4.

12. Fischer MA, Stedman MR, Lii J, Vogeli C, Shrank WH, Brookhart MA, et al. Primary Medication Non-Adherence: Analysis of 195,930 Electronic Prescriptions. Journal of general internal medicine. 2010;25(4):284–90.

13. Barda N, Dagan N, Ben-Shlomo Y, Kepten E, Waxman J, Ohana R, et al. Safety of the BNT162b2 mRNA Covid-19 Vaccine in a Nationwide Setting. New England Journal of Medicine. 2021;385(12):1078–90.

14. Simpson CR, Shi T, Vasileiou E, Katikireddi SV, Kerr S, Moore E, et al. First-dose ChAdOx1 and BNT162b2 COVID-19 vaccines and thrombocytopenic, thromboembolic and hemorrhagic events in Scotland. Nature medicine. 2021;27(7):1290–7.

15. Petersen I, Douglas I, Whitaker H. Self controlled case series methods: an alternative to standard epidemiological study designs. 2016;354:i4515.

16. Hong G. Causality in a social world : moderation, meditation and spill-over. Chichester, England: Wiley; 2015. p. 76–99.

17. Ihle-Hansen H, Bøås H, Tapia G, Hagberg G, Ihle-Hansen H, Berild JD, et al. Stroke After SARS-CoV-2 mRNA Vaccine: A Nationwide Registry Study. Stroke. 2023;54(5):e190–e3.

18. Haendel MA, Chute CG, Bennett TD, Eichmann DA, Guinney J, Kibbe WA, et al. The National COVID Cohort Collaborative (N3C): Rationale, design, infrastructure, and deployment. Journal of the American Medical Informatics Association. 2021;28(3):427–43.

19. Centers for Medicare and Medicaid Services. Medicare Monthly Enrollment 2026 [Available from: https://data.cms.gov/summary-statistics-on-beneficiary-enrollment/medicare-and-medicaid-reports/medicare-monthly-enrollment.

20. Fu EL. Target Trial Emulation to Improve Causal Inference from Observational Data: What, Why, and How? Journal of the American Society of Nephrology. 2023;34(8):1305–14.

21. Hong G. Marginal mean weighting through stratification: a generalized method for evaluating multivalued and multiple treatments with nonexperimental data. Psychological methods. 2012;17(1):44–60.

22. Austin PC. Advances in propensity score analysis. Statistical methods in medical research. 2020;29(3):641–3.

23. Farrington CP, Whitaker HJ, Hocine MN. Case series analysis for censored, perturbed, or curtailed post-event exposures. Biostatistics. 2009;10(1):3–16.

24. Cadarette SM, Maclure M, Delaney JAC, Whitaker HJ, Hayes KN, Wang SV, et al. Control yourself: ISPE-endorsed guidance in the application of self-controlled study designs in pharmacoepidemiology. Pharmacoepidemiology and drug safety. 2021;30(6):671–84.

25. Zaidan M, Puebla Neira D, Polychronopoulou E, Yong-Fang K, Sharma G. Healthcare utilization 9 months pre- and post-COVID-19 hospitalization among patients discharged alive. PloS one. 2024;19(6):e0303509.

26. Agarwal R, Liao JM, Gupta A, Navathe AS. The Impact Of Bundled Payment On Health Care Spending, Utilization, And Quality: A Systematic Review. Health Aff (Millwood). 2020;39(1):50–7.

27. Carroll NM, Ellis JL, Luckett CF, Raebel MA. Improving the validity of determining medication adherence from electronic health record medications orders. Journal of the American Medical Informatics Association : JAMIA. 2011;18(5):717–20.

28. Sattler ELP, Lee JS, Perri M. Medication (Re)fill Adherence Measures Derived from Pharmacy Claims Data in Older Americans: A Review of the Literature. Drugs & Aging. 2013;30(6):383–99.

29. HHS Officer of Inspector General. Medicare Advantage Compliance Audit of Specific Diagnosis Codes That HealthAssurance Pennsylvania, Inc. (Contract H5522) Submitted to CMS2024 August, 2024. Available from: https://oig.hhs.gov/documents/audit/10007/A-05-22-00020RIB.pdf.

30. Centers for Medicare and Medicaid Services. Documentation Integrity in Electronic Health Records2016 August, 2026. Available from: https://www.cms.gov/files/document/ehrdocumentationfs062816pdf.

31. HHS Officer of Inspector General. Medicare Advantage: Questionable Use of Health Risk Assessments Continues To Drive Up Payments to Plans by Billions 2024 August, 2026. Available from: https://oig.hhs.gov/documents/evaluation/10029/OEI-03-23-00380-highlights.pdf.

32. Stefanou M-I, Palaiodimou L, Aguiar de Sousa D, Theodorou A, Bakola E, Katsaros DE, et al. Acute Arterial Ischemic Stroke Following COVID-19 Vaccination. A Systematic Review and Meta-analysis. 2022;99(14):e1465–e74.

33. Torabi F, Bedston S, Lowthian E, Akbari A, Owen RK, Bradley DT, et al. Risk of thrombocytopenic, haemorrhagic and thromboembolic disorders following COVID-19 vaccination and positive test: a self-controlled case series analysis in Wales. Scientific Reports. 2022;12(1):16406.

34. Wong HL, Hu M, Zhou CK, Lloyd PC, Amend KL, Beachler DC, et al. Risk of myocarditis and pericarditis after the COVID-19 mRNA vaccination in the USA: a cohort study in claims databases. Lancet (London, England). 2022;399(10342):2191–9.

35. Lu Y, Matuska K, Nadimpalli G, Ma Y, Duma N, Zhang HT, et al. Stroke Risk After COVID-19 Bivalent Vaccination Among US Older Adults. JAMA. 2024;331(11):938–50.

36. Devoe JE, Gold R, McIntire P, Puro J, Chauvie S, Gallia CA. Electronic health records vs Medicaid claims: completeness of diabetes preventive care data in community health centers. Ann Fam Med. 2011;9(4):351–8.

37. Schneider KL, Bell EJ, Zhou CK, Yang G, Lloyd P, Clarke TC, et al. Use of Immunization Information Systems in Ascertainment of COVID-19 Vaccinations for Claims-Based Vaccine Safety and Effectiveness Studies. JAMA network open. 2023;6(5):e2313512.

38. Jackson LA, Jackson ML, Nelson JC, Neuzil KM, Weiss NS. Evidence of bias in estimates of influenza vaccine effectiveness in seniors. International journal of epidemiology. 2006;35(2):337–44.

39. Whitaker HJ, Farrington CP, Spiessens B, Musonda P. Tutorial in biostatistics: the self-controlled case series method. Statistics in medicine. 2006;25(10):1768–97.

40. Fine PEM, Chen RT. Confounding in Studies of Adverse Reactions to Vaccines. American journal of epidemiology. 1992;136(2):121–35.

41. Rahmig J, Altarsha E, Siepmann T, Barlinn K. Acute Ischemic Stroke in the Context of SARS-CoV-2 Vaccination: A Systematic Review. Neuropsychiatric disease and treatment. 2022;18:1907–16.

42. Farrington P. The self-controlled case series method and covid-19. BMJ (Clinical research ed). 2022;377:o625.

43. Klein NP, Lewis N, Goddard K, Fireman B, Zerbo O, Hanson KE, et al. Surveillance for Adverse Events After COVID-19 mRNA Vaccination. JAMA. 2021;326(14):1390–9.

44. Botton J, Jabagi MJ, Bertrand M, Baricault B, Drouin J, Le Vu S, et al. Risk for Myocardial Infarction, Stroke, and Pulmonary Embolism Following COVID-19 Vaccines in Adults Younger Than 75 Years in France. Annals of internal medicine. 2022;175(9):1250–7.

45. Shrank WH, Patrick AR, Brookhart MA. Healthy user and related biases in observational studies of preventive interventions: a primer for physicians. Journal of general internal medicine. 2011;26(5):546–50.

46. Saelee R, Zell E, Murthy BP, Castro-Roman P, Fast H, Meng L, et al. Disparities in COVID-19 Vaccination Coverage Between Urban and Rural Counties - United States, December 14, 2020-January 31, 2022. MMWR Morbidity and mortality weekly report. 2022;71(9):335–40.

47. Centers for Medicare and Medicaid Services. Assessing the Completeness of Medicare Claims Data for Measuring COVID-19 Vaccine Administration. Report. Baltimore, MD: Centers for Medicare and Medicaid Services; 2022.

48. Wong H-L, Tworkoski E, Ke Zhou C, Hu M, Thompson D, Lufkin B, et al. Surveillance of COVID-19 vaccine safety among elderly persons aged 65 years and older. Vaccine. 2023;41(2):532–9.

